# Cell type-specific contextualisation of the human phenome: towards the systematic treatment of all rare diseases

**DOI:** 10.1101/2023.02.13.23285820

**Authors:** Brian M. Schilder, Kitty B. Murphy, Hiranyamaya Dash, Yichun Zhang, Robert Gordon-Smith, Jai Chapman, Momoko Otani, Nathan G. Skene

## Abstract

Rare diseases (RDs) are an extremely heterogeneous and underserved category of medical conditions. While the majority of RDs are strongly genetic, it remains largely unknown via which physiological mechanisms genetics cause RD. Therefore, we sought to systematically characterise the cell type-specific mechanisms underlying all RD phenotypes with a known genetic cause by leveraging the Human Phenotype Ontology and transcriptomic single-cell atlases of the entire human body from embryonic, foetal, and adult samples. In total we identified significant associations between 201 cell types and 9,575/11,028 (86.7%) unique phenotypes across 8,628 RDs. This greatly the collective knowledge of RD phenotype-cell type mechanisms. Next, we sought to systematically identify phenotypes in which the application of these results would have the greatest clinical impact based on metrics of severity (e.g. lethality, motor/mental impairment) and compatibility with gene therapy (e.g. filtering out physical malformations). Furthermore, we have made these results entirely reproducible and freely accessible to the global community to maximise their impact, including an interactive web portal (https://neurogenomics-ukdri.dsi.ic.ac.uk/). To summarise, this work represents a significant step forward in the mission to treat patients across an extremely diverse spectrum of serious RDs.

## Introduction

While rare diseases (RDs) are individually uncommon, they collectively account for an enormous global disease burden with over 10,000 recognised RDs affecting at least 300-400 million people globally^1^ (1 in 10-20 people)^2^. Over 75% of RDs primarily affect children with a 30% mortality rate by five years of age^3^. Despite the prevalence and severity of RDs, patients suffering from these conditions are vastly underserved due to several contributing factors. First, diagnosis is extremely challenging due to the highly variable clinical presentations of many of these diseases. The diagnostic odyssey can take patients and their families decades, with an average time to diagnosis of five years^4^. Of those, ∼46% receive at least one incorrect diagnosis and over 75% of all patients never receive any diagnosis^5^. Second, prognosis is also made difficult by high variability in disease course and outcomes which makes matching patients with effective and timely treatment plans even more challenging. Finally, even for patients who receive an accurate diagnosis/prognosis, treatments are currently only available for less than 5% of all RDs^6^. In addition to the scientific challenges of understanding RDs, there are strong financial disincentives for pharmaceutical and biotechnology companies to develop expensive therapeutics for exceedingly small RD patient populations with little or no return on investment^7,8^. Those that have been produced are amongst the world’s most expensive drugs, greatly limiting patients’ ability to access it^9,10^. New high-throughput approaches for the development of rare disease therapeutics could greatly reduce costs (for manufacturers and patients) and accelerate the timeline from discovery to delivery.

A major challenge in both healthcare and scientific research is the lack of standardised medical terminology. Even in the age of electronic healthcare records (EHR) much of the information about an individual’s history is currently fractured across healthcare providers, often with differing nomenclatures for the same conditions. The Human Phenotype Ontology (HPO) is a hierarchically organised set of controlled clinical terms that provides a much needed common framework by which clinicians and researchers can precisely communicate patient conditions^14^. The HPO spans all domains of human physiology and currently describes 18,082 phenotypes across 10,300 RDs. Each phenotype and disease is assigned its own unique identifier and organised as a hierarchical graph, such that higher-level terms describe broad phenotypic categories or *branches* (e.g. *HP:0033127* : ‘Abnormality of the musculoskeletal system’ which contains 4,495 unique phenotypes) and lower-level terms describe increasingly precise phenotypes (e.g. *HP:0030675*: ‘Contracture of proximal interphalangeal joints of 2nd-5th fingers’). It has already been integrated into healthcare systems and clinical diagnostic tools around the world, with increasing adoption over time^11^. Standardised frameworks like the HPO also allow us to aggregate relevant knowledge about the molecular mechanisms underlying each RD.

Over 80% of RDs have a known genetic cause^15,16^. Since 2008, the HPO has been continuously updated using curated knowledge from the medical literature, as well as by integrating databases of expert validated gene-phenotype relationships, such as OMIM^17–19^, Orphanet^20,21^, and DECIPHER^22^. Mappings between HPO terms to other commonly used medical ontologies (e.g. SNOMED CT^23^, UMLS^24,25^, ICD-9/10/11^26^) make the HPO even more valuable as a clinical resource (provided in Mappings section of Methods). Many of these gene annotations are manually or semi-manually curated by expert clinicians from case reports of rare disease patients in which the causal gene is identified through whole exome or genome sequencing. Currently, the HPO contains gene annotations for 11,047 phenotypes across 8,631 diseases. Yet genes alone do not tell the full story of how RDs come to be, as their expression and functional relevance varies drastically across the multitude of tissues and cell types contained within the human body. Our knowledge of the physiological mechanisms via which genetics cause pathogenesis is lacking for most RDs, severely hindering our ability to effectively diagnose, prognose and treat RD patients.

Our knowledge of cell type-specific biology has exploded over the course of the last decade and a half, with numerous applications in both scientific and clinical practices^27–29^. In particular, single-cell RNA-seq (scRNA-seq) has allowed us to quantify the expression of every gene (i.e. the transcriptome) in individual cells. More recently, comprehensive single-cell transcriptomic atlases across tissues have also emerged^30,31^. In particular, the Descartes Human^32^ and Human Cell Landscape^33^ projects provide comprehensive multi-system scRNA-seq atlases in embryonic, foetal, and adult human samples from across the human body. These datasets provide data-driven gene signatures for hundreds of cell subtypes. Given that many disease-associated genes are expressed in some cell types but not others, we can infer that disruptions to these genes will have varying impact across cell types. By comparing the aggregated disease gene annotations with cell type-specific expression profiles, we can therefore uncover the cell types and tissues via which diseases mediate their effects.

Here, we combine and extend several of the most comprehensive genomic and transcriptomic resources currently available to systematically uncover the cell types underlying granular phenotypes across 8,628 diseases Fig. 1. Conversely, this approach also allows us to better understand the roles of understudied cell types by observing which phenotypes they tend to associate this. For example, the original authors proposed that a novel class *AFB*+/*ALB*+ cells may represent hepatoblasts circulating through the bloodstream during foetal development^34^. Our results support this hypothesis as *AFB*+/*ALB*+ cells were significantly associated with 12 liver-related phenotypes, as well as 58 blood-related phenotypes.

**Figure 1:**
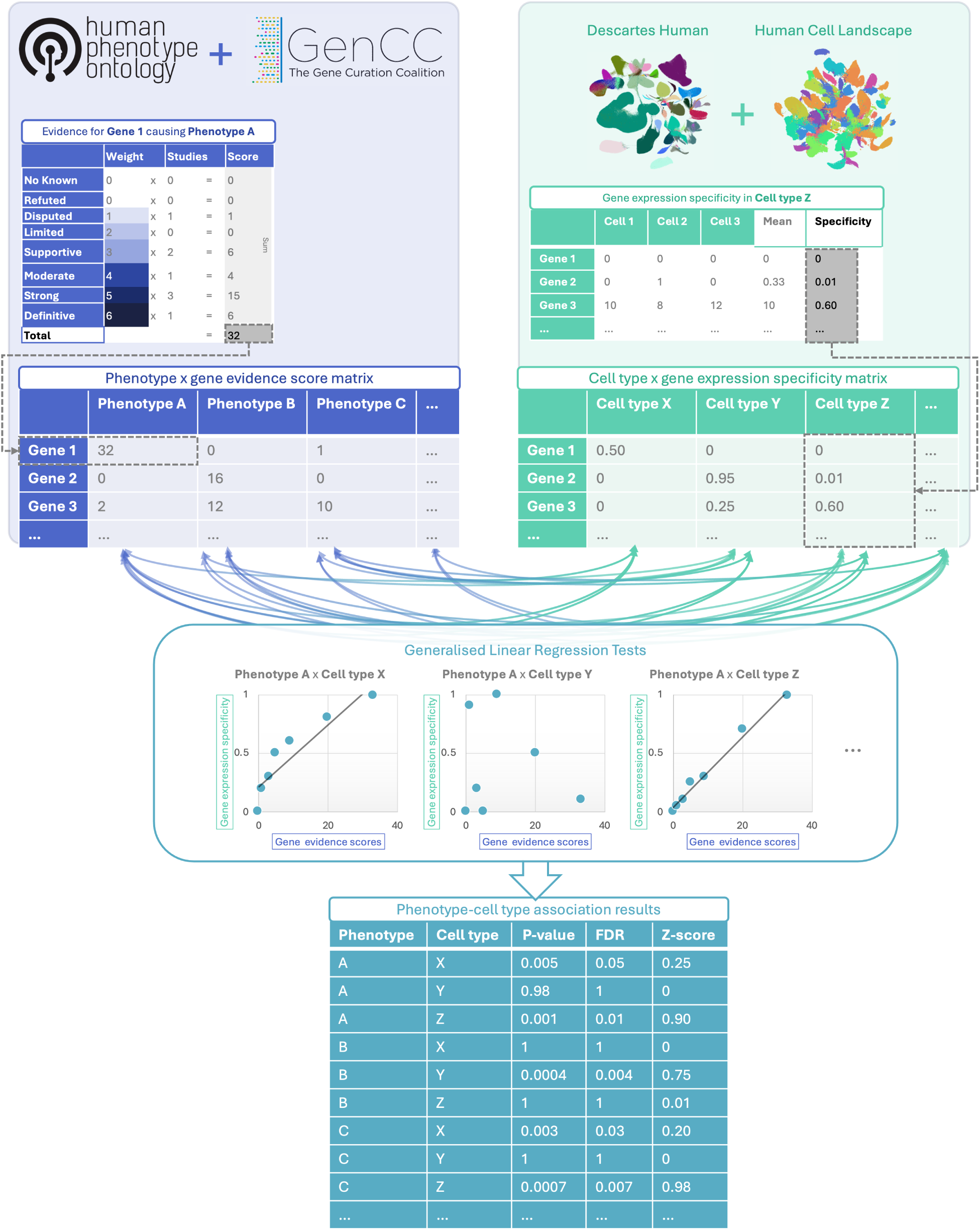
Multi-modal data fusion reveals the cell types underlying thousands of human phenotypes. Schematic overview of study design in which we numerically encoded the strength of evidence linking each gene and each phenotype (using the Human Phenotype Ontology and GenCC databases). We then created gene signature profiles for all cell types in the Descartes Human and Human Cell Landscape scRNA-seq atlases. Finally, we iteratively ran generalised linear regression tests between all pairwise combinations of phenotype gene signatures and cell type gene signatures. The resulting associations were then used to nominate cell type-resolved gene therapy targets for thousands of rare diseases.

Beyond making discoveries in basic science, our phenome-wide cell type associations provide essential context for the development of novel therapeutics, especially gene therapy modalities such as adeno-associated viral (AAV) vectors in which advancement have been made in their ability selectively target specific cell types^35,36^. Precise knowledge of relevant cell types and tissues causing the disease can improve safety by minimising harmful side effects in off-target cell types and tissues. It can also enhance efficacy by efficiently delivering expensive therapeutic payloads to on-target cell types and tissues. For example, if a phenotype primarily effects retinal cells, then the gene therapy would be optimised for delivery to retinal cells of the eye. Using this information, we developed a high-throughput pipeline for comprehensively nominating cell type-resolved gene therapy targets across thousands of RD phenotypes. As a prioritisation tool, we sorted these targets based on the severity of their respective phenotypes, using a generative AI-based approach^37^. Together, our study dramatically expands the available knowledge of the cell types, organ systems and life stages underlying RD phenotypes.

## Results

### Phenotype-cell type associations

In this study we systematically investigated the cell types underlying phenotypes across the HPO. We hypothesised that genes which are specifically expressed in certain cell types will be most relevant for the proper functioning of those cell types. Thus, phenotypes caused by disruptions to specific genes will have greater or lesser effects across different cell types. To test this, we computed associations between the weighted gene lists for each phenotype with the gene expression specificity for each cell type in our transcriptomic reference atlases.

More precisely, for each phenotype we created a list of associated genes weighted by the strength of the evidence supporting those associations, imported from the Gene Curation Coalition (GenCC)^38^. Analogously, we created gene expression profiles for each cell type in our scRNA-seq atlases and then applied normalisation to compute how specific the expression of each gene is to each cell type. To assess consistency in the phenotype-cell type associations, we used multiple scRNA-seq atlases: Descartes Human (∼4 million single-nuclei and single-cells from 15 fetal tissues)^32^ and Human Cell Landscape (∼703,000 single-cells from 49 embryonic, fetal and adult tissues)^33^. We ran a series of linear regression models to test for the relationship between every unique combination of phenotype and cell type. We applied multiple testing correction to control the false discovery rate (FDR) across all tests.

Within the results using the Descartes Human single-cell atlas, 19,929/ 848,078 (2.35%) tests across 77/ 77 (100%) cell types and 7,340/11,047 (66.4%) phenotypes revealed significant phenotype-cell type associations after multiple-testing correction (FDR<0.05). Using the Human Cell Landscape single-cell atlas, 26,585/1,358,916 (1.96%) tests across 124/124 (100%) cell types and 9,049/11,047 (81.9%) phenotypes showed significant phenotype-cell type associations (FDR<0.05). The median number of significantly associated phenotypes per cell type was 252 (Descartes Human) and 200 (Human Cell Landscape), respectively. Overall, using the Human Cell Landscape reference yielded a greater percentage of phenotypes with at least one significant cell type association than the Descartes Human reference. This is expected at the Human Cell Landscape contains a greater diversity of cell types across multiple life stages (embryonic, fetal, adult).

Across both single-cell references, the median number of significantly associated cell types per phenotype was 3, suggesting reasonable specificity of the testing strategy. Within the HPO, 8,628/8,631 (∼100%) of diseases gene annotations showed significant cell type associations for at least one of their respective phenotypes. A summary of the phenome-wide results stratified by single-cell atlas can be found in Table 2.

### Validation of expected phenotype-cell type relationships

We intuitively expect that abnormalities of an organ system will often be driven by cell types within that system. The HPO has broad categories at the higher level of the ontology, enabling us to systematically test this. For example, phenotypes associated with the heart should generally be caused by cell types of the heart (i.e. cardiocytes), while abnormalities of the nervous system should largely be caused by neural cells. There will of course be exceptions to this. For example, some immune disorders can cause intellectual disability through neurodegeneration. Nevertheless, it is reasonable to expect that abnormalities of the nervous system will be most often associated with neural cells. All cell types in our single-cell reference atlases were mapped onto the Cell Ontology (CL); a controlled vocabulary of cell types organised into hierarchical branches (e.g. neural cell include neurons and glia, which in turn include their respective subtypes).

Here, we consider a cell type to be *on-target* relative to a given HPO branch if it belongs to one of the matched CL branches (see Table 4). Within each high-level branch in the HPO shown in Fig. 2b, we tested whether each cell type was more often associated with phenotypes in that branch relative to those in all other branches (including those not shown). We then checked whether each cell type was overrepresented (at FDR<0.05) within its respective on-target HPO branch, where the number of phenotypes within that branch. Indeed, we found that all 7 HPO branches were disproportionately associated with on-target cell types from their respective organ systems.

**Figure 2.**
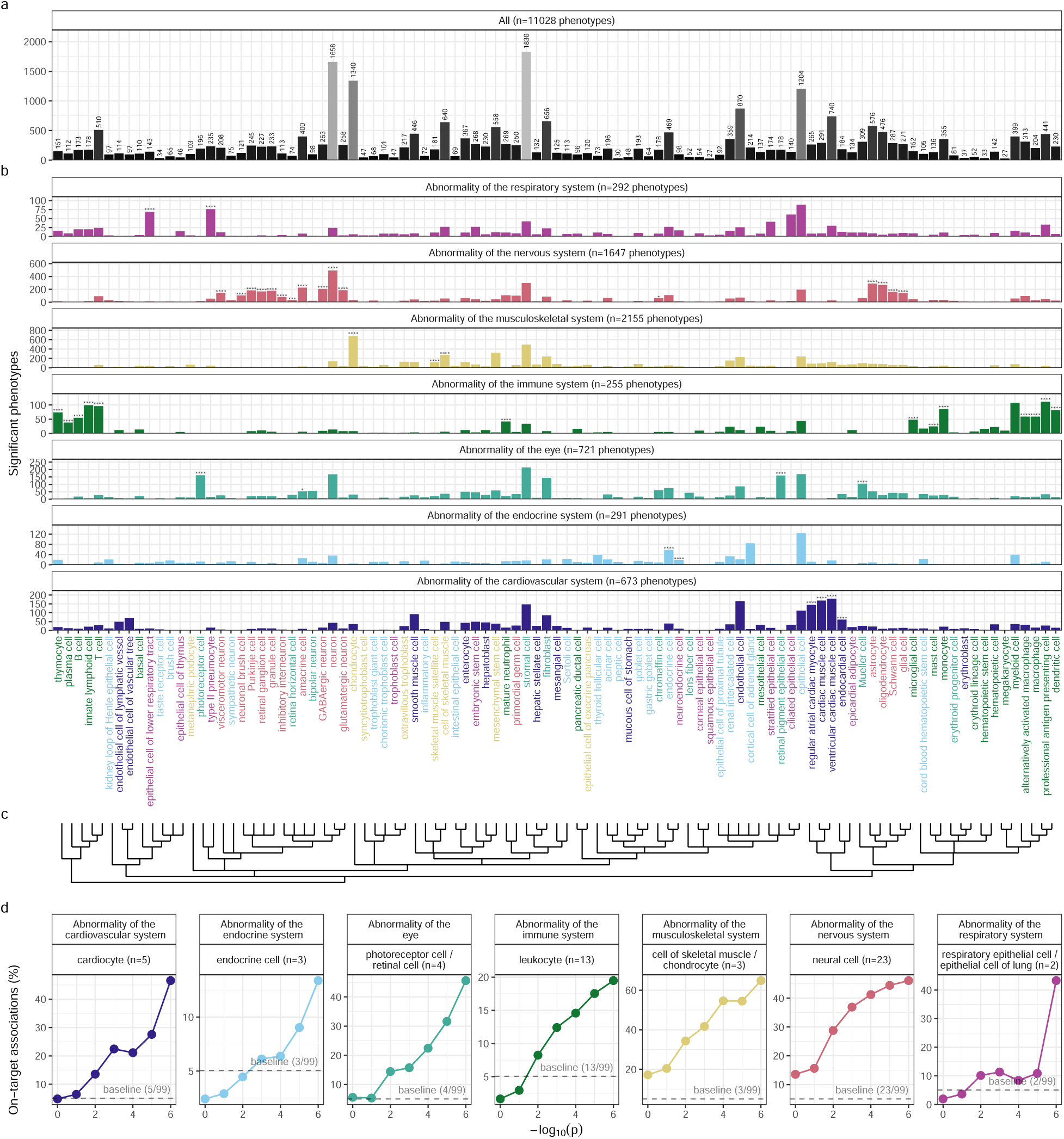
(a) High-throughput analysis reveals cell types underlying thousands of rare disease phenotypes. **a**, Some cell types are much more commonly associated with phenotypes than others. Bar height indicates the total number of significant phenotype enrichments per cell type (FDR<0.05) across all branches of the HPO. **b**, Analyses reveal expected and novel cell type associations within high-level HPO branches. Asterisks above each bar indicate whether that cell type was significantly more often enriched in that branch relative to all other HPO branches, including those not shown here, as a proxy for how specifically that cell type is associated with that branch; FDR<0.0001 (****), FDR<0.001 (***), FDR<0.01 (**), FDR<0.05 (*). **c**, Ontological relatedness of cell types in the Cell Ontology (CL)^39^. **d**, The proportion of on-target associations (*y-axis*) increases with greater test significance (*x-axis*). Percentage of significant phenotype associations with on-target cell types (second row of facet labels), respective to the HPO branch.

In addition to binary metrics of a cell type being associated with a phenotype or not, we also used association test p-values as a proxy for the strength of the association. We hypothesized that the more significant the association between a phenotype and a cell type, the more likely it is that the cell type is on-target for its respective HPO branch. To evaluate whether this, we grouped the association −*log*_10_(p-values) into 6 bins. For each HPO-CL branch pairing, we then calculated the proportion of on-target cell types within each bin. We found that the proportion of on-target cell types increased with increasing significance of the association (*rho* =0.63, *p* =1.1 × 10^−6^). For example, abnormalities of the nervous system with −*log*_10_(p-values) = 1, only 16% of the associated cell types were neural cells. Whereas for those with −*log*_10_(p-values) = 6, 46% were neural cells despite the fact that this class of cell types only constituted 23% of the total cell types tested (i.e. the baseline). This shows that the more significant the association, the more likely it is that the cell type is on-target.

### Validation of inter- and intra-dataset consistency

If our methodology works, it should yield consistent phenotype-cell type associations across different datasets. We therefore tested for the consistency of our results across the two single-cell reference datasets (Descartes Human vs. Human Cell Landscape) across the subset of overlapping cell types Fig. 11. In total there were 142,285 phenotype-cell type associations to compare across the two datasets (across 10,945 phenotypes and 13 cell types annotated to the exact same CL term. We found that the correlation between p-values of the two datasets was high (*rho*=0.91, *p*=5.7 × 10^−6^). Within the subset of results that were significant in both single-cell datasets (FDR<0.05), we found that degree of correlation between the association effect sizes across datasets was even stronger (*rho* =0.82, *p* =5.7 × 10^−6^). We also checked for the intra-dataset consistency between the p-values of the foetal and adult samples in the Human Cell Landscape, showing a very similar degree of correlation as the inter-dataset comparison (*rho* =0.95, *p* =5.0 × 10^−15^). Together, these results suggest that our approach to identifying phenotype-cell type associations is highly replicable and generalisable to new datasets.

### More specific phenotypes are associated with fewer genes and cell types

Higher levels of the ontology are broad classes of phenotype (e.g. ‘Abnormality of the nervous system’) while the lower levels can get very detailed (e.g. ‘Spinocerebellar atrophy’). The higher level phenotypes inherit all genes associated with lower level phenotypes, so naturally they have more genes than the lower level phenotypes (Fig. 3a; *rho* =−0.56, *p* =2.2 × 10^−308^).

**Figure 3.**
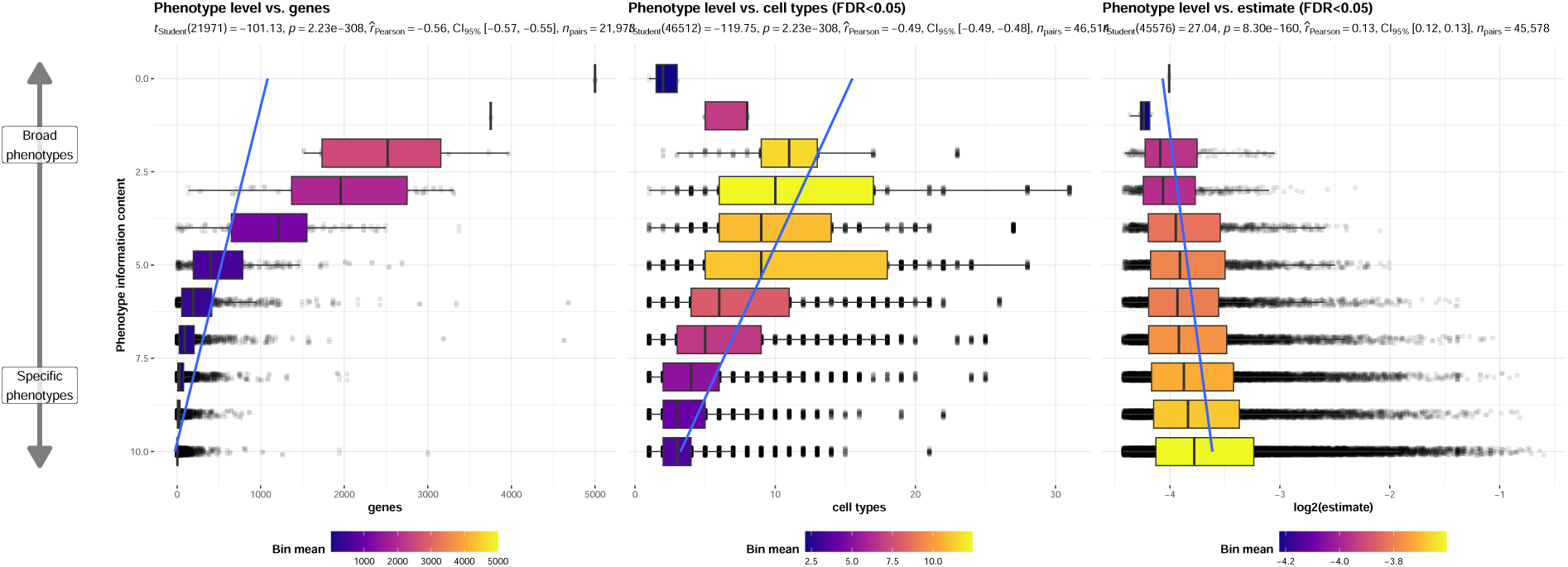
(a) More specific phenotypes are associated with fewer, more specific genes and cell types. Information content (IC), is a normalised measure of ontology term specificity. Terms with lower IC represent the broadest HPO terms (e.g. ‘All’), while terms with higher IC indicate progressively more specific HPO terms (e.g. ‘Contracture of proximal interphalangeal joints of 2nd-5th fingers’). Box plots show the relationship between HPO phenotype IC and **a**, the number of genes annotated to each phenotype, **b**, the number of significantly enriched cell types, **c**, the effect sizes (absolute model *R*^2^ estimates after log-transformation) of significant phenotype-cell type association tests. Boxes are coloured by the mean value within each IC bin (after rounding continuous IC values to the nearest integer).

Next, we reasoned that the more detailed and specific a phenotype is, the more likely it is to be driven by one cell type. For example, while ‘Neurodevelopmental abnormality’ could plausibly be driven by any/all cell types in the brain, it is more likely that ‘Impaired visuospatial constructive cognition’ is driven by fewer cell types. This was indeed the case, as we observed a strongly significant negative correlation between the two variables (Fig. 3b; *rho* =−0.49, *p* =2.2 ×10^−308^). We also found that the phenotype-cell type association effect size increased with greater phenotype specificity, reflecting the decreasing overall number of associated cell types at each ontological level (Fig. 3c; *rho* =0.13, *p* =8.3 × 10^−160^).

### Validation of phenotype-cell type associations using biomedical knowledge graphs

In order to validate our phenotype-cell type associations without the bias introduced by manually searching literature that affirmed our discoveries, we use formalised biomedical knowledge from the scientific community stored in a knowledge graph. In particular, the Monarch Knowledge Graph (MKG) is a comprehensive, standardised database that aggregates up-to-date knowledge about biomedical concepts and the relationships between them. This currently includes 103 well-established phenotype-cell type relationships^40^. We used the MKG as a proxy for the field’s current state of knowledge of causal phenotype-cell type associations. We evaluated the proportion of MKG associations that were recapitulated by our results Fig. 12. For each phenotype-cell type association in the MKG, we computed the percent of cell types recovered in our association results at a given ontological distance according to the CL ontology. An ontological distance of 0 means that our nominated cell type was as close as possible to the MKG cell type after adjusting for the cell types available in our single-cell references. Instances of exact overlap of terms between the MKG and our results would qualify as an ontological distance of 0 (e.g. ‘monocyte’ vs. ‘monocyte’). Greater ontological distances indicate further divergence between the MKG cell type and our nominated cell type. A distance of 1 indicating that the MKG cell type was one step away from our nominated cell type in the CL ontology graph (e.g. ‘monocyte’ vs. ‘classical monocyte’). The maximum possible percent of recovered terms is capped by the percentage of MKG ground-truth phenotypes we were able to find at least one significant cell type association for at *FDR*_*pc*_.

In total, our results contained at least one significant cell type associations for 90% of the phenotypes described in the MKG. Of these phenotypes, we captured 57% of the MKG phenotype-cell associations at an ontological distance of 0 (i.e. the closest possible Cell Ontology term match). Recall increased with greater flexibility in the matching of cell type annotations. At an ontological distance of 1 (e.g. ‘monocyte’ vs. ‘classical monocyte’), we captured 77% of the MKG phenotype-cell associations. Recall reached a maximum of 90% at a ontological distance of 5. This recall percentage is capped by the proportion of phenotypes for which we were able to find at least one significant cell type association for. It should be noted that we were unable to compute precision as the MKG (and other knowledge databases) only provide true positive associations. Identifying true negatives (e.g. a cell type is definitely never associated with a phenotype) is a fundamentally more difficult task to resolve as it would require proving the null hypothesis. Regardless, these benchmarking tests suggests that our results are able to recover the majority of known phenotype-cell type associations while proposing many new associations.

### Phenome-wide analyses discover novel phenotype-cell type associations

Having established that many of the phenotype-cell type associations align with prior expectations, we then sought to discover novel relationships with undercharacterised phenotypes. We reasoned that recurrent bacterial infections (and all its descendant phenotypes) should primarily be associated with immune cell types. The HPO term ‘Recurrent bacterial infections’ has 19 different descendant phenotypes, e.g. staphylococcal, streptococcal, and Neisserial infections. Each of these phenotypes are associated with partially overlapping subsets of immune cells and other cell types (Fig. 4). As expected, these phenotypes are primarily associated with immune cell types (e.g. macrophages, dendritic cells, T cells, monocytes, neutrophils). Some associations confirm relationships previously suggested in the literature, such as that between ‘Recurrent staphylococcal infections’ and myeloid cells^41–44^. Specifically, our results pinpoint monocytes as the most strongly associated cell subtypes (FDR=1.0 × 10^−30^, *β*=0.18).

**Figure 4.**
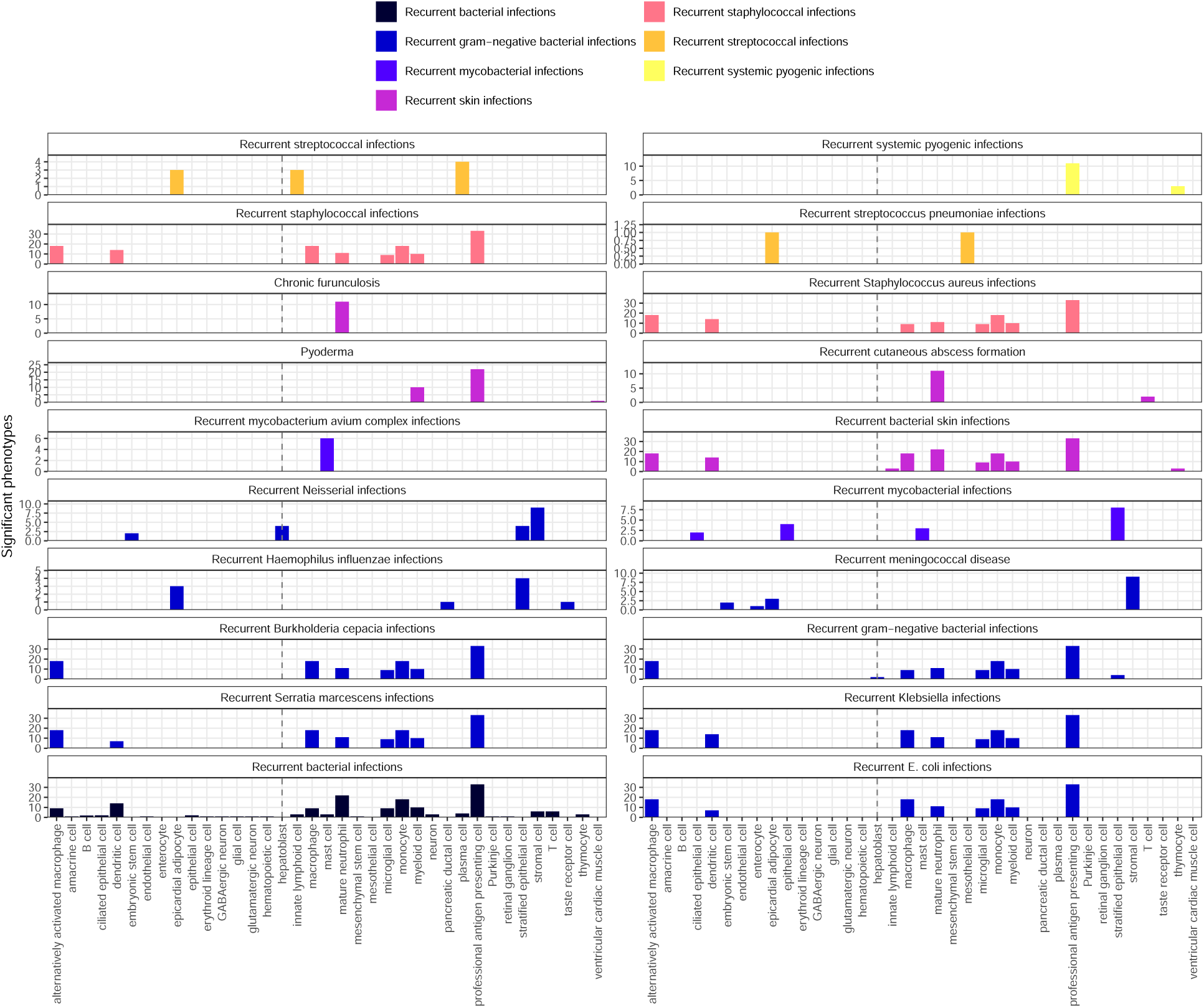
(a) Association tests reveal that hepatoblasts have a unique role in recurrent Neisserial infections. Significant phenotype-cell type tests for phenotypes within the branch ‘Recurrent bacterial infections’. Amongst all different kinds of recurrent bacterial infections, hepatoblasts (highlighted by vertical dotted lines) are exclusively enriched in ‘Recurrent gram−negative bacterial infections’. Note that terms from multiple levels of the same ontology branch are shown as separate facets (e.g. ‘Recurrent bacterial infections’ and ‘Recurrent gram−negative bacterial infections’).

Next, we sought to uncover novel, unexpected associations between recurrent bacterial infection phenotypes and cell types. In contrast to all other recurrent infection types, ‘Recurrent Neisserial infections’ highlighted a novel association with hepatoblasts (Descartes Human : FDR=1.1 × 10^−6^, *β*=8.2 × 10^−2^). Whilst unexpected, a convincing explanation involves the complement system, a key driver of innate immune response to Neisserial infections. Hepatocytes, which derive from hepatoblasts, produce the majority of complement proteins^45^, and Kupffer cells express complement receptors^46^. In addition, individuals with deficits in complement are at high risk for Neisserial infections^47,48^, and a genome-wide association study in those with a Neisserial infection identified risk variants within complement proteins^49^. While the potential of therapeutically targeting complement in RDs (including Neisserial infections) has been proposed previously^50,51^, performing this in a gene- and cell type-specific manner may help to improve efficacy and reduce toxicity (e.g. due to off-target effects). Importantly, there are over 56 known genes within the complement system^52^, highlighting the need for a systematic, evidence-based approach to identify effective gene targets.

Also of note, despite the fact that our datasets contain both hepatoblasts and their mature counterpart, hepatocytes, only the hepatoblasts showed this association. This suggests that the genetic factors that predispose individuals for risk of Neisserial infections are specifically affecting hepatoblasts before they become fully differentiated. It is also notable that these phenotypes were the only ones within the ‘Recurrent bacterial infections’ branch, or even the broader ‘Recurrent infections’ branch, perhaps indicating a unique role for hepatoblasts in recurrent infectious disease. The only phenotypes within the even broader ‘Abnormality of the immune system’ HPO branch that significantly associated with mature hepatocytes were ‘Pancreatitis’ (FDR=2.1 × 10^−2^, *β*=5.3 × 10^−2^) and ‘Susceptibility to chickenpox’ (FDR=1.2 × 10^−2^, *β*=5.5 × 10^−2^) both of which are well-known to involve the liver^53–55^.

Phenotypes can be associated with multiple diseases, cell types and genes. In addition to hepatoblasts, ‘Recurrent Neisserial infections’ were also associated with stromal cells (FDR=4.6 × 10^−6^, *β*=7.9 × 10^−2^), stratified epithelial cells (FDR=1.7 × 10^−23^, *β*=0.15), and embryonic stem cells (FDR=5.4 × 10^−5^, *β*=7.4 × 10^−2^). ‘Recurrent Neisserial infections’ is a phenotype of 7 different diseases (‘C5 deficiency’, ‘C6 deficiency’, ‘C7 deficiency’, ‘Complement component 8 deficiency, type II’, ‘Complement factor B deficiency’, ‘Complement factor I deficiency’, ‘Mannose-Binding lectin deficiency’). The monogenic nature of these diseases makes it very difficult to statistically infer the cell types underlying them. By aggregating these genes to the level of phenotype (the observed symptom) we can better understand the cell types underlying all of these diseases.

Having found four distinct cell types associated with RNI, we asked whether the RNI-associated genes were equally expressed across all of these cell types, or whether they differentially contributed to each of the associations. RNI provides a convenient case study to investigate this because each of the seven diseases that have RNI as a phenotype are purely monogenic. This makes is relatively straightforward to demonstrate how genes can drive associations between cell types, phenotypes and their respective diseases.

Diseases that have ‘Recurrent Neisserial infections’ as a phenotype were collected from the HPO annotation files. Genes that were annotated to a given phenotype (e.g. ‘Recurrent Neisserial infections’) via a particular disease (e.g. ‘C5 deficiency’) constituted “symptom”-level gene sets. Only diseases whose symptom-level gene sets had >25% overlap with the driver gene sets for at least one cell type were retained in the network plot. Using this approach, we were able to construct and refine causal networks tracing multiple scales of disease biology.

This procedure revealed that genetic deficiencies in various complement system genes (e.g. *C5*, *C8*, and *C7*) are primarily mediated by different cell types (hepatoblasts, stratified epithelial cells, and stromal cells, respectively). While genes of the complement system are expressed throughout many different tissues and cell types, these results indicate that different subsets of these genes may mediate their effects through different cell types. While almost all of these genes show high expression specificity in hepatoblasts, only *C6*, *C7* and *CFI* meet the threshold for the status of driver genes in stromal cells.

Recall that we showed in Fig. 4b that as we approach the leaf nodes of the HPO we tends towards a given phenotype being associated with a single cell type. Note that mean this in a theoretical sense, as we do not necessarily demonstrate a single cell type for each phenotype in this particular dataset. However, as more granular phenotypes are defined over time, we would expect this hypothesis to bear out. The corollary of this is that we would expect there to be at least four subtypes of the RNI phenotype, as predicted by the four distinct cell types found to underlying this phenotype. This may present as different clinical courses (e.g. early onset, late onset, relapse-remitting) or biomarkers (e.g. histological) to be reveal in future examinations of clinical cohorts. Based on this, we predict that forms of RNI caused by genes expressed in stromal cells would have phenotypic differences from those caused by genes expressed in stratified epithelial cell. In other words, phenotypic similarity is driven by the underlying causal cell types.

### Prioritising phenotypes based on severity

Some phenotypes are more severe than others and thus could be given priority for developing treatments. For example, ‘Leukonychia’ (white nails) is much less severe than ‘Leukodystrophy’ (white matter degeneration in the brain). Given the large number of significant phenotype-cell type associations, we needed a way of prioritising phenotypes for further investigation. We therefore used the large language model GPT-4 to systematically annotate the severity of all HPO phenotypes^37^.

Severity annotations were gathered from GPT-4 for 16,982/18,082 (94%) HPO phenotypes in our companion study^37^. Benchmarking tests of these results using ground-truth HPO branch annotations. For example, phenotypes within the ‘Blindness’ HPO branch (*HP:0000618*) were correctly annotated as causing blindness by GPT-4. Across all annotations, the recall rate of GPT-4 annotations was 96% (min=89%, max=100%, SD=4.5) with a mean consistency score of 91% (min=81%, max=97%, SD=5.7) for phenotypes whose annotation were collected more than once. This clearly demonstrates the ability of GPT-4 to accurately annotate phenotypes. This allowed us to begin using these annotations to compute systematically collected severity scores for all phenotypes in the HPO.

From these annotations we computed a weighted severity score metric for each phenotype ranging from 0-100 (100 being the theoretical maximum severity of a phenotype that always causes every annotation). Within our annotations, the most severe phenotype was ‘Atrophy/Degeneration affecting the central nervous system’ (*HP:0007367*) with a severity score of 47, followed by ‘Anencephaly’ (*HP:0002323*) with a severity score of 45. There were 677 phenotypes with a severity score of 0 (e.g. ‘Thin toenail’). The mean severity score across all phenotypes was 10 (median=9.4, standard deviation=6.4).

We next sought to answer the question “are disruptions to certain cell types more likely to cause severe phenotypes?”. To address this, we merged the GPT annotations with the significant (FDR<0.05) phenotype-cell type association results and computed the frequency of each severity annotation per cell type (Fig. Figure 13). We found that neuronal brush cells were associated with phenotypes that had the highest average composite severity scores, followed by Mueller cells and glial cells. This suggests that disruptions to these cell types are more likely to cause generally severe phenotypes. Meanwhile, megakaryocytes were associated with phenotypes that had the lowest average composite severity scores, suggesting that disruptions to these cell types can be better tolerated than others.

Different aspects of phenotype severity will be more associated with some cell types than others. After encoding the GPT annotations numerically (0=“never”, 1=“rarely”, 2=“often”, 3=“always”) we computed the mean encoded value per cell type within each annotation. We then ran a series of one-sided Wilcoxon rank-sum tests to objectively determine whether some cell types tended to be associated with phenotypes that more frequently caused certain severity metrics (death, intellectual disability, impaired mobility, etc.) relative to all other cell types (Fig. 5a). This consistently yielded expected relationships between cell types (e.g. retinal pigment epithelial cells) and phenotype characteristics (e.g. blindness). Similarly, phenotypes that more commonly cause death are most commonly associated with ventricular cardiac muscle cells, and least commonly associated with squamous epithelial cells and bipolar neurons. Analagous patterns of expected associations are shown consistently across all annotations (e.g. fertility-reducing phenotypes associated with cortical cell of adrenal glands, immunodeficiency-causing phenotypes associated with T cells, mobility-impairing phenotypes associated with chondrocytes, cancer-causing phenotypes associated with T cells, etc.).

**Figure 5.**
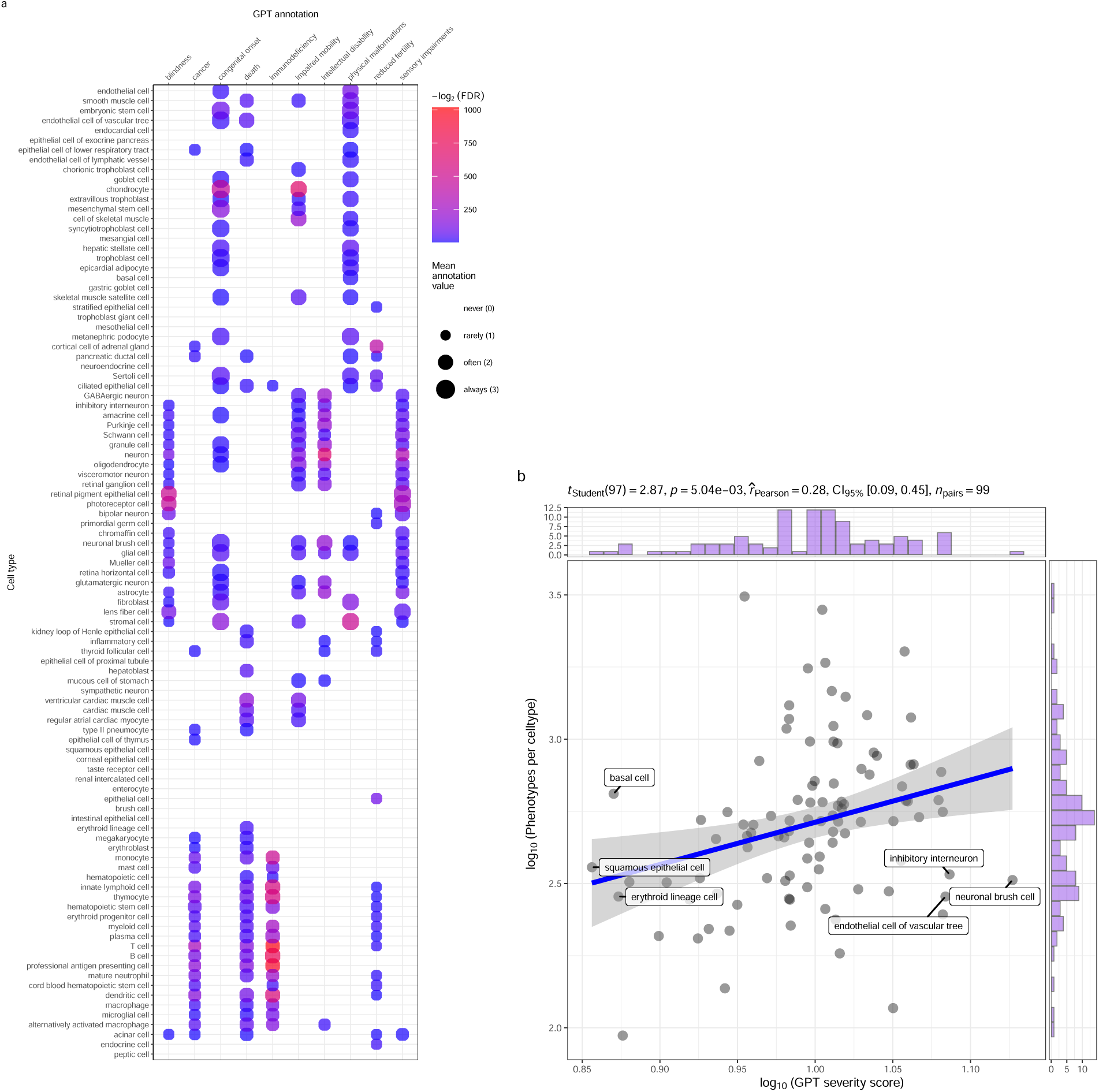
(a) Genetic disruptions to some cell types cause more clinically severe phenotypes than others. **a**, Different cell types are associated with different aspects of phenotypic severity. The dot plot shows the mean encoded frequency value for a given severity annotation (0=“never”, 1=“rarely”, 2=“often”, 3=“always”; shown as dot size), aggregated by the associated cell type. One-sided Wilcoxon rank-sum tests were performed for each cell type (within each GPT annotation) to determine which cell types more frequently caused severe phenotypes than all other cell types. Dots are colored by −*log*_2_(*FDR*) when Wilcoxon test FDR values were less than 0.05. All dots with non-significant Wilcoxon tests are instead colored grey. Cell types (rows) are clustered according to the p-values of the Wilcoxon tests. **b**, Cell types that affect more phenotypes tend to have more clinically severe consequences. Specifically, the number of phenotypes each cell type is significantly associated with, and the mean composite severity score of each cell type. The cell types with the top/bottom three x/y axis values are labeled to illustrate the cell types that cause the most/least phenotypic disruption when dysfunctional. Side histograms show the density of data points along each axis. Summary statistics for the linear regression are shown in the title (*t*_*Student*_ = Student t-test statistic, *p* = p-value, *r*_*Pearson*_ = Pearson correlation coefficient, *CI*_95%_ = confidence intervals, *n*_*Pairs*_= number of observed data pairs).

We also sought to answer whether the number of phenotypes that a cell type is associated with has a relationship with the severity of those phenotypes (Fig. 5b). Our working hypothesis is that when a cell type that affect many different phenotypes is disrupted, the cell type likely performs some critical function that affect many physiological systems. It also means that the individual phenotypes tend to be more severe than other phenotypes that involve less critical cell types. Indeed, we found a significant relationship between number of associated and mean composite phenotype severity (p=5.0 × 10^−3^, Pearson coefficient=0.28).

### Congenital phenotypes are associated with foetal cell types

Which life stage a phenotype affects an individual is clinically important and can have profound implications for how patients are treated and whether that are treatable with currently available interventions. For example, beyond a certain point gene therapies may not be an effective means of treating morphological defects that arise during development. Within the DescartesHuman dataset, 100% of the cells were from foetal tissues. Meanwhile, the Human Cell Landscape was derived from embryonic, foetal, and adult tissue samples. Within the Human Cell Landscape, 29% of cell types were found in foetal tissue, and 71% were found in adult tissues. Many of the cell types in our datasets have both foetal and adult versions (e.g. chondrocytes), while some only exist in the course of foetal development (e.g. neural crest cells). This presents a unique opportunity to provide an additional layer of contextualisation in our phenotype-cell type association results that may provide critical information when determining viable patient treatment options.

We reasoned that phenotypes that are most frequently congenital are more likely to be associated with foetal cell types than adult cell types. As expected, the frequency of congenital onset with each phenotype (as determined by GPT-4 annotations) was strongly predictive of the proportion of significantly associated foetal cell types in our results (*p* =4.7 × 10^−261^, 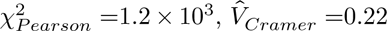, Fig. 6a). This result is consistent with the expected role of foetal cell types in development and the aetiology of congenital disorders.

**Figure 6.**
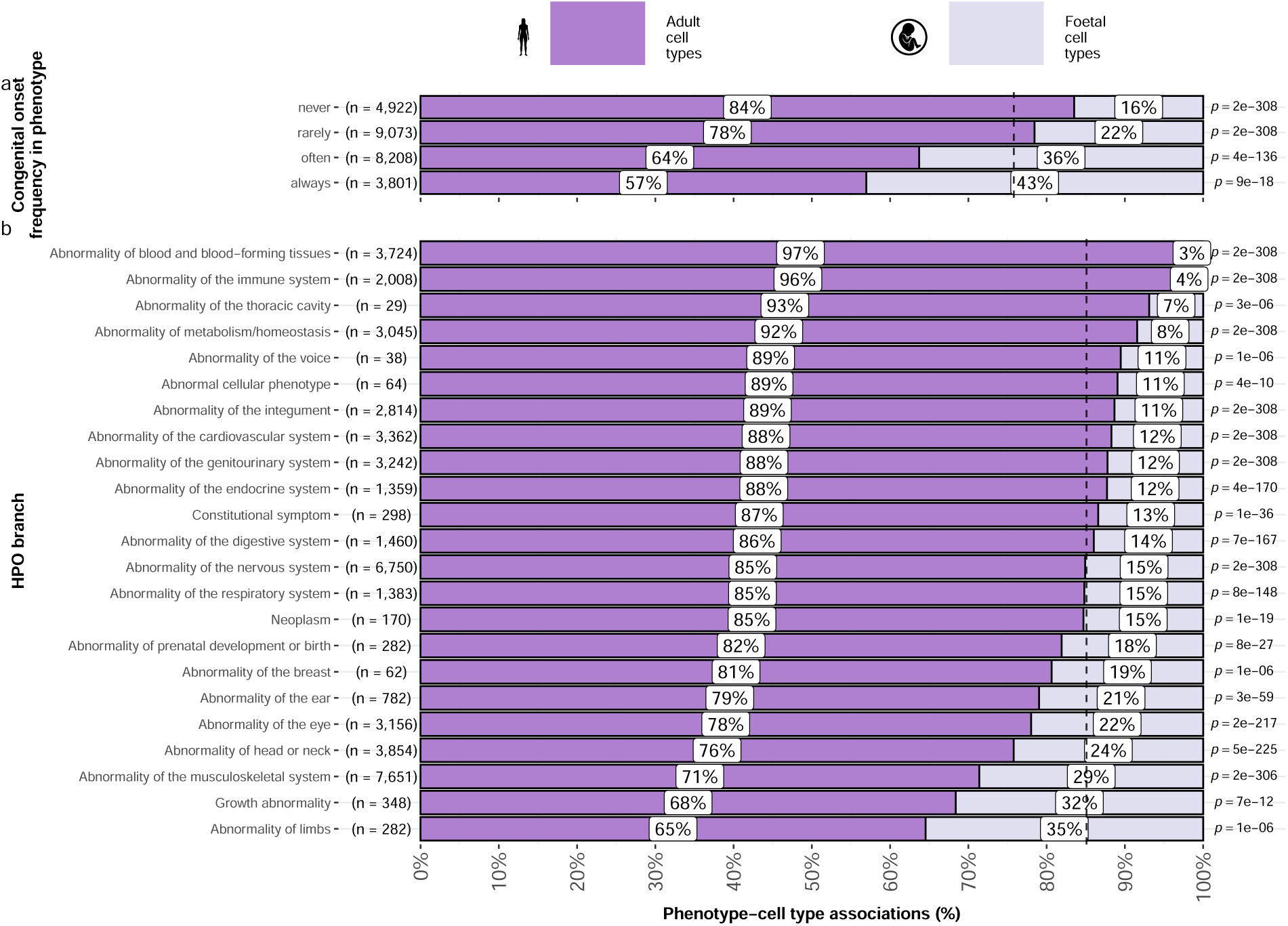
(a) Foetal vs. adult cell type references provide development context to phenotype aetiology. **a**, Congenital phenotypes are more often associated with foetal cell types. As a phenotype is more often congenital in nature, the greater proportion of foetal cell types are significantly associated with it. **b**, The proportion of phenotype-cell type association tests that are enriched for foetal cell types within each HPO branch. The p-values to the right of each bar are the results of an additional series of *χ*^2^ tests to determine whether the proportion of foetal vs. non-foetal cell types significantly different differs from the proportions expected by chance (the dashed vertical line). The feotal silhouette was generated with DALL-E. The adult silhouette is from phylopic.org and is freely available via CC0 1.0 Universal Public Domain Dedication.

Some branches of the HPO were more commonly enriched in foetal cell types compared to others (*V*_*Cramer*_=0.22, *p*<2.2 × 10^−308^, Fig. 6b). The branch with the greatest proportion of foetal cell type enrichments was ‘Abnormality of limbs’ (35%), followed by ‘Growth abnormality’ (32%) and ‘Abnormality of the musculoskeletal system’ (29%). Notably, ‘Abnormality of limbs’ branch was most disproportionatltey enriched for foetal cell type associations relative to all other branches (35% cell types). These results align well with the fact that physical malformations tend to be developmental in origin.

Conversely, the HPO branches that were most biased towards adult cell types were ‘Abnormality of blood and blood-forming tissues’ (97%), ‘Abnormality of the immune system’ (96%), and ‘Abnormality of the thoracic cavity’ (93%).

Some phenotypes exclusively involve the foetal version of a cell type, while others exclusively involve the adult version. We sought to find those phenotypes which had the greatest bias towards either end of this spectrum. To do so, we designed a metric to identify which phenotypes were more often associated with foetal cell types than adult cell types. For each phenotype, we calculated the difference in the association p-values between the foetal and adult version of the equivalent cell type. The resulting metric ranges from 1 (indicating the phenotype is only associated with the foetal version of the cell type) and −1 (indicating the phenotype is only associated with the adult version of the cell type). To summarise the most foetal-biased phenotype categories, we ran an ontological enrichment test with the HPO graph Table 7. To identify foetal cell type-biased phenotype categories, we fed the top 50 phenotypes with the greatest foetal cell type bias (closer to 1) into the enrichment function Table 8. Conversely, we used the top 50 phenotypes with the greatest adult cell type bias (closer to −1) to identify adult cell type-biased phenotype categories.

The phenotype categories with the greatest bias towards foetal cell types were ‘Abnormal nasal morphology’ (p=2.4 × 10^−7^, *log*_2_(fold-change)=4.5) and ‘Abnormal external nose morphology’ (p=2.5 × 10^−6^, *log*_2_(fold-change)=5.4).

Specific examples of such phenotypes include ‘Short middle phalanx of the 2nd finger’, ‘Abnormal morphology of the nasal alae’, and ‘Abnormal labia minora morphology’. Indeed, these phenotypes are morphological defects apparent at birth caused by abnormal developmental processes.

Conversely, the most adult cell type-biased phenotype categories were ‘Abnormal elasticity of skin’ (p=3.6 × 10^−7^, *log*_2_(fold-change)=6.0) and ‘Abnormally lax or hyperextensible skin’ (p=1.3 × 10^−5^,*log*_2_(fold-change)=6.0).

Specific examples of such phenotypes include ‘Excessive wrinkled skin’ and ‘Paroxysmal supraventricular tachycardia’ Table 8. It is well known that ageing naturally causes a loss of skin elasticity (due to decreasing collagen production) and vascular degeneration^56^. Next, we were interested whether some cell types tend to show strong differences in their phenotype associations between their foetal and adult forms. To test this, we performed an analogous enrichment procedure as with the phenotypes, except using Cell Ontology terms and the Cell Ontology graph. This analysis identified the cell type category connective tissue cell (p=1.8 × 10^−3^, *log*_2_(fold-change)=3.2) as the most foetal-biased cell type. No cell type categories were significantly enriched for the most adult-biased cell types. This is likely due to the fact that cell types can be disrupted at different stages of life, resulting in different phenotypes. Thus there the same cell types may be involved in both the most foetal-biased and adult-biased phenotypes. Together, these findings serve to further validate our methodology as a tool for identifying the causal cell types underlying a wide range of phenotypes.

### Therapeutic target identification

In the above sections, we demonstrated how gene association databases can be used to investigate the cell types underlying disease phenotypes at scale. While these associations are informative on their own, we wished to take these results further in order to have a more translational impact. Knowledge of the causal cell types underlying each phenotype can be incredibly informative for scientists and clinicians in their quest to study and treat them. Therapeutic targets with supportive genetic evidence have 2.6x higher success rates in clinical trials^57–59^. Furthermore, knowing which cell types to target with gene therapy can maximise the efficacy of highly expensive payloads, and minimise side effects (e.g. immune reaction to viral vectors). Recent biotechnological advances have greatly enhanced our ability to target specific cell types with gene therapy, making specific and accurate knowledge the correct underlying cell types more pertinent than ever^35,36^.

However, given the sheer number of results, we wished to develop a principled and reproducible approach to filter and rank putative cell type-specific gene targets for diseases where there is the greatest urgent need for improved treatments. We therefore systematically identified putative cell type-specific gene targets for severe phenotypes. First, we transformed our phenotype-cell type association results and merged them with primary data sources (e.g. GenCC gene-disease relationships, scRNA-seq atlas datasets) to create a large table of multi-scale relationships, where each row represented a tetrad of disease-phenotype-cell type-gene relationships. We then filtered non-significant phenotype-cell type relationships (only associations with *FDR* < 0.05) as well as phenotype-gene relationships with strong causal evidence (GenCC score > 3). We also removed any phenotypes that were too broad to be clinically useful, as quantified using the information content (IC) (*IC* > 8), which measures the how specific each term is within an ontology (i.e. HPO). Gene-cell type relationships were established by taking genes that had the top 25% expression specificity quantiles within each cell type. When connecting cell types to diseases via phenotypes, we used a symptom intersection threshold of >.25. Next, we sorted the remaining results in descending order of phenotype severity using the GPT4 composite severity scores described earlier. Finally, to limit the size of the resulting multi-scale networks we took only the top 10 rows, where each row represented a tetrad of disease-phenotype-cell type-gene relationships. This resulted in number of relatively small, high-confidence disease-phenotype-cell type-gene networks that could be reasonably interrogated through manual inspection and network visualisation. For example, if one was interested in the mechanisms causing ‘Recurrent Neisserial infections’, one would need only select all rows that include this phenotype to find all of its most relevant connection to diseases, cell types, and genes.

This yielded putative therapeutic targets for 5,252 phenotypes across 4,819 diseases in 201 cell types and 3,148 genes (Fig. 15). While this constitutes a large number of genes in total, each phenotype was assigned a median of 2.0 gene targets (mean=3.3, min=1, max=10). Relative to the number of genes annotations per phenotype in the HPO overall (median=7.0, mean=62, min=1, max=5,003) this represents a substantial decrease in the number of candidate target genes, even when excluding high-level phenotypes (HPO level>3.0). It is also important to note that the phenotypes in the prioritised targets list are ranked by their severity, allowing us to distinguish between phenotypes with a high medical urgency (e.g. ‘Hydranencephaly’) from those with lower medical urgency (e.g. ‘Increased mean corpuscular volume’). This can be useful for clinicians, biomedical scientists, and pharmaceutical manufacturers who wish to focus their research efforts on phenotypes with the greatest need for intervention.

Across all phenotypes, epithelial cell were most commonly implicated (838 phenotypes), followed by stromal cell (626 phenotypes), stromal cell (626 phenotypes), neuron (475 phenotypes), chondrocyte (383 phenotypes), and endothelial cell (361 phenotypes). Grouped by higher-order ontology category, ‘Abnormality of the musculoskeletal system’ had the greatest number of enriched phenotypes (959 phenotypes, 857 genes), followed by ‘Abnormality of the nervous system’ (733 phenotypes, 1,138 genes), ‘Abnormality of head or neck’ (543 phenotypes, 986 genes), ‘Abnormality of the genitourinary system’ (443 phenotypes, 695 genes), and ‘Abnormality of the eye’ (377 phenotypes, 545 genes).

### Therapeutic target validation

To determine whether the genes prioritised by our therapeutic targets pipeline were plausible, we checked what percentage of gene therapy targets we recapitulated. Data on therapeutic approval status was gathered from the Therapeutic Target Database (TTD; release 2025-08-08)^60^. Overall, we prioritised 87% (120 total) of all non-failed existing gene therapy targets (ie. those which are currently approved, investigative, or undergoing clinical trials). A hypergeometric test confirmed that our prioritised targets were significantly enriched for non-failed gene therapy targets (*p* =1.8 × 10^−5^). For these hypergeometric tests, the background gene set was composed of the union of all phenotype-associated genes in the HPO and all gene therapy targets listed in TTD.

Even when considering therapeutics of any kind (Fig. 16), not just gene therapies, we recapitulated 40% of the non-failed therapeutic targets and 0% of the terminated/withdrawn therapeutic targets (n=1,255). Here we found that our prioritised targets were highly significantly depleted for failed therapeutics (*p* =2.2 × 10^−142^). This suggests that our multi-scale evidence-based prioritisation pipeline is capable of selectively identifying genes that are likely to be effective therapeutic targets.

In addition to aggregate enrichment results, we also provide specific examples of successful gene therapies whose cell type-specific mechanism were recapitulated by our phenotype-cell associations. In particular, our pipeline nominated the gene *RPE65* within ‘retinal pigment epithelial cells’ as the top target for ‘Fundus atrophy’ vision-related phenotypes that are hallmarks of ‘Leber congenital amaurosis, type II’ and ‘Severe early-childhood-onset retinal dystrophy’. Indeed, gene therapies targeting *RPE65* within the retina of patients with these rare genetic conditions are some of the most successful clinical applications of this technology to date, able to restore vision in many cases^61^. In other cases, a tissue (e.g. liver) may be known to be causally involved in disease genesis, but the precise causal cell types within that tissue remain unknown (e.g. heptocytes, Kupffer cells, Cholangiocytes, Hepatic stellate cells, Natural killer cells, etc.). Tissue-level investigations (e.g. using bulk transcriptomics or epigenomics) would be dominated by hepatocytes, which comprise 75% of the liver. Our prioritized gene therapy targets can aid in such scenarios by providing the cell type-resolution context most likely to be causal for a given phenotype or set of phenotypes.

From our prioritised targets, we selected four phenotype or disease examples: ‘GM2-ganglioside accumulation’, ‘Spinocerebellar atrophy’, ‘Neuronal loss in central nervous system’. To focus on clinically relevant phenotypes and reduce overplotting, we limited selection to those with GPT severity scores above 15 Fig. 8a-h. Selection was based on severity and network simplicity to allow compact visualisation.

Tay-Sachs disease (TSD) is a fatal neurodegenerative condition caused by *HEXA* deficiency and ganglioside buildup. We identified alternatively activated macrophages as the cell type most associated with ‘GM2-ganglioside accumulation’ Fig. 18. This aligns with prior findings of ganglioside accumulation in TSD macrophages^62,63.,64,65^. Our results support macrophages as causal in TSD and the most promising therapeutic target.

Spinocerebellar atrophy is a progressive neurodegenerative phenotype in disorders like Spinocerebellar ataxia. Our pipeline implicates M2 macrophages (‘Alternatively activated macrophages’) as the only causal cell type Fig. 18. This suggests Purkinje cell loss is downstream of macrophage dysfunction, consistent with microglial roles in neurodegeneration^66–68^. Our findings provide the first statistically supported link between risk genes and this cell type, which is supported by relevant mouse models (e.g. *Atxn1*, *Pnpla6*) that replicate cellular and behavioural disease phenotypes.

Despite its broad definition, ‘Neuronal loss in central nervous system’ was associated with only 3 cell types: alternatively activated macrophage, macrophage, epithelial cell, specifically M2 macrophages and sinusoidal endothelial cells Fig. 18.

Skeletal dysplasia comprises 450+ disorders affecting bone and cartilage, often leading to lethal outcomes via organ compression. While surgeries offer partial relief, pharmacological options remain limited. Our analysis identified chondrocytes as causal Fig. 19, consistent with known gene–cell links (e.g. *SLC26A2*, *COL2A1* in Achondrogenesis Type 1B and Torrance-type dysplasia). Chondrocyte-targeted therapy may offer long-term solutions where surgery falls short.

Alzheimer’s disease (AD), a common neurodegenerative condition, presents with variable symptoms such as memory loss and proteinopathy. Our analysis shows distinct monogenic AD subtypes associate with different cell types and phenotypes Fig. 19. For example, AD subtypes 3 and 4 implicate digestive cells (‘enterocyte’, ‘gastric goblet cell’), while AD subtype 2 involves immune cells (‘alternatively activated macrophage’). These findings may explain heterogeneity in AD onset and presentation.

Parkinson’s disease (PD) includes motor and systemic symptoms. PD subtypes 19a and 8 implicate oligodendrocytes and neurons Fig. 19, suggesting *LRRK2* variants act via gliosis in the substantia nigra. Other PD mechanisms involved chondrocytes (PD 20), amacrine cells (late-onset PD), and respiratory/immune cells (PD 14). This diversity may underlie PD’s multisystem features.

### Experimental model translatability

We computed interspecies translatability scores using a combination of both ontological (*SIM*_*o*_) and genotypic (*SIM*_*g*_) similarity relative to each homologous human phenotype and its associated genes Fig. 17. In total, we mapped 1,221 non-human phenotypes (in *Caenorhabditis elegans*, *Danio rerio*, *Mus musculus*, *Rattus norvegicus*) to 3,319 homologous human phenotypes. Amongst the 5,252 phenotype within our prioritised therapy targets, 1,788 had viable animal models in at least on non-human species. Per species, the number of homologous phenotypes was: *Mus musculus* (n=1705)*Danio rerio* (n=244)*Rattus norvegicus* (n=85)*Caenorhabditis elegans* (n=23). Amongst our prioritised targets with a GPT-4 severity score of >10, the phenotypes with the greatest animal model similarity were “Rudimentary to absent tibiae” (*SIM*_*og*_ = 1), “Hypoglutaminemia” (*SIM*_*og*_ = 1), “Bilateral ulnar hypoplasia” (*SIM*_*og*_ = 0.99), “Disproportionate shortening of the tibia” (*SIM*_*og*_ = 0.99), “Acrobrachycephaly” (*SIM*_*og*_ = 0.98).

### Mappings

Mappings from HPO phenotypes and other commonly used medical ontologies were gathered in order to facilitate use of the results in this study in both clinical and research settings. Direct mappings, with a cross-ontology distance of 1, are the most precise and reliable. Counts of mappings at each distance are shown in Table 1. In total, there were 15,105 direct mappings between the HPO and other ontologies, with the largest number of mappings coming from the UMLS ontology (12,898 UMLS terms).

The mappings files can be accessed with the function HPOExplorer::get_mappings or directly via the HPOExplorer Releases page on GitHub (https://github.com/neurogenomics/HPOExplorer/releases/tag/latest).

## Discussion

Investigating RDs at the level of phenotypes offers numerous advantages in both research and clinical medicine. First, the vast majority of RDs only have one associated gene (7,671/8,631 diseases = 89%). Aggregating gene sets across diseases into phenotype-centric “buckets” permits sufficiently well-powered analyses, with an average of ∼76 genes per phenotype (median=7) see Fig. 10. Second, we hypothesised that these phenotype-level gene sets converge on a limited number of molecular and cellular pathways. Perturbations to these pathways manifest as one or more phenotypes which, when considered together, tend to be clinically diagnosed as a certain disease. Third, RDs are often highly heterogeneous in their clinical presentation across individuals, leading to the creation of an ever increasing number of disease subtypes (some of which only have a single documented case). In contrast, a phenotype-centric approach enables us to more accurately describe a particular individual’s version of a disease without relying on the generation of additional disease subcategories. By characterising an individual’s precise phenotypes over time, we may better understand the underlying biological mechanisms that have caused their condition. However, in order to achieve a truly precision-based approach to clinical care, we must first characterise the molecular and cellular mechanisms that cause the emergence of each phenotype. Here, we provide a highly reproducible framework that enables this at the scale of the entire phenome.

Across the 201 cell types and 11,047 RD-associated phenotypes investigated, more than 46,514 significant phenotype-cell type relationships were discovered. This presents a wealth of opportunities to trace the mechanisms of rare diseases through multiple biological scales. This in turn enhances our ability to study and treat causal factors in disease with deeper understanding and greater precision. These results recapitulate well-known relationships, while providing additional cellular context to many of these known relationships, and discovering novel relationships.

It was paramount to the success of this study to ensure our results were anchored in ground-truth benchmarks, generated falsifiable hypotheses, and rigorously guarded against false-positive associations. Extensive validation using multiple approaches demonstrated that our methodology consistently recapitulates expected phenotype-cell type associations (Fig. 2-Fig. 6). This was made possible by the existence of comprehensive, structured ontologies for all phenotypes (the Human Phenotype Ontology) and cell types (the Cell Ontology), which provide an abundance of clear and falsifiable hypotheses for which to test our predictions against. Several key examples include 1) strong enrichment of associations between cell types and phenotypes within the same anatomical systems (Fig. 2b-d), 2) a strong relationship between phenotype-specificity and the strength and number of cell type associations (Fig. 3), 3) identification of the precise cell subtypes involved in susceptibility to various subtypes of recurrent bacterial infections (Fig. 4), 4) a strong positive correlation between the frequency of congenital onset of a phenotype and the proportion of developmental cell types associated with it (Fig. 6)), and 5) consistent phenotype-cell type associations across multiple independent single-cell datasets (Fig. 11).

Unfortunately, there are currently only treatments available for less than 5% of RDs^6^. Novel technologies including CRISPR, prime editing, antisense oligonucleotides, viral vectors, and/or lipid nanoparticles, have been undergone significant advances in the last several years^69–73^ and proven remarkable clinical success in an increasing number of clinical applications^74–77^. The U.S. Food and Drug Administration (FDA) recently announced an landmark program aimed towards improving the international regulatory framework to take advantage of the evolving gene/cell therapy technologies^78^ with the aim of bringing dozens more therapies to patients in a substantially shorter timeframe than traditional pharmaceutical product development (typically 5-20 years with a median of 8.3 years)^79^. While these technologies have the potential to revolutionise RD medicine, their successful application is dependent on first understanding the mechanisms causing each disease.

To address this critical gap in knowledge, we used our results to create a reproducible and customisable pipeline to nominate cell type-resolved therapeutic targets (Fig. 15-Fig. 8). Targeting cell type-specific mechanisms underlying granular RD phenotypes can improve therapeutic effectiveness by treating the causal root of an individual’s conditions^70,80^. A cell type-specific approach also helps to reduce the number of harmful side effects caused by unintentionally delivering the therapeutic to off-target tissues/cell types (which may induce aberrant gene activity), especially when combined with technologies that can target cell surface antigens (e.g viral vectors)^81^. This has the additional benefit of reducing the minimal effective dose of a therapeutic, which can be both immunogenic and extremely financially costly^9,10,69,72^. Here, we demonstrate the utility of a high-throughput evidence-based approach to RD therapeutics discovery by highlighting several of the most promising therapeutic candidates. Our pipeline takes into account a myriad of factors, including the strength of the phenotype-cell type associations, symptom-cell type associations, cell type-specificity of causal genes, the severity and frequency of the phenotypes, suitability for gene therapy delivery systems (e.g. recombinant adeno-associated viral vectors (rAAV)), as well as a quantitative analysis of phenotypic and genetic animal model translatability (Fig. 17). We validated these candidates by comparing the proportional overlap with gene therapies that are presently in the market or undergoing clinical trials, in which we recovered 87% of all active gene therapies (Fig. 7, Fig. 16). Despite nominating a large number of putative targets, hypergeometric tests confirmed that our targets were strongly enriched for targets of existing therapies that are either approved or currently undergoing clinical trials.

**Figure 7.**
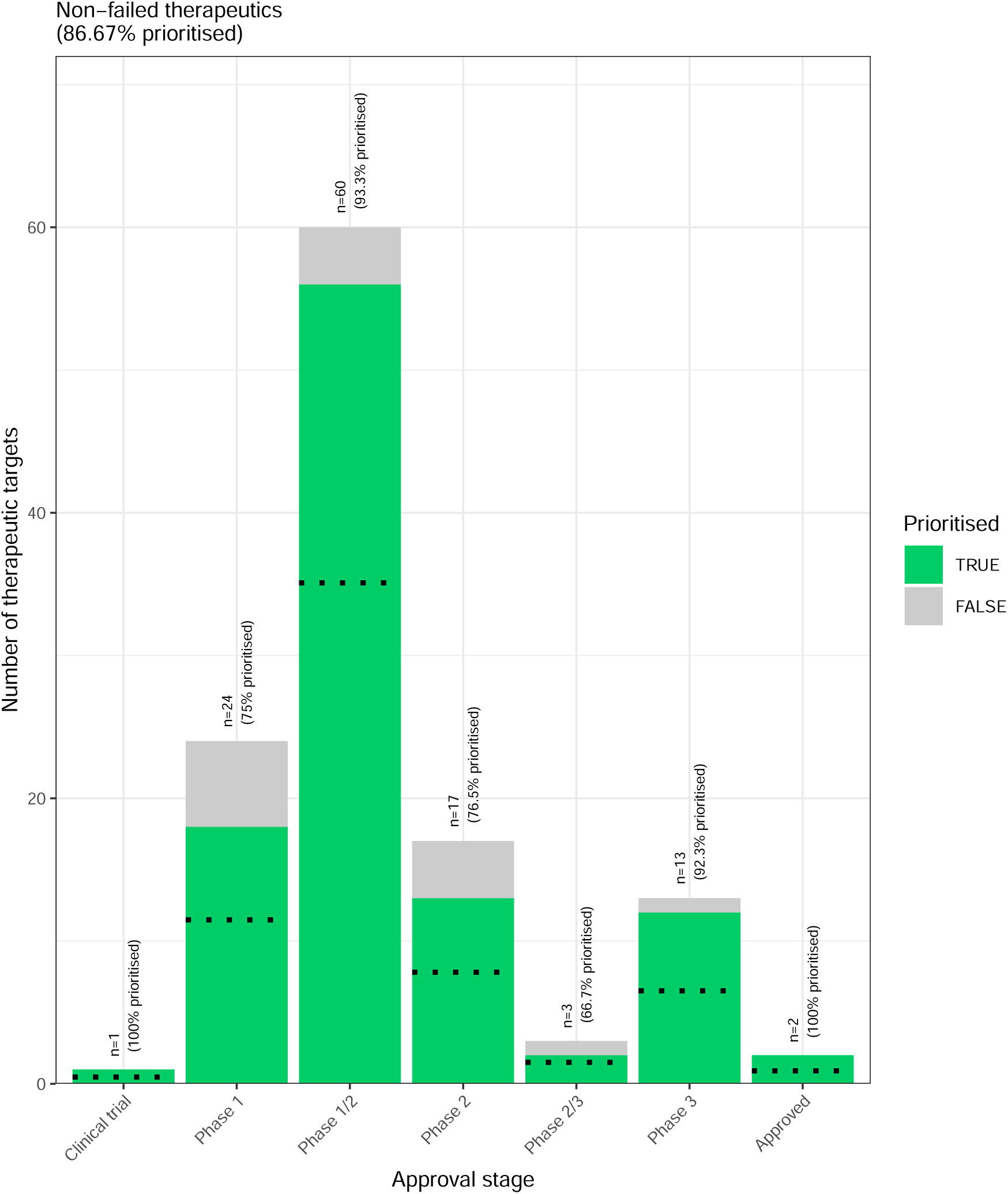
(a) Prioritised targets recapitulate existing gene therapy targets. The proportion of existing gene therapy targets (documented in the Therapeutic Target Database) recapitulated by our prioritisation pipeline. Therapeutics are stratified by the stage of clinical development they were at during the time of writing. While our prioritized targets did not include any failed (‘Terminated’) therapies, the fact that only one such therapy exists in the dataset preclude us from making any conclusions about depletion of failed gene therapy targets in our prioritised targets list.

From our target prioritisation pipeline results, we highlight cell type-specific mechanisms for ‘GM2-ganglioside accumulation’ in Tay-Sachs disease, spinocerebellar atrophy in spinocerebellar ataxia, and ‘Neuronal loss in central nervous system’ in a variety of diseases (Fig. 8). Of interest, all three of these neurodegenerative phenotypes involved alternatively activated (M2) macrophages. The role of macrophages in neurodegeneration is complex, with both neuroprotective and neurotoxic functions, including the clearance of misfolded proteins, the regulation of the blood-brain barrier, and the modulation of the immune response^82^. We also recapitulated prior evidence that microglia, the resident macrophages of the nervous system, are causally implicated in Alzheimer’s disease (AD) (Fig. 19)^83^. An important contribution of our current study is that we were able to pinpoint the specific phenotypes of AD caused by macrophages to neurofibrillary tangles and long-tract signs (reflexes that indicate the functioning of spinal long fiber tracts). Other AD-associated phenotypes were caused by other cell types (e.g. gastric goblet cells, enterocytes).

**Figure 8.**
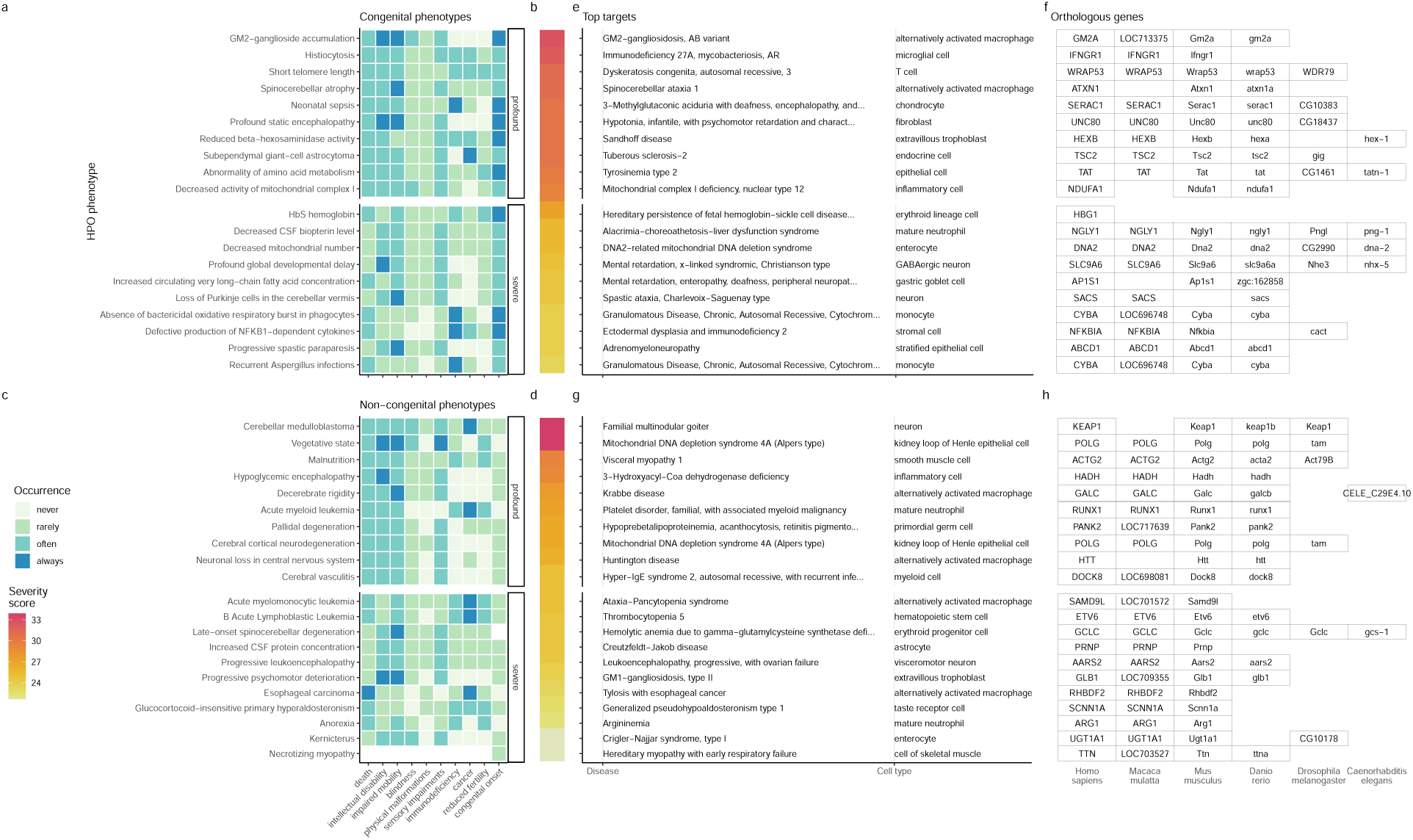
(a) Evidence-based pipeline nominates causal mechanisms to target for gene therapy. Shown here are the top 40 prioritised gene therapy targets at multiple biological scales, stratified by congenital (top row) vs. noncongential phenotypes (bottom row) as well as severity class (“profound” or “severe”). In this plot, only the top 10 most severe phenotypes within a given strata/substrata are shown **a,c**, Severity annotation generated by GPT-4. **b,d**, Composite severity scores computed across all severity metrics. **e,g**, Top mediator disease and cell type-specific target for each phenotype. **f,h** top target gene for each phenotype within humans (*Homo sapiens*). We also include the 1:1 ortholog of each human gene in several commonly used animal models, including monkey (*Macaca mulatta*), mouse (*Mus musculus*), zebrafish (*Danio rerio*), fly (*Drosophila melanogaster*) and nematode (*Caenorhabditis elegans*). Boxes are empty where no 1:1 ortholog is known. See supplement Fig. 18 for network plots of cell type-specific gene therapy targets for several severe phenotypes and their associated diseases.

It should be noted that our study has several key limitations. First, while our cell type datasets are amongst the most comprehensive human scRNA-seq references currently available, they are nevertheless missing certain tissues, cell types (e.g. spermatocytes, oocytes), and life stages (post-natal childhood, senility). It is also possible that we have not captured certain cell state signatures that only occur in disease (e.g. disease-associated microglia^84,85^). Though we reasoned that using only control cell type signatures would mitigate bias towards any particular disease, and avoid degradation of gene signatures due to loss of function mutations. Second, the collective knowledge of gene-phenotype and gene-disease associations is far from complete and we fully anticipate that these annotations will continue to expand and change well into the future. It is for this reason we designed this study to be easily reproduced within a single containerised script so that we (or others) may rerun it with updated datasets at any point. Finally, causality is notoriously difficult to prove definitively from associative testing alone, and our study is not exempt from this rule. Despite this, there are several reasons to believe that our approach is able to better approximate causal relationships than traditional approaches. First, we did not intentionally preselect any subset of phenotypes or cell types to investigate here. Along with a scaling prestep during linear modelling, this means that all the results are internally consistent and can be directly compared to one another (in stark contrast to literature meta-analyses). Furthermore, for the phenotype gene signatures we used expert-curated GenCC annotations^86,87^ to weight the current strength of evidence supporting a causal relationship between each gene and phenotype. This is especially important for phenotypes with large genes lists (thousands of annotations) for which some of the relationships may be tenuous. Within the cell type references, we deliberately chose to use specificity scores (rather than raw gene expression) as this normalisation procedure has previously been demonstrated to better distinguish between signatures of highly similar cell types/subtypes^88^.

Common ontology-controlled frameworks like the HPO open a wealth of new opportunities, especially when addressing RDs. Services such as the Matchmaker Exchange^89,90^ have enabled the discovery of hundreds of underlying genetic etiologies, and led to the diagnosis of many patients. This also opens the possibility of gathering cohorts of geographically dispersed patients to run clinical trials, the only viable option for treatment in many individuals. To further increase the number of individuals who qualify for these treatments, as well as the trial sample size, proposals have been made deviate from the traditional single-disease clinical trial model and instead perform basket trials on groups of RDs with shared molecular etiologies (SaME)^91^.

Moving forward, we are now actively seeking industry and academic partnerships to begin experimentally validating our multi-scale target predictions and exploring their potential for therapeutic translation. Nevertheless, there are more promising therapeutic targets here than our research group could ever hope to pursue by ourselves. In the interest of accelerating research and ensuring RD patients are able to benefit from this work as quickly as possible, we have decided to publicly release all of the results described in this study. These can be accessed in multiple ways, including through a suite of R packages as well as a web app, the Rare Disease Celltyping Portal (https://neurogenomics-ukdri.dsi.ic.ac.uk/). The latter allows our results to be easily queried, filtered, visualised, and downloaded without any knowledge of programming. Through these resources we aim to make our findings useful to a wide variety of RD stakeholders including subdomain experts, clinicians, advocacy groups, and patients.

## Conclusions

In this study we aimed to develop a methodology capable of generating high-throughput phenome-wide predictions while preserving the accuracy and clinical utility typically associated with more narrowly focused studies. With the rapid advancement of gene therapy technologies, and a regulatory landscape that is evolving to better meet the needs of a large and diverse patient population, there is finally momentum to begin to realise the promise of genomic medicine. This has especially important implications for the global RD community which has remained relatively neglected. Here, we have provided a scalable, cost-effective, and fully reproducible means of resolving the multi-scale, cell-type specific mechanisms of virtually all rare diseases.

## Methods

### Human Phenotype Ontology

The latest version of the HPO (release 2024-02-08) was downloaded from the EMBL-EBI Ontology Lookup Service^92^ and imported into R using the `HPOExplorer` package. This R object was used to extract ontological relationships between phenotypes as well as to assign absolute and relative ontological levels to each phenotype. The latest version of the HPO phenotype-to-gene mappings and phenotype annotations were downloaded from the official HPO GitHub repository and imported into R using `HPOExplorer`. This contains lists of genes associated with phenotypes via particular diseases, formatted as three columns in a table (gene, phenotype, disease).

However, not all genes have equally strong evidence of causality with a disease or phenotype, especially when considering that the variety of resources used to generate these annotations (OMIM, Orphanet, DECIPHER) use variable methodologies (e.g. expert-curated review of the medical literature vs. automated text mining of the literature). Therefore we imported data from the Gene Curation Coalition (GenCC)^86,87^, which (as of 2025-08-02) 24,112 evidence scores across 7,566 diseases and 5,533 genes. Evidence scores are defined by GenCC using a standardised ordinal rubric which we then encoded as a semi-quantitative score ranging from 0 (no evidence of disease-gene relationship) to 6 (strongest evidence of disease-gene relationship) (see Table 5). As each Disease-Gene pair can have multiple entries (from different studies) with different levels of evidence, we then summed evidence scores per Disease-Gene pair to generate aggregated Disease-by-Gene evidence scores. This procedure can be described as follows.

Let us denote:

- *D* as diseases.
- *P* as phenotypes in the HPO.
- *G* as genes
- *S* as the evidence scores describing the strength of the relationship between each Disease-Gene pair.
- *M*_*ij*_ as the aggregated Disease-by-Gene evidence score matrix.

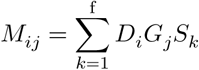

Next, we extracted Disease-Gene-Phenotype relationships from the annotations file distributed by the HPO (*phenotype_to_genes.txt*). This provides a list of genes associated with phenotypes via particular diseases, but does not include any strength of evidence scores.

Here we define: - *A*_*ijk*_ as the Disease-Gene-Phenotype relationships. - *D*_𝑖_ as the 𝑖th disease. - *G*_𝑗_ as the 𝑗th gene. - *P*_𝑘_ as the 𝑘th phenotype.

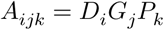

In order to assign evidence scores to each Phenotype-Gene relationship, we combined the aforementioned datasets from GenCC (*M*_*ij*_) and HPO (*A*_*ijk*_) by merging on the gene and disease ID columns. For each phenotype, we then computed the mean of Disease-Gene scores across all diseases for which that phenotype is a symptom. This resulted in a final 2D tensor of Phenotype-by-Gene evidence scores (𝐿_*ij*_):

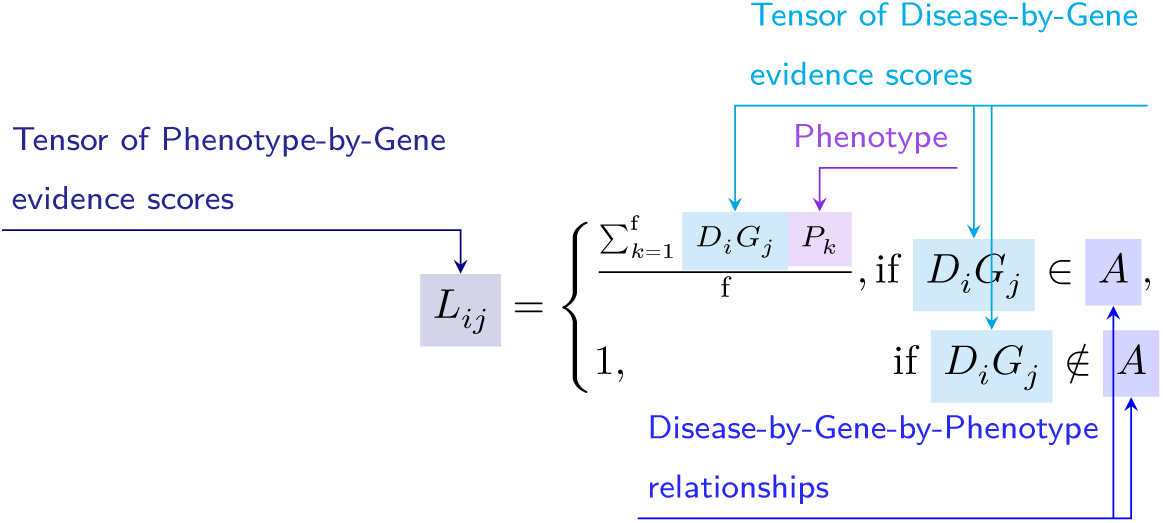

**Construction of the tensor of Phenotype-by-Gene evidence scores.**

Histograms of evidence score distributions at each step in processing can be found in Fig. 9.

### Single-cell transcriptomic atlases

In this study, the gene by cell type specificity matrix was constructed using the Descartes Human transcriptome atlas of foetal gene expression, which contains a mixture of single-nucleus and single-cell RNA-seq data (collected with sci-RNA-seq3)^32^. This dataset contains 377,456 cells representing 77 distinct cell types across 15 tissues. All 121 human foetal samples ranged from 72 to 129 days in estimated postconceptual age. To independently replicate our findings, we also used the Human Cell Landscape which contains single-cell transcriptomic data (collected with microwell-seq) from embryonic, foetal, and adult human samples across 49 tissues^33^.

Specificity matrices were generated separately for each transcriptomic atlas using the R package `EWCE` (v1.11.3)^88^. Within each atlas, cell types were defined using the authors’ original freeform annotations in order to preserve the granularity of cell subtypes as well as incorporate expert-identified rare cell types. Cell types were only aligned and aggregated to the level of corresponding Cell Ontology (CL)^39^ annotations afterwards when generating summary figures and performing cross-atlas analyses. Using the original gene-by-cell count matrices from each single-cell atlas, we computed gene-by-cell type expression specificity matrices as follows. Genes with very no expression across any cell types were considered to be uninformative and were therefore removed from the input gene-by-cell matrix 𝐹 (*g*, 𝑖, 𝑐).

Next, we calculated the mean expression per cell type and normalised the resulting matrix to transform it into a gene-by-cell type expression specificity matrix (*S*_*g*,𝑐_). In other words, each gene in each cell type had a 0-1 score where 1 indicated the gene was mostly specifically expressed in that particular cell type relative to all other cell types. This procedure was repeated separately for each of the single-cell atlases and can be summarised as:

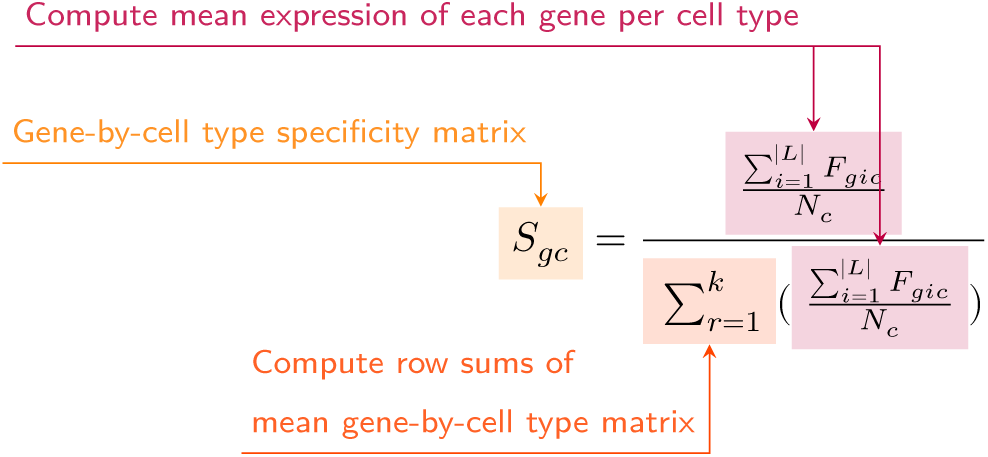

### Phenotype-cell type associations

To test for relationships between each pairwise combination of phenotype (n=11,047) and cell type (n=201) we ran a series of univariate generalised linear models implemented via the `stats::glm` function in R. First, we filtered the gene-by-phenotype evidence score matrix (𝐿_*ij*_) and the gene-by-cell type expression specificity matrix (*S*_*g*𝑐_) to only include genes present in both matrices (n=4,949 genes in the Descartes Human analyses; n=4,653 genes in the Human Cell Landscape analyses). Then, within each matrix any rows or columns with a sum of 0 were removed as these were uninformative data points that did not vary. To improve interpretability of the results *β* coefficient estimates across models (i.e. effect size), we performed a scaling prestep on all dependent and independent variables. Initial tests showed that this had virtually no impact on the total number of significant results or any of the benchmarking metrics based on p-value thresholds Fig. 2. This scaling prestep improved our ability to rank cell types by the strength of their association with a given phenotype as determined by separate linear models.

We repeated the aforementioned procedure separately for each of the single-cell references. Once all results were generated using both cell type references (2,206,994 association tests total), we applied Benjamini-Hochberg false discovery rate^93^ (denoted as *FDR*_*pc*_) to account for multiple testing. Of note, we applied this correction across all results at once (as opposed to each single-cell reference separately) to ensure the *FDR*_*pc*_ was stringently controlled for across all tests performed in this study.

### Symptom-cell type associations

Here we define a symptom as a phenotype as it presents within the context of the specific disease. The features of a given symptom can be described as the subset of genes annotated to phenotype *p* via a particular disease 𝑑, denoted as *G*_𝑑*p*_(see Fig. 10). To attribute our phenotype-level cell type enrichment signatures to specific diseases, we first identified the gene subset that was most strongly driving the phenotype-cell type association by computing the intersect of genes that were both in the phenotype annotation and within the top 25% specificity percentile for the associated cell type. We then computed the intersect between symptom genes (*G*_𝑑*p*_) and driver genes (*G*_*pc*_), resulting in the gene subset *G*_𝑑∩*p*∩𝑐_. Only *G*_𝑑∩*p*∩𝑐_gene sets with 25% or greater overlap with the symptom gene subset (*G*_𝑑*p*_) were kept. This procedure was repeated for all phenotype-cell type-disease triads, which can be summarised as follows:

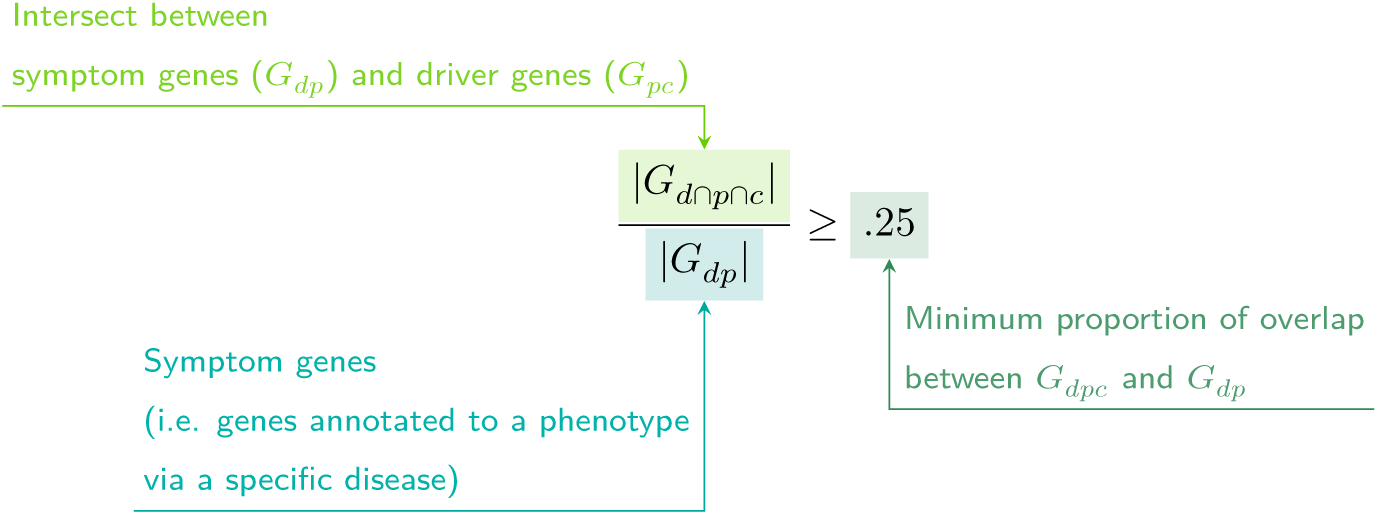

### Validation of expected phenotype-cell type relationships

We first sought to confirm that our tests (across both single-cell references) were able to recover expected phenotype-cell type relationships across seven high-level branches within the HPO (Fig. 2), including abnormalities of the cardiovascular system, endocrine system, eye, immune system, musculoskeletal system, nervous system, and respiratory system. Within each branch the number of significant tests in a given cell type were plotted (Fig. 2b). Mappings between freeform annotations (the level at which we performed our phenotype-cell type association tests) provided by the original atlas authors and their closest CL term equivalents were provided by CellxGene^30^. CL terms along the *x-axis* of Fig. 2b were assigned colours corresponding to which HPO branch showed the greatest number of enrichments (after normalising within each branch to account for differences in scale). The normalised colouring allows readers to quickly assess which HPO branch was most often associated with each cell type, while accounting for differences in the number of phenotypes across branches. We then ran a series of Analysis of Variance (ANOVA) tests to determine whether (within a given branch) a given cell type was more often enriched (FDR<0.05) within that branch relative to all of the other HPO branches of an equivalent level in the ontology (including all branches not shown in Fig. 2b). After applying Benjamini-Hochberg multiple testing correction^93^ (denoted as *FDR*_𝑏,𝑐_), we annotated each respective branch-by-cell type bar according to the significance (**** : *FDR*_𝑏,𝑐_ < 1𝑒 − 04, *** : *FDR*_𝑏,𝑐_ < 0.001, ** : *FDR*_𝑏,𝑐_ < 0.01, * : *FDR*_𝑏,𝑐_ < 0.05). Cell types in Fig. 2a-b were ordered along the *x-axis* according to a dendrogram derived from the CL ontology (Fig. 2c), which provides ground-truth semantic relationships between all cell types (e.g. different neuronal subtypes are grouped together).

As an additional measure of the accuracy of our phenotype-cell types test results we identified conceptually matched branches across the HPO and the CL (Fig. 2d and Table 6). For example, ‘Abnormality of the cardiovascular system’ in the HPO was matched with ‘cardiocytes’ in the CL which includes all cell types specific to the heart. Analogously, ‘Abnormality of the nervous system’ in the HPO was matched with ‘neural cell’ in the CL which includes all descendant subtypes of neurons and glia. This cross-ontology matching was repeated for each HPO branch and can be referred to as on-target cell types. Within each branch, the −*log*_10_(*FDR*_*pc*_) values of on-target cell types were binned by rounding to the nearest integer (*x-axis*) and the percentage of tests for on-target cell types relative to all cell types were computed at each bin (*y-axis*) (Fig. 2d). The baseline level (dotted horizontal line) illustrates the percentage of on-target cell types relative to the total number of observed cell types. Any percentages above this baseline level represent greater than chance representation of the on-target cell types in the significant tests.

### Validation of inter- and intra-dataset consistency

We tested for inter-dataset consistency of our phenotype-cell type association results across different single-cell reference datasets (Descartes Human and Human Cell Landscape). For all tests reported here, the relevant association metrics (p-values or effect size) were first averaged to the level of ancestral HPO terms (5 levels down the hierachy) to reduce figure size. For association tests with exactly matching Cell Ontology ID across the two references, we tested for a relationship between the p-values generated with each of the references by fitting linear regression model (`stats::lm` via the R function `ggstatsplot::ggscatterstats`). Next, we performed an additional linear regression between the effect sizes (each GLM model’s *R*^2^ estimates after applying a *log*_2_fold-change tranformation) of all significant phenotype-cell type associations (FDR < 0.05) with exactly matching cell types across the two references.

We also tested for intra-dataset consistency within the Human Cell Landscape by running additional linear regressions between the phenotype-cell type association test statistics of the foetal and the adult samples (using both p-values and model *R*^2^ estimates). While we would not expect the same exact cell type associations across different developmental stages, we would nevertheless expect there to be some degree of correlation between the developing and mature versions of the same cell types.

### More specific phenotypes are associated with fewer genes and cell types

To explore the relationship between HPO phenotype specificity and various metrics from our results, we computed the information content (IC) scores for each term in the HPO. IC is a measure of how much specific information a term within an ontology contains. In general, terms deeper in an ontology (closer to the leaves) are more specific, and thus informative, than terms at the very root of the ontology (e.g. ‘Phenotypic abnormality’). Where 𝑘 denotes the number of offspring terms (including the term itself) and 𝑁 denotes the total number of terms in the ontology, IC can be calculated as:

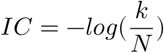

Next, IC scores were quantised into 10 bins using the `ceiling` R function to improve visualisation. We then performed a series of linear regressions between phenotype binned IC scores and: 1) number of genes annotated per HPO phenotype, 2) the number of significantly associated cell types per HPO phenotype, and 3) the model estimate of each significant phenotype-cell type associations (at FDR < 0.05) after taking the log of the absolute value (*log*_2_(|𝑒𝑠*t*𝑖𝑚𝑎*t*𝑒|)).

### Monarch Knowledge Graph recall

Finally, we gathered known phenotype-cell type relationships from the Monarch Knowledge Graph (MKG), a comprehensive database of links between many aspects of disease biology^40^. This currently includes 103 links between HPO phenotypes (n=103) and CL cell types (n=79). Of these, we only considered the 82 phenotypes that we were able to test given that our ability to generate associations was dependent on the existence of gene annotations within the HPO. We considered instances where we found a significant relationship between exactly matching pairs of HPO-CL terms as a hit.

However, as the cell types in MKG were not necessarily annotated at the same level as our single-cell references, we considered instances where the MKG cell type was an ancestor term of our cell type (e.g. ‘myeloid cell’ vs. ‘monocyte’), or *vice versa*, as hits. We also adjusted ontological distance by computing the ratio between the observed ontological distance and the smallest possible ontological distance for that cell type given the cell type that were available in our references 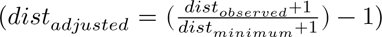. This provides a way of accurately measuring how dissimilar our identified cell types were for each phenotype-cell type association (Fig. 12).

### Prioritising phenotypes based on severity

Only a small fraction of the the phenotypes in HPO (<1%) have metadata annotations containing information on their time course, consequences, and severity. This is due to the time-consuming nature of manually annotating thousands of phenotypes. To generate such annotations at scale, we previously used Generative Pre-trained Transformer 4 (GPT-4), a large language model (LLM) as implemented within OpenAI’s Application Programming Interface (API)^37^. After extensive prompt engineering and ground-truth benchmarking, we were able to acquire annotations on how often each phenotype directly causes intellectual disability, death, impaired mobility, physical malformations, blindness, sensory impairments, immunodeficiency, cancer, reduced fertility, or is associated with a congenital onset. These criteria were previously defined in surveys of medical experts as a means of systematically assessing phenotype severity^94^. Responses for each metric were provided in a consistent one-word format which could be one of: ‘never’, ‘rarely’, ‘often’, ‘always’. This procedure was repeated in batches (to avoid exceeding token limits) until annotations were gathered for 16,982/18,082 HPO phenotypes.

We then encoded these responses into a semi-quantitative scoring system (‘never’=0, ‘rarely’=1, ‘often’=2, ‘always’=3), which were then weighted by multiplying a semi-subjective scoring of the relevance of each metric to the concept of severity on a scale from 1.0-6.0, with 6.0 being the most severe (‘death’=6, ‘intellectual_disability’=5, ‘impaired_mobility’=4, ‘physical_malformations’=3, ‘blindness’=4, ‘sensory_impairments’=3, ‘immunodeficiency’=3, ‘cancer’=3, ‘reduced_fertility’=1, ‘congenital_onset’=1). Finally, the product of the score was normalised to a quantitative severity score ranging from 0-100, where 100 is the theoretical maximum severity score. This phenotype severity scoring procedure can be expressed as follows.

Let us denote:

- *p* : a phenotype in the HPO.
- 𝑗 : the identity of a given annotation metric (i.e. clinical characteristic, such as ‘intellectual disability’ or ‘congenital onset’).
- 𝑊_𝑗_: the assigned weight of metric 𝑗.
- 𝐹_𝑗_: the maximum possible value for metric 𝑗, equal to 3 (“always”). This value is equivalent across all 𝑗 annotations.
- 𝐹_*p*𝑗_ : the numerically encoded value of annotation metric 𝑗 for phenotype *p*.
- 𝑁*SS*_*p*_: the final composite severity score for phenotype *p* after applying normalisation to align values to a 0-100 scale and ensure equivalent meaning regardless of which other phenotypes are being analysed in addition to *p*. This allows for direct comparability of severity scores across studies with different sets of phenotypes.

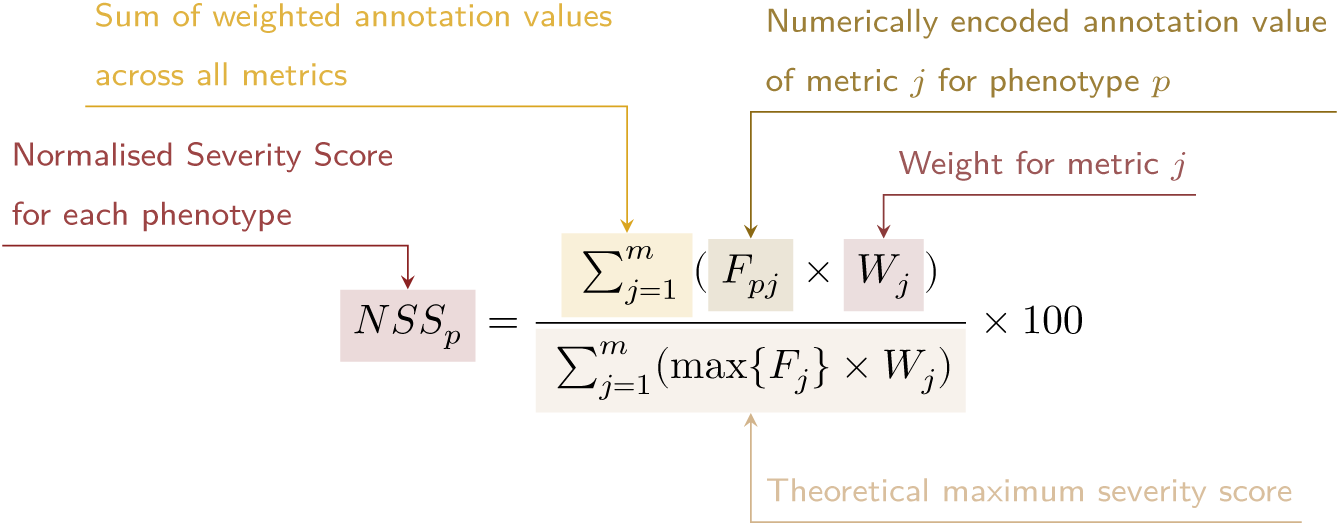

Using the numerically encoded GPT annotations (0=“never”, 1=“rarely”, 2=“often”, 3=“always”) we computed the mean encoded value per cell type within each annotation. One-sided Wilcoxon rank-sum tests were run using the `rstatix::wilcox_test()` function to test whether each cell type was associated with more severe phenotypes relative to all other cell types. This procedure was repeated for severity annotation independently (death, intellectual disability, impaired mobility, etc.) Fig. 5a. Next, we performed a Pearson correlation test between the number of phenotypes that a cell type is significantly associated with (at FDR<0.05) has a relationship with the mean composite GPT severity score of those phenotypes (Fig. 5b). This was performed using the `ggstatsplot::ggscatterstats()` R function.

### Congenital phenotypes are associated with foetal cell types

The GPT-4 annotations also enabled us to assess whether foetal cell types were more often significantly associated with congenital phenotypes in our Human Cell Landscape results as this single-cell reference contained both adult and foetal versions of cell types (Fig. 6). To do this, we performed a chi-squared (*χ*^2^) test on the proportion of significantly associated cell types containing any of the substrings ‘fetal’, ‘fetus’, ‘primordial’, ‘hESC’ or ‘embryonic’ (within cell types annotations from the original Human Cell Landscape authors^33^) vs. those associated without, stratified by how often the corresponding phenotype had a congenital onset according to the GPT phenotype annotations (including ‘never’, ‘rarely’, ‘often’, ‘always’). In addition, a series of *χ*^2^ tests were performed within each congenital onset frequency strata, to determine whether the observed proportion of foetal cell types vs. non-foetal cell types significantly deviated from the proportions expected by chance.

We next tested whether the proportion of tests with significant associations with foetal cell types varied across the major HPO branches using a *χ*^2^ test. We also performed separate *χ*^2^ test within each branch to determine whether the proportion of significant associations with foetal cell types was significantly different from chance.

Next, we aimed to create a continuous metric from −1 to 1 that indicated how biased each phenotype is towards associations with the foetal or adult form of a cell type. For each phenotype we calculated the foetal-adult bias score as the difference in the association p-values between the foetal and adult version of the equivalent cell type (foetal-adult bias ∶ *p*_𝑎𝑑𝑢𝑙*t*_ − *p*_𝑓*o*𝑒*t*𝑎𝑙_ = Δ*p* ∈ [−1, 1]). A score of 1 indicates the phenotype is only associated with the foetal version of the cell type and −1 indicates the phenotype is only associated with the adult version of the cell type.

In order to summarise higher-order HPO phenotype categories that were most biased towards foetal or adult cell types, ontological enrichment tests were run on the phenotypes with the top/bottom 50 greatest/smallest foetal-adult bias scores. The enrichment tests were performed using the `simona::dag_enrich_on_offsprings` function, which uses a hypergeometric test to determine whether a list of terms in an ontology are enriched for offspring terms (descendants) of a given ancestor term within the ontology. Phenotypes categories with an HPO ontological enrichment a p-value < 0.05 were considered significant.

We were similarly interested in which higher-order cell type categories tended to be most commonly associated with these strongly foetal-/adult-biased phenotype s. Another set of ontological enrichment tests were run on the cell types associated with the top/bottom 50 phenotypes from the previous analysis. The CL ontology-aligned IDs for each group cell types were fed into the `simona::dag_enrich_on_offsprings` using the CL ontology. Significiantly enriched cell type categories were defined as those with a CL ontological enrichment p-value < 0.05.

### Therapeutic target identification

We developed a systematic and automated strategy for identifying putative cell type-specific gene targets for each phenotype based on a series of filters at phenotype, cell type, and gene levels. The entire target prioritisation procedure can be replicated with a single function: `MSTExplorer::prioritise_targets`. This function automates all of the reference data gathering (e.g. phenotype metadata, cell type metadata, cell type signature reference, gene lengths, severity tiers) and takes a variety of arguments at each step for greater customisability. Each step is described in detail in Table 3. Phenotypes that often or always caused physical malformations (according to the GPT-4 annotations) were also removed from the final prioritised targets list, as these were unlikely to be amenable to gene therapy interventions. Finally, phenotypes were sorted by their composite severity scores such that the most severe phenotypes were ranked the highest.

### Therapeutic target validation

To assess whether our prioritised therapeutic targets were likely to be viable, we computed the overlap between our gene targets and those of existing gene therapies at various stages of clinical development (Fig. 7). Gene targets were obtained for each therapy from the Therapeutic Target Database (TTD; release 2025-08-08) and mapped onto standardised HUGO Gene Nomenclature Committee (HGNC) gene symbols using the `orthogene` R package. We stratified our overlap metrics according to whether the therapies had failed (unsuccessful clinical trials or withdrawn), or were non-failed (successful or ongoing clinical trials). We then conducted hypergeometric tests to determine whether the observed overlap between our prioritised targets and the non-failed therapy targets was significantly greater than expected by chance (i.e. enrichment). We also conducted a second hypergeometric test to determine whether the observed overlap between our prioritised targets and the failed therapy targets was significantly less than expected by chance (i.e. depletion). Finally, we repeated the analysis against all therapeutic targets, not just those of gene therapies, to determine whether our prioritised targets had relevance to other therapeutic modalities.

### Experimental model translatability

To improve the likelihood of successful translation between preclinical animal models and human patients, we created an interspecies translatability prediction tool for each phenotype nominated by our gene therapy prioritised pipeline (Fig. 17). First, we extracted ontological similarity scores of homologous phenotypes across species from the MKG^40^. Briefly, the ontological similarity scores (*SIM*_*o*_) are computed for each homologous pair of phenotypes across two ontologies by calculating the overlap in homologous phenotypes that are ancestors or descendants of the target phenotype. Next, we generated genotypic similarity scores (*SIM*_*g*_) for each homologous phenotype pair by computing the proportion of 1:1 orthologous genes using gene annotation from their respective ontologies. Interspecies orthologs were also obtained from the MKG. Finally, both scores are multiplied together to yield a unified ontological-genotypic similarity score (*SIM*_*og*_).

### Novel R packages

To facilitate all analyses described in this study and to make them more easily reproducible by others, we created several open-source R packages. `KGExplorer` imports and analyses large-scale biomedical knowledge graphs and ontologies. `HPOExplorer` aids in managing and querying the directed acyclic ontology graph within the HPO. `MSTExplorer` facilitates the efficient analysis of many thousands of phenotype-cell type association tests, and provides a suite of multi-scale therapeutic target prioritisation and visualisation functions. These R packages also include various functions for distributing the post-processed results from this study in an organised, tabular format. Of note, `MSTExplorer::load_example_results` loads all summary statistics from our phenotype-cell type tests performed here.

### Rare Disease Celltyping Portal

To further increase the ease of access for stakeholders in the RD community without the need for programmatic experience, we developed a series of web apps to interactively explore, visualise, and download the results from our study. Collectively, these web apps are called the Rare Disease Celltyping Portal. The website can be accessed at https://neurogenomics-ukdri.dsi.ic.ac.uk/.

The Rare Disease Celltyping Portal integrates diverse datasets, including the HPO, cell types, genes, and phenotype severity, into a unified platform that allows users to perform flexible, bidirectional queries. Users can start from any entry point: either phenotype, cell type, genes, or severity, and seamlessly trace relationships across these dimensions.

The portal provides a dynamic and intuitive exploration experience with its real-time interaction capabilities and responsive interface including network graphs, bar charts, and heat maps. It has the ability to handle large datasets efficiently and offer fast query response by building with FARM stack (FastAPI, React, MongoDB). The portal is designed for a broad audience, including researchers, clinicians, and biologists, by offering user-friendly navigation and interactive visual outputs. By enabling users to intuitively explore complex biological relationships, the portal aims to accelerate rare disease research, enhance diagnostic accuracy, and drive therapeutic innovation.

All code used to generate the website can be found at https://github.com/neurogenomics/Rare-Disease-Web-Portal.

### Mappings

Mappings from the HPO to other medical ontologies were extracted from the EMBL-EBI Ontology Xref Service (OxO; https://www.ebi.ac.uk/spot/oxo/) by selecting the National Cancer Institute metathesaurus (NCIm) as the target ontology and either “SNOMED CT”, “UMLS”, “ICD-9” or “ICD-10CM” as the data source. HPO terms were then selected as the ID framework with to mediate the cross-ontology mappings. Mappings between each pair of ontologies were then downloaded, stored in a tabular format, and uploaded to the public `HPOExplorer` Releases page (https://github.com/neurogenomics/HPOExplorer/releases).

## Data Availability

All data is publicly available through the following resources:

- Human Phenotype Ontology (https://hpo.jax.org)
- GenCC (https://thegencc.org/)
- Descartes Human scRNA-seq atlas (https://cellxgene.cziscience.com/collections/c114c20f-1ef4-49a5-9c2e-d965787fb90c)
- Human Cell Landscape scRNA-seq atlas (https://cellxgene.cziscience.com/collections/38833785-fac5-48fd-944a-0f62a4c23ed1)
- Processed Cell Type Datasets (*ctd_DescartesHuman.rds* and *ctd_HumanCellLandscape.rds*; https://github.com/neurogenomics/MSTExplorer/releases)
- Gene x Phenotype association matrix (*hpo_matrix.rds*; https://github.com/neurogenomics/MSTExplorer/releases)
- GPT-4 phenotype severity annotations (https://github.com/neurogenomics/rare_disease_celltyping/releases/download/latest/gpt_check_annot.csv.gz)
- Full phenotype-cell type association test results https://github.com/neurogenomics/MSTExplorer/releases/download/v0.1.10/phenomix_results.tsv.gz
- Rare Disease Celltyping Portal (https://neurogenomics-ukdri.dsi.ic.ac.uk/)

## Code Availability

All code is made freely available through the following GitHub repositories:

- `KGExplorer` (https://github.com/neurogenomics/KGExplorer)
- `HPOExplorer` (https://github.com/neurogenomics/HPOExplorer)
- `MSTExplorer` (https://github.com/neurogenomics/MSTExplorer)
- Code to replicate analyses (https://github.com/neurogenomics/rare_disease_celltyping)
- Cell type-specific gene target prioritisation (https://neurogenomics.github.io/RareDiseasePrioritisation/reports/prioritise_targets)
- Complement system gene list (https://www.genenames.org/data/genegroup/#!/group/492)

## Acknowledgements

We would like to thank the following individuals for their insightful feedback and assistance with data resources: Sarah J. Marzi, Gerton Lunter, Peter Robinson, Melissa Haendel, Ben Coleman, Nico Matentzoglu, Shawn T. O’Neil, Alan E. Murphy, Sarada Gurung.

## Funding

This work was supported by a UK Dementia Research Institute (UK DRI) Future Leaders Fellowship [MR/T04327X/1] and the UK DRI which receives its funding from UK DRI Ltd, funded by the UK Medical Research Council, Alzheimer’s Society and Alzheimer’s Research UK.

## Supplementary Materials

### Supplementary Results

#### Selected example targets

From our prioritised targets, we selected the following four sets of phenotypes or diseases as examples: ‘GM2-ganglioside accumulation’, ‘Spinocerebellar atrophy’, ‘Neuronal loss in central nervous system’. Only phenotypes with a GPT severity score greater than 15 were considered to avoid overplotting and to focus on the more clinically relevant phenotypes Fig. 8a-h. These examples were then selected partly on the basis of severity rankings, and partly for their relatively smaller, simpler networks than lent themselves to compact visualisations.

Tay-Sachs disease (TSD) is a devastating hereditary condition in which children are born appearing healthy, which gradually degrades leading to death after 3-5 years. The underlying cause is the toxic accumulation of gangliosides in the nervous system due to a loss of the enzyme produced by *HEXA*. While this could in theory be corrected with gene editing technologies, there remain some outstanding challenges. One of which is identifying which cell types should be targeted to ensure the most effective treatments. Here we identified alternatively activated macrophages as the cell type most strongly associated with ‘GM2-ganglioside accumulation’ Fig. 8i. The role of aberrant macrophage activity in the regulation of ganglioside levels is supported by observation that gangliosides accumulate within macrophages in TSD^62^, as well as experimental evidence in rodent models^63.,64,65^. Our results not only corroborate these findings, but propose macrophages as the primary causal cell type in TSD, making it the most promising cell type to target in therapies.

Another challenge in TSD is early detection and diagnosis, before irreversible damage has occurred. Our pipeline implicated extravillous trophoblasts of the placenta in ‘GM2-ganglioside accumulation’. While not necessarily a target for gene therapy (as the child is detached from the placenta after birth), checking these cells *in utero* for an absence of *HEXA* may serve as a viable biomarker as these cells normally express the gene at high levels. Early detection of TSD may lengthen the window of opportunity for therapeutic intervention^95^, especially when genetic sequencing is not available or variants of unknown significance are found within *HEXA*^96^.

Spinocerebellar atrophy is a debilitating and lethal phenotype that occurs in diseases such as Spinocerebellar ataxia and Boucher-Nenhauser syndrome. These diseases are characterised by progressive degeneration of the cerebellum and spinal cord, leading to severe motor and cognitive impairments. Our pipeline identified M2 macrophages (labeled as the closest CL term ‘Alternatively activated macrophages’ in Fig. 8j) as the only causal cell type associated with ‘Spinocerebellar atrophy’. This strongly suggests that degeneration of cerebellar Purkinje cells are in fact downstream consequences of macrophage dysfunction, rather than being the primary cause themselves. This is consistent with the known role of macrophages, especially microglia, in neuroinflammation and other neurodegenerative conditions such as Alzheimer’s and Parkinsons’ disease^66–68^. While experimental and postmortem observational studies have implicated microglia in spinocerebellar atrophy previously^66^, our results provide a statistically-supported and unbiased genetic link between known risk genes and this cell type. Therefore, targeting M2 microglia in the treatment of spinocerebellar atrophy may therefore represent a promising therapeutic strategy. This is aided by the fact that there are mouse models that perturb the ortholog of human spinocerebellar atrophy risk genes (e.g. *Atxn1*, *Pnpla6*) and reliably recapitulate the effects of this diseases at the cellular (e.g. loss of Purkinje cells), morphological (e.g. atrophy of the cerebellum, spinal cord, and muscles), and functional (e.g. ataxia) levels.

Next, we investigated the phenotype ‘Neuronal loss in the central nervous system’. Despite the fact that this is a fairly broad phenotype, we found that it was only significantly associated with 3 cell types (alternatively activated macrophage, macrophage, epithelial cell), specifically M2 macrophages and sinusoidal endothelial cells Fig. 8k.

Skeletal dysplasia is a heterogeneous group of over 450 disorders that affect the growth and development of bone and cartilage. This phenotype can be lethal when deficient bone growth leads to the constriction of vital organs such as the lungs. Even after surgical interventions, these complications continue to arise as the child develops. Pharmacological interventions to treat this condition have largely been ineffective. While there are various cell types involved in skeletal system development, our pipeline nominated chondrocytes as the causal cell type underlying the lethal form of this condition (Fig. 19). Assuringly, we found that the disease ‘Achondrogenesis Type 1B’ is caused by the genes *SLC26A2* and *COL2A1* via chondrocytes. We also found that ‘Platyspondylic lethal skeletal dysplasia, Torrance type’. Thus, in cases where surgical intervention is insufficient, targeting these genes within chondrocytes may prove a viable long-term solution for children suffering from lethal skeletal dysplasia.

Alzheimer’s disease (AD) is the most common neurodegenerative condition. It is characterised by a set of variably penetrant phenotypes including memory loss, cognitive decline, and cerebral proteinopathy. Interestingly, we found that different forms of early onset AD (which are defined by the presence of a specific disease gene) are each associated with different cell types via different phenotypes (Fig. 19). For example, AD 3 and AD 4 are primarily associated with cells of the digestive system (‘enterocyte’, ‘gastric goblet cell’) and are implied to be responsible for the phenotypes ‘Senile plaques’, ‘Alzheimer disease’, ‘Parietal hypometabolism in FDG PET’. Meanwhile, AD 2 is primarily associated with immune cells (‘alternatively activated macrophage’) and is implied to be responsible for the phenotypes ‘Neurofibrillary tangles’, ‘Long-tract signs’. This suggests that different forms of AD may be driven by different cell types and phenotypes, which may help to explain its variability in onset and clinical presentation.

Finally, Parkinson’s disease (PD) is characterised by motor symptoms such as tremor, rigidity, and bradykinesia. However there are a number of additional phenotypes associated with the disease that span multiple physiological systems. PD 19a and PD 8 seemed to align most closely with the canonical understanding of PD as a disease of the central nervous system in that they implicated oligodendrocytes and neurons (Fig. 19). Though the reference datasets being used in this study were not annotated at sufficient resolution to distinguish between different subtypes of neurons, in particular dopaminergic neurons. PD 19a/8 also suggested that risk variants in *LRRK2* mediate their effects on PD through both myeloid cells and oligodendrocytes by causing gliosis of the substantia nigra. The remaining clusters of PD mechanisms revolved around chondrocytes (PD 20), amacrine cells of the eye (hereditary late-onset PD), and the respiratory/immune system (PD 14). While the diversity in cell type-specific mechanisms is somewhat surprising, it may help to explain the wide variety of cross-system phenotypes frequently observed in PD.

It should be noted that the HPO only includes gene annotations for the monogenic forms of AD and PD. However it has previously been shown that there is at least partial overlap in their phenotypic and genetic aetiology with respect to their common forms. Thus understanding the monogenic forms of these diseases may shed light onto their more common counterparts.

### Phenome-wide analyses discover novel phenotype-cell type associations

We visualised the putative causal relationships between genes, cell types and diseases associated with RNI as a network (Fig. 14). The phenotype ‘Recurrent Neisserial infections’ was connected to cell types through the aforementioned association test results (FDR<0.05). Genes that were primarily driving these associations (i.e. genes that were both strongly linked with ‘Recurrent Neisserial infections’ and were highly specifically expressed in the given cell type) were designated as “driver genes” and retained for plotting. Across all phenotypes in the HPO, more specific phenotypes (terms in the HPO with greater IC) are not only more specific to certain cell types (Fig. 3b), but are also associated with genes that have greater cell type-specific expression within those cell types. Even so, we should note that the choice of which specificity quantiles to include is arbitrary. It should also be noted that simply because a gene is not specific to a cell type does not mean it is not important for the function of the cell type. Indeed, there are many genes that are ubiquitously expressed throughout many tissues in the body and are essential for cell function. Gene expression specificity is nevertheless a useful metric to help distinguish many hundreds of cell (sub)types with overlapping gene signatures.

### Supplementary Figures

**Figure 9.**
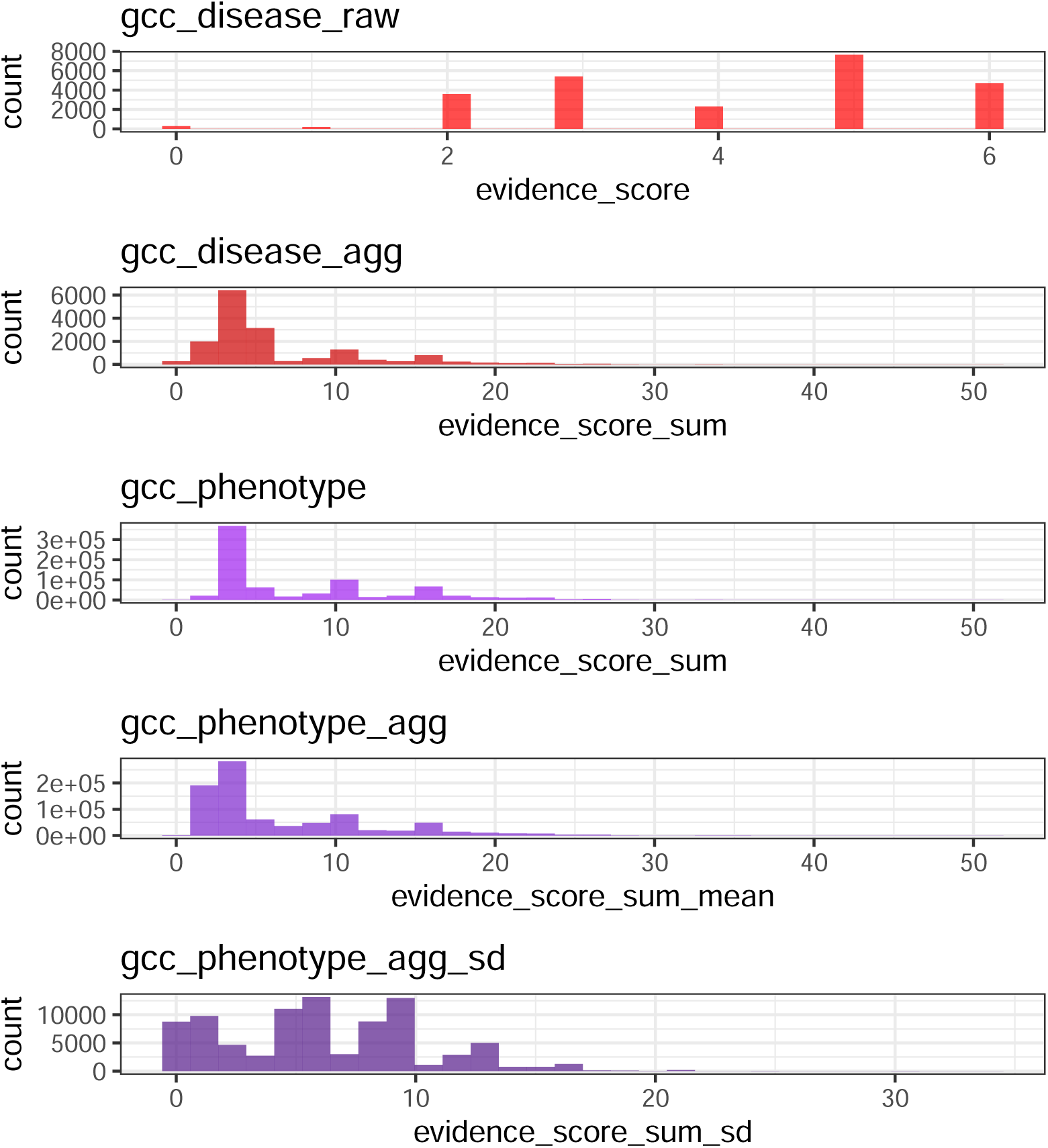
(a) Distribution of GenCC evidence scores at each processing step. GenCCC (https://thegencc.org/) is a database where semi-quantitative scores for the current strength of evidence attributing disruption of a gene as a causal factor in a given disease. “gcc_disease_raw” is the distribution of raw GenCC scores before any aggregation. “gcc_disease_agg” is the distribution of GenCC scores after aggregating by disease. “gcc_phenotype” is the distribution of scores after linking each phenotype to one or more disease. “gcc_phenotype_agg” is the distribution of scores after aggregating by phenotype, while “gcc_phenotype_agg_sd” is the standard deviation of those aggregated scores.

**Figure 10:**
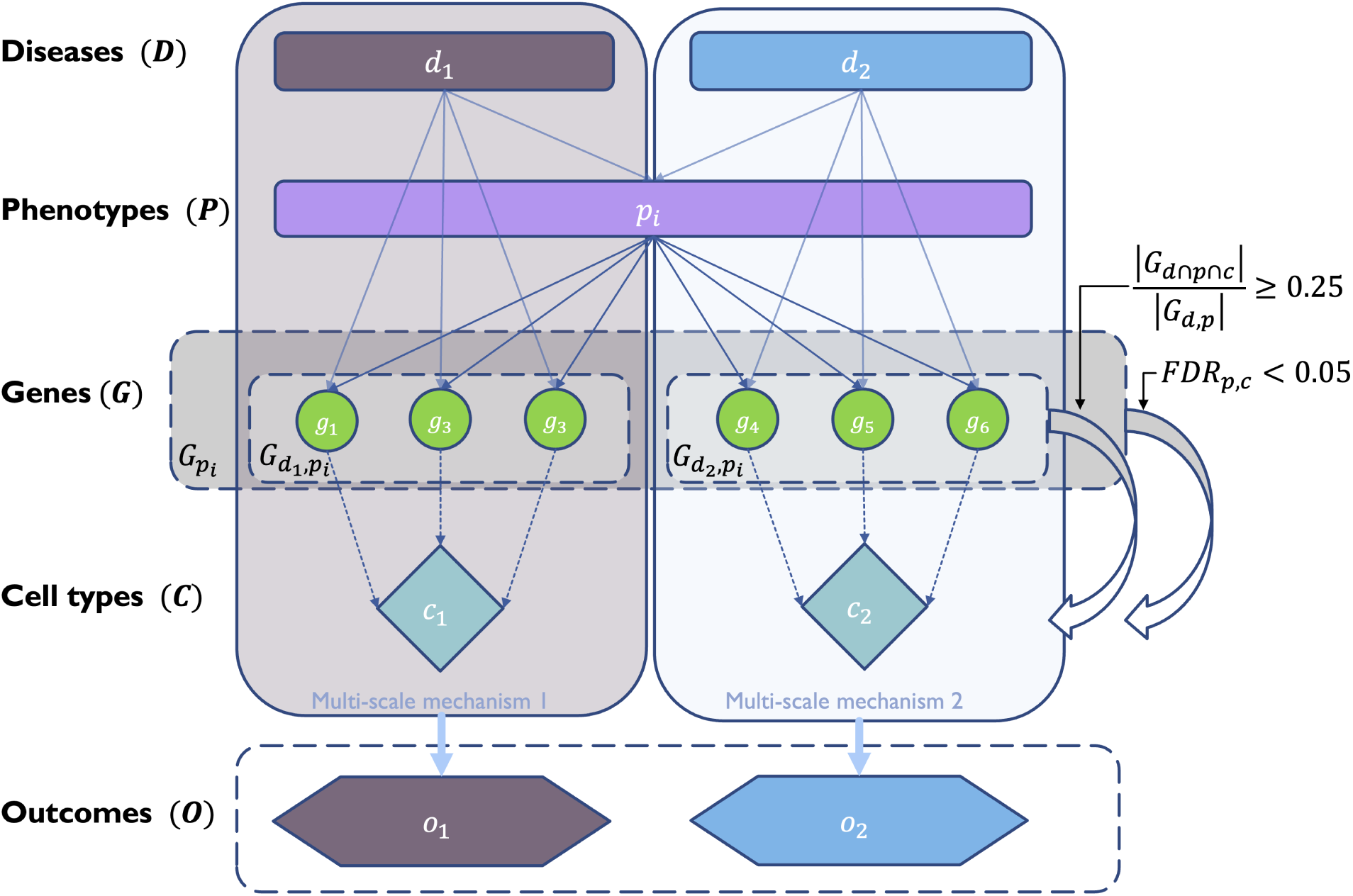
Diagrammatic overview of multi-scale disease investigation strategy. Here we provide an abstract example of differential disease aetiology across multiple scales: diseases (*D*), phenotypes (*P*), cell types (𝐶), genes (*G*), and clinical outcomes (𝑂). In the HPO, genes are assigned to phenotypes via particular diseases (*G*_𝑑*p*_). Therefore, the final gene list for each phenotype is aggregated from across multiple diseases (*G*_*p*_). We performed association tests for all pairwise combinations of cell types and phenotypes and filtered results after multiple testing corrections (FDR<0.05). Each phenotype in the context of a given disease is referred to here as a symptom. Links were established between symptoms and cell types through proportional gene set overlap at a minimum threshold of 25%.

**Figure 11.**
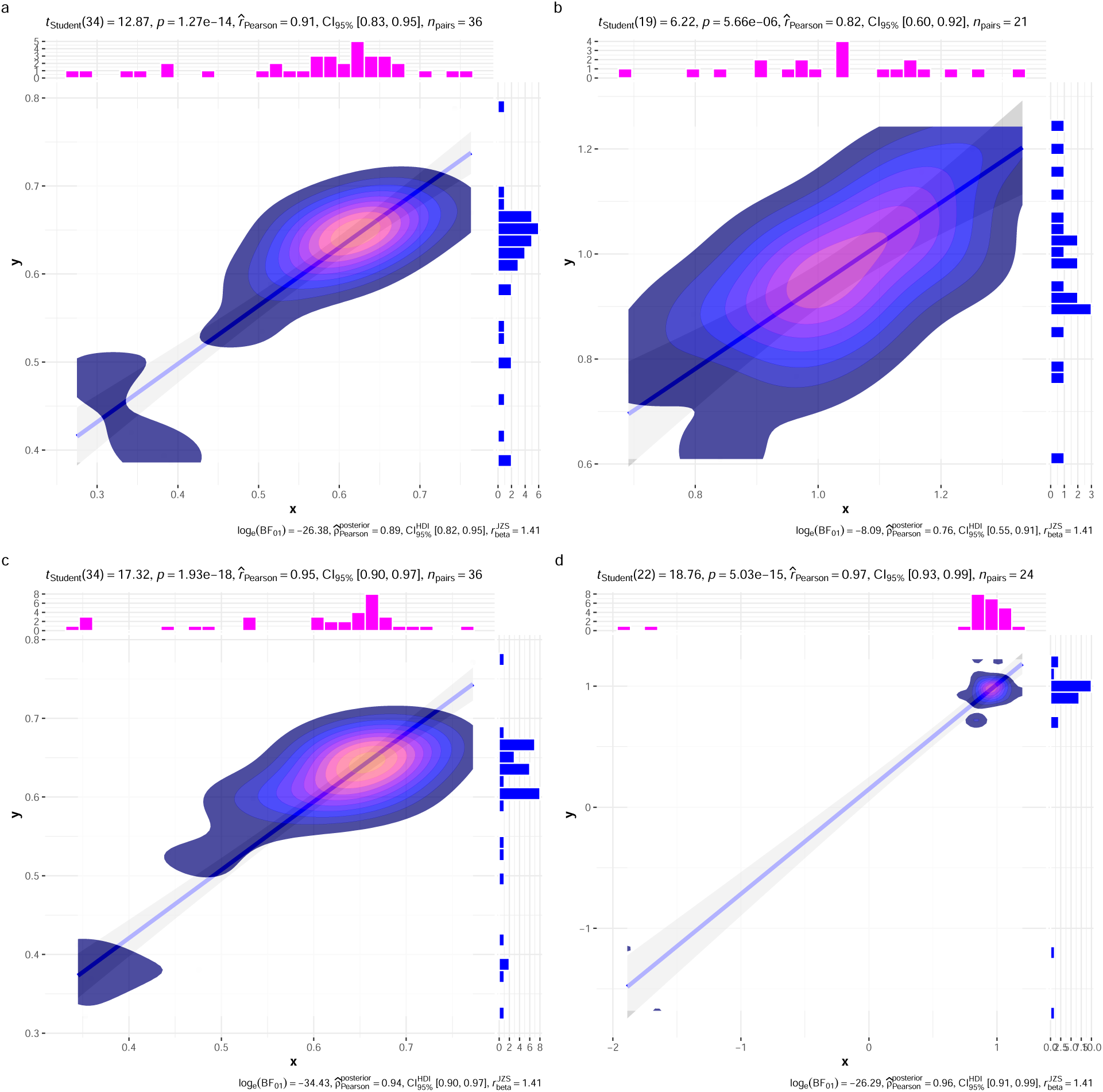
(a) Inter- and intra-dataset validation across the different CellTypeDataset (CTD) and developmental stages. Correlations are computed using Pearson correlation coefficient. Point density is plotted using a 2D kernel density estimate. **a** Correlation between the uncorrected p-values from all phenotype-cell type association tests using the Descartes Human vs. Human Cell Landscape CTDs. **b** Correlation between the *log*_10_(𝑓*o*𝑙𝑑 − 𝑐ℎ𝑎*n*_*g*_𝑒) from significant phenotype-cell type association tests (FDR<0.05) using the Descartes Human vs. Human Cell Landscape CTDs. **c** Correlation between the uncorrected p-values from all phenotype-cell type association tests using the Human Cell Landscape fetal samples vs. Human Cell Landscape adult samples. **d** Correlation between the *log*_10_(*fold-change*) from significant phenotype-cell type association tests (FDR<0.05) using the Human Cell Landscape fetal samples vs. Human Cell Landscape adult samples.

**Figure 12.**
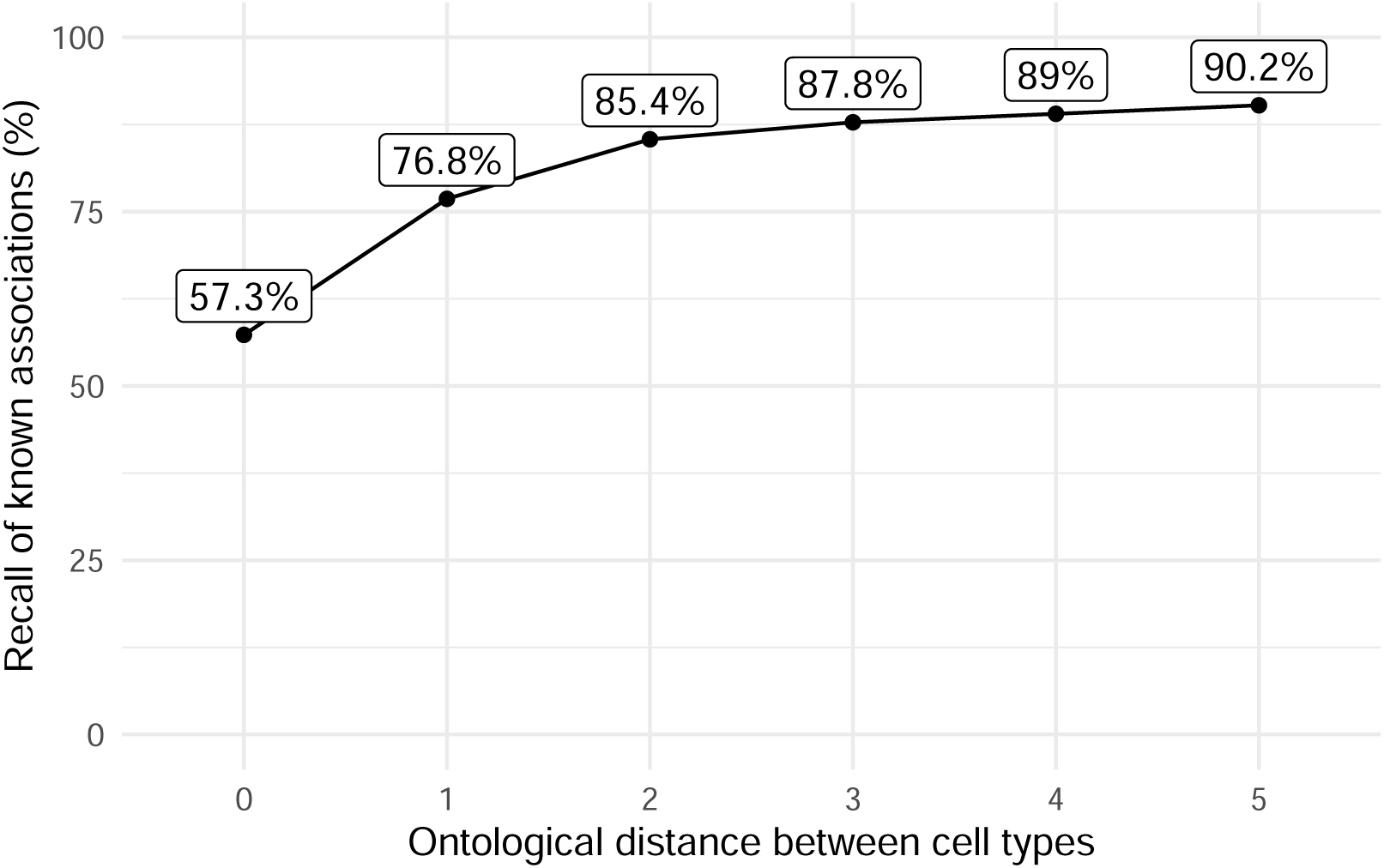
(a) Recall of ground-truth Monarch Knowledge Graph phenotype-cell type relationships at each ontological distance between cell types according to the Cell Ontology.

**Figure 13.**
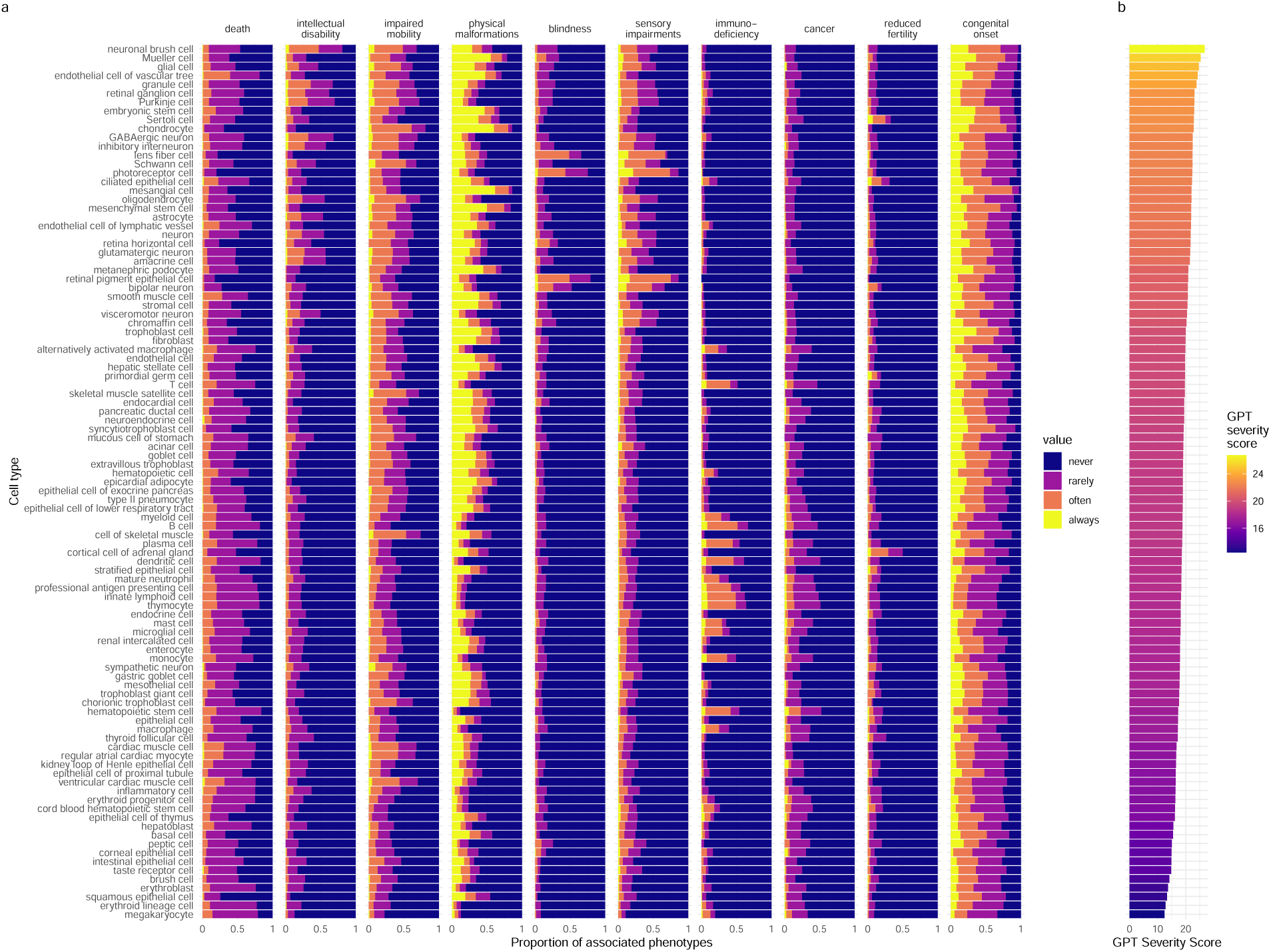
(a) Cell types ordered by the mean severity of the phenotypes they’re associated with. **a**, The disribution of phenotype severity annotation frequencies aggregated by cell type. **b**, The composite severity score, averaged across all phenotypes associated with each cell type.

**Figure 14.**
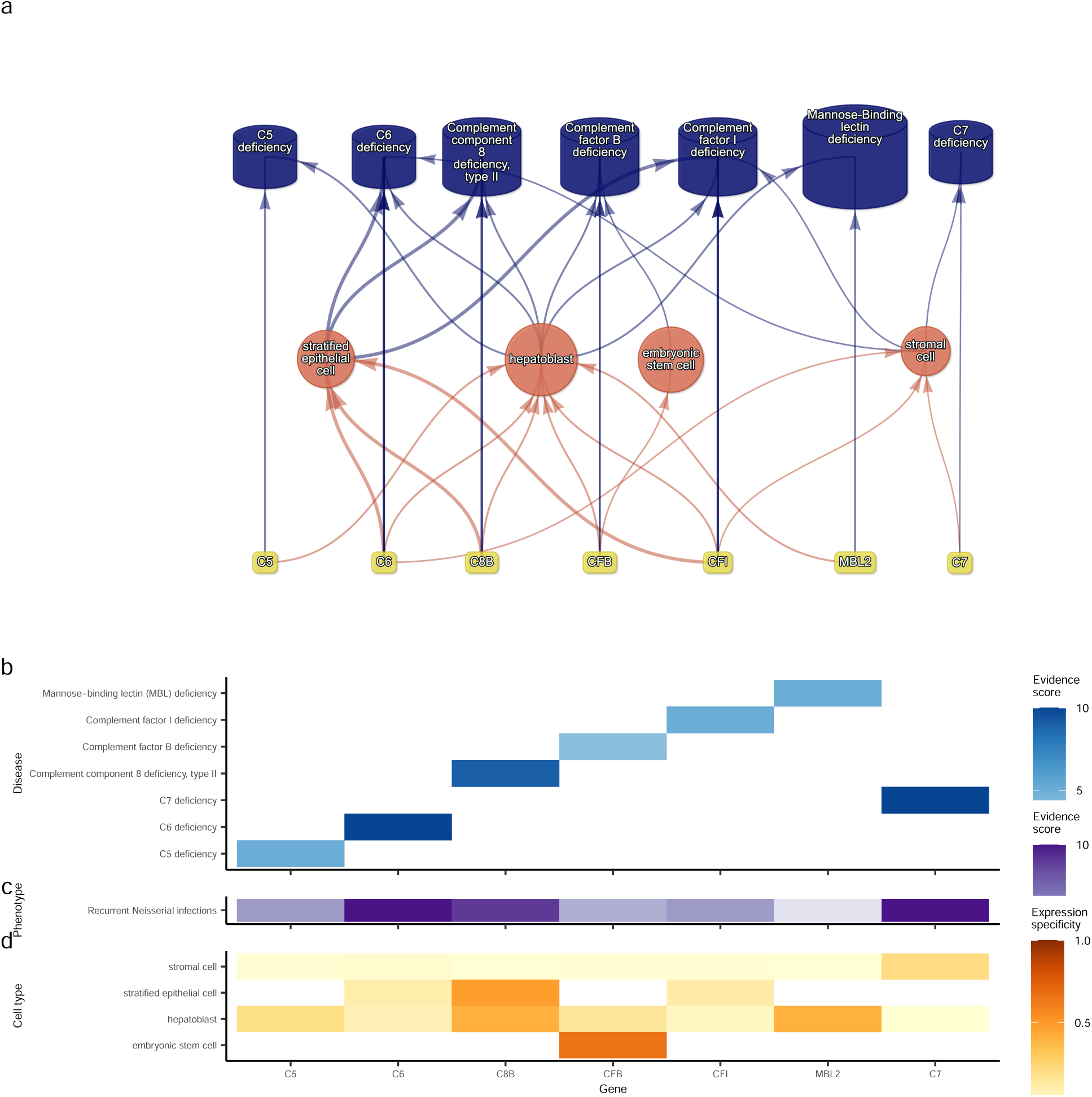
(a) Causal network of recurrent Neisserial infections (RNI) reveals multi-scale disease aetiology. RNI is a phenotype in seven different monogenic diseases caused by disruptions to specific complement system genes. Four cell types were significantly associated with RNI. **a**, One can trace how genes causal for RNI (yellow boxes, bottom) mediate their effects through cell types (orange circles, middle) and diseases (blue cylinders, top). Cell types are connected to RNI via association testing (FDR<0.05). Genes shown here have both strong evidence for a causal role in RNI and high expression specificity in the associated cell type. Cell types can be linked to monogenic diseases via the genes specifically expressed in those cell types (i.e. are in the top 25% of cell type specificity expression quantiles). Nodes are arranged using the Sugiyama algorithm^97^. **b** Expression specificity quantiles (1-40 scale) of each driver gene in each cell type (darker = greater specificity). **c** GenCC-derived eevidence scores between the RNI phenotype and each gene. **d** Expression specificity (0 = least specific, 1 = most specific) of each gene in each cell type.

**Figure 15.**
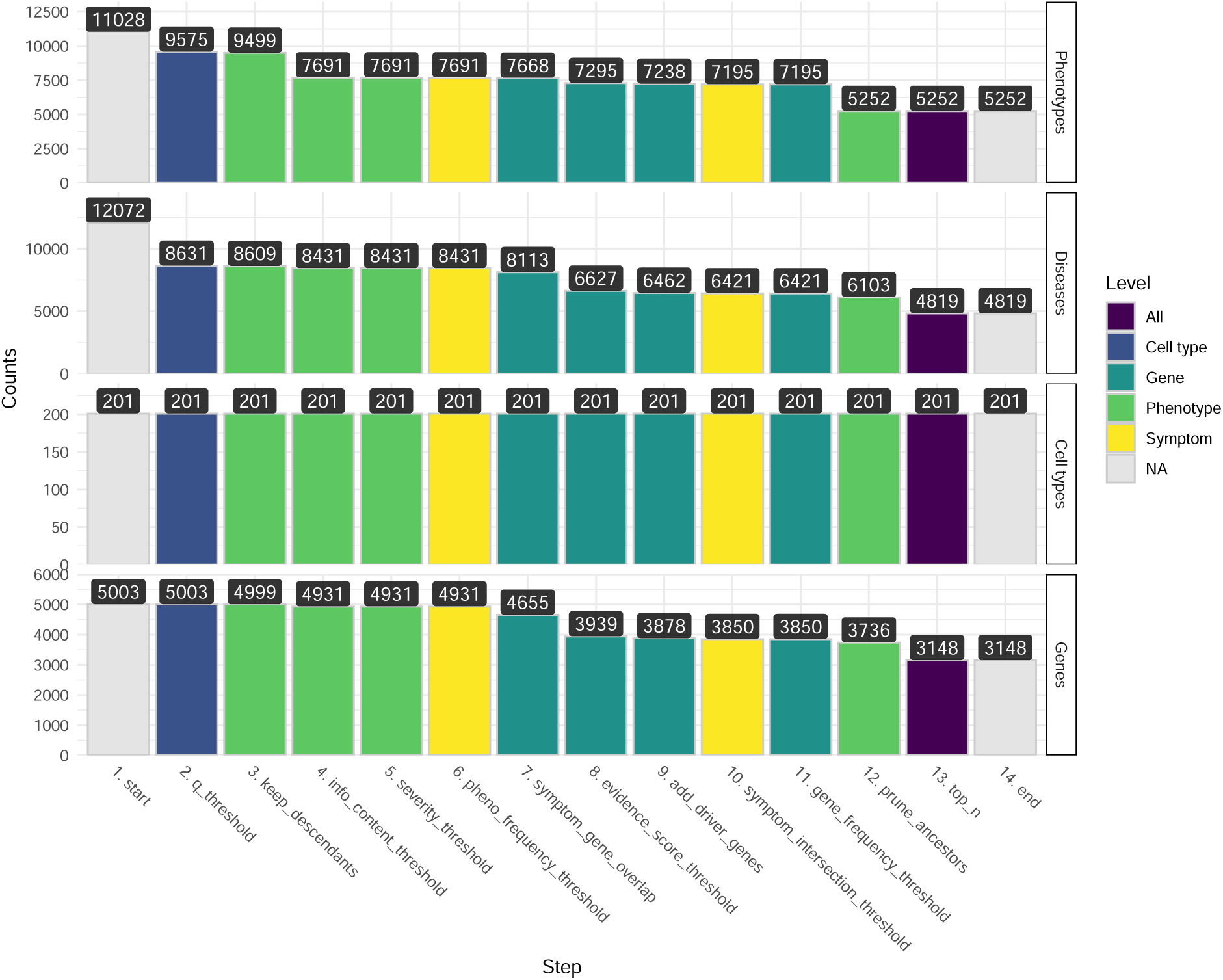
(a) Prioritised target filtering steps. This plot visualises the number of unique phenotype-cell type associations, cell types, genes, and phenotypes (*y-axis*) at each filtering step (*x-axis*) within the multi-scale therapeutic target prioritisation pipeline. Each step in the pipeline can be easily adjusted according to user preference and use case. See Table 3 for descriptions and criterion of each filtering step.

**Figure 16.**
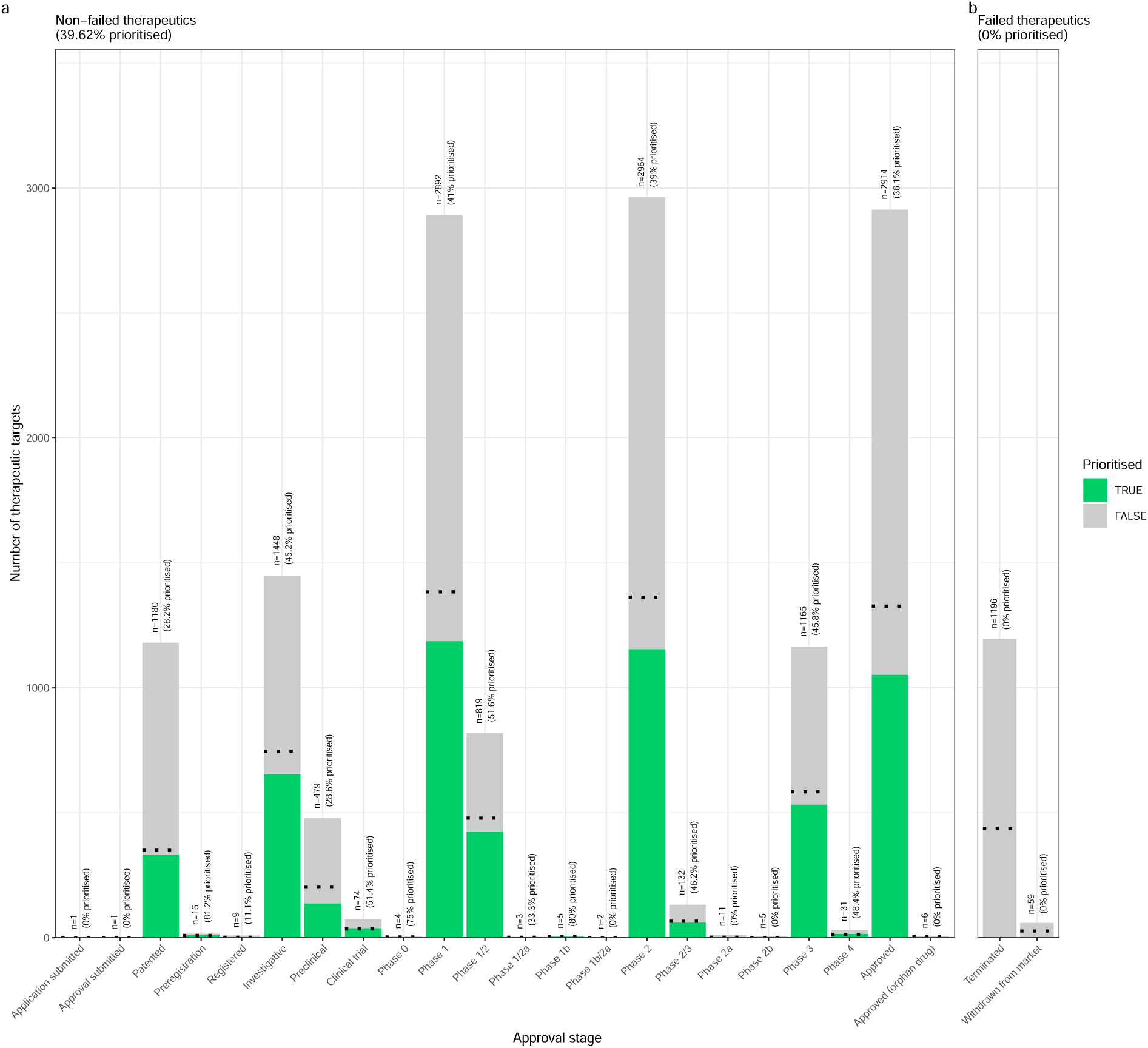
(a) Validation of prioritised therapeutic targets. Proportion of existing all therapy targets (documented in the Therapeutic Target Database) recapitulated by our prioritisation pipeline.

**Figure 17.**
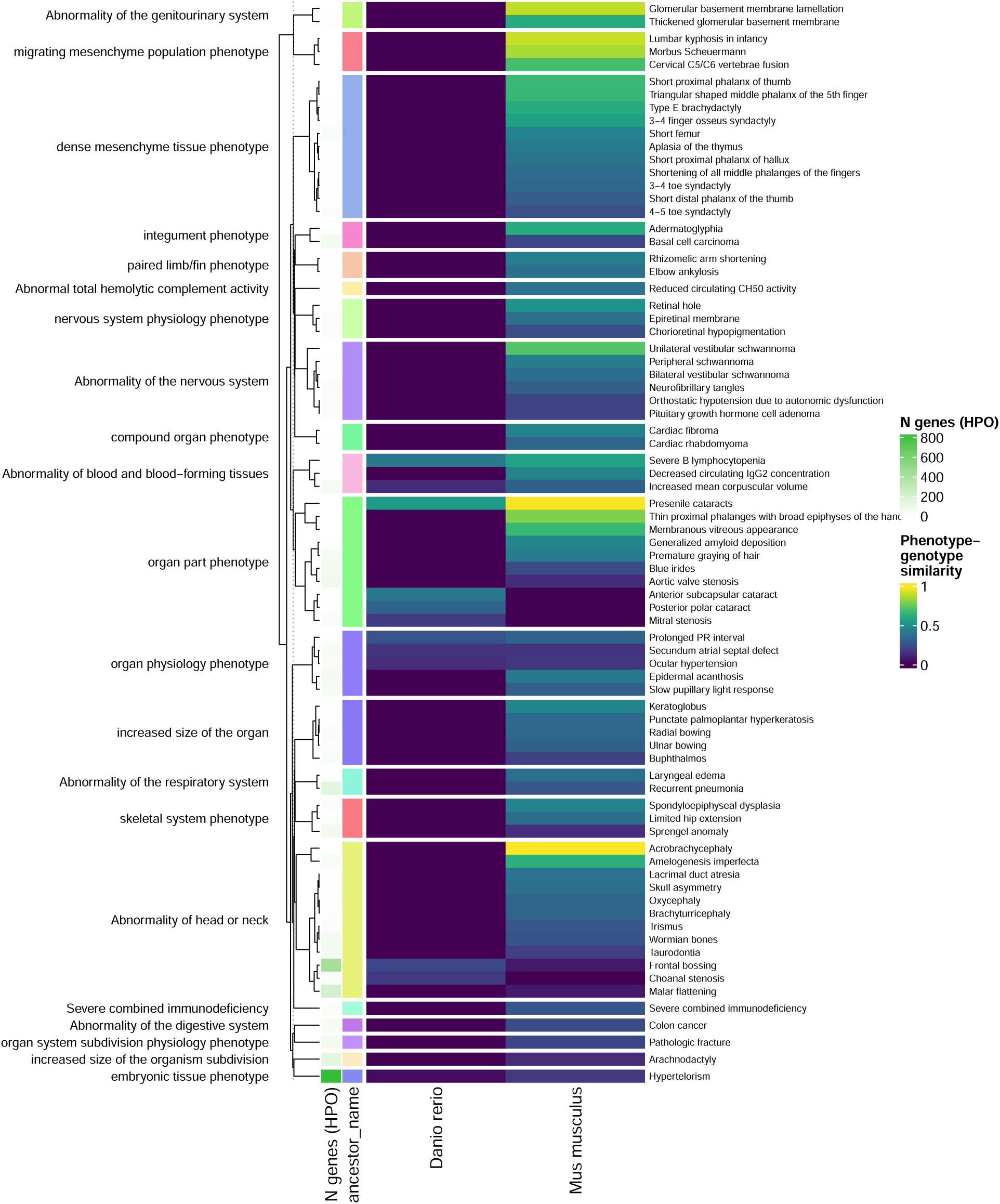
(a) Identification of translatable experimental models. Interspecies translatability of the top 200 human phenotypes nominated by the gene therapy prioritised pipeline. Above, the combined ontological-genotypic similarity score (*SIM*_*og*_) is displayed as the heatmap fill colour stratified by the model organism (*x-axis*). An additional column (“n_genes_db1” on the far left) displays the total number of unique genes annotated to the phenotypic within the HPO. Phenotypes are clustered according to their ontological similarity in the HPO (*y-axis*).

**Figure 18.**
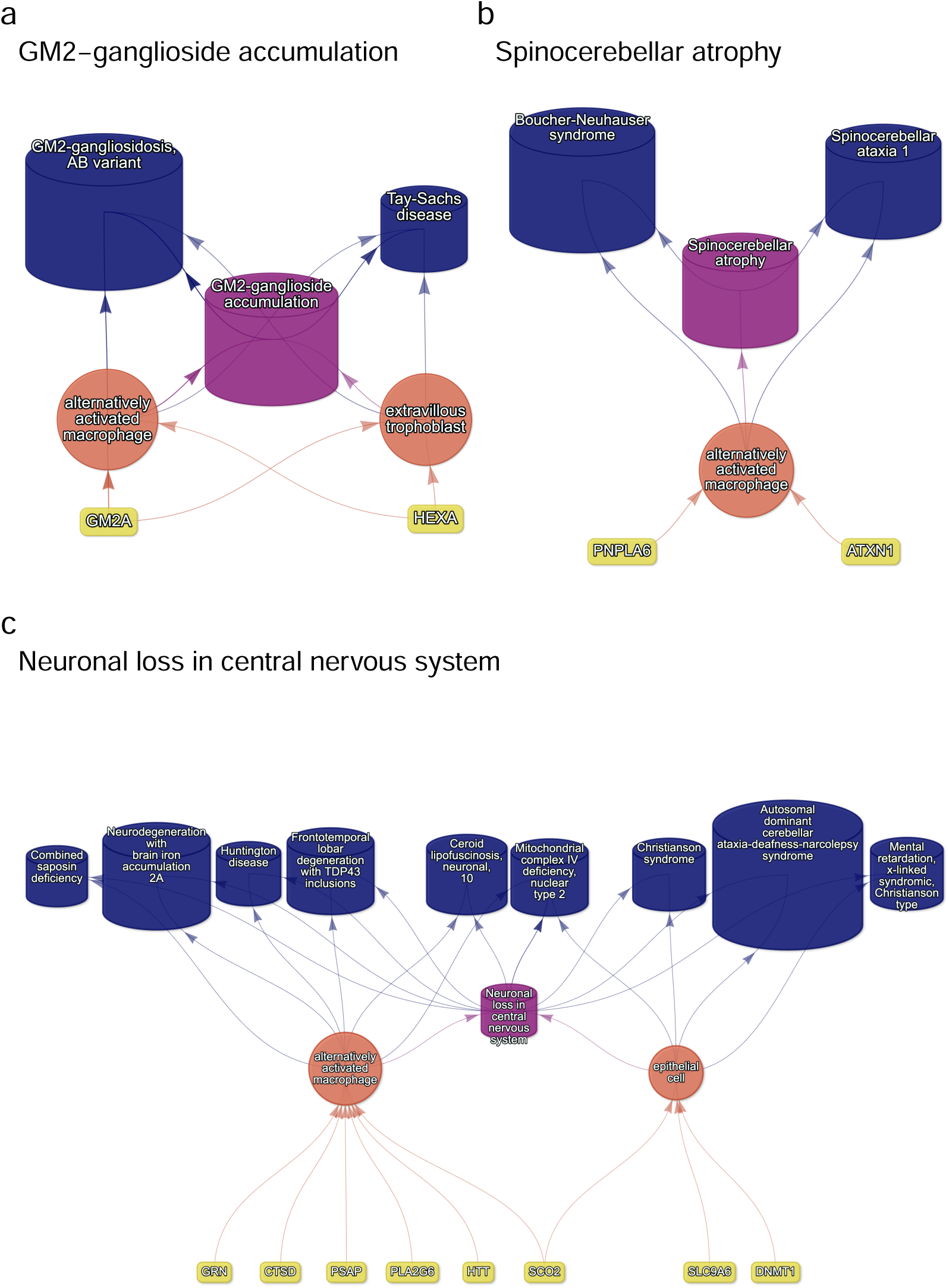
(a) Causal multi-scale networks reveal cell type-specific therapeutic targets. Each disease (blue cylinders) is connected to its phenotype (purple cylinders) based on well-established clinical observations recorded within the HPO^11^. Phenotypes are connected to cell types (orange circles) via association testing between weighted gene sets (FDR<0.05). Each cell type is connected to the prioritised gene targets (yellow boxes) based on the driver gene analysis. The thickness of the edges connecting the nodes represent the (mean) fold-change from the bootstrapped enrichment tests. Nodes were spatially arranged using the Sugiyama algorithm^97^.

**Figure 19:**
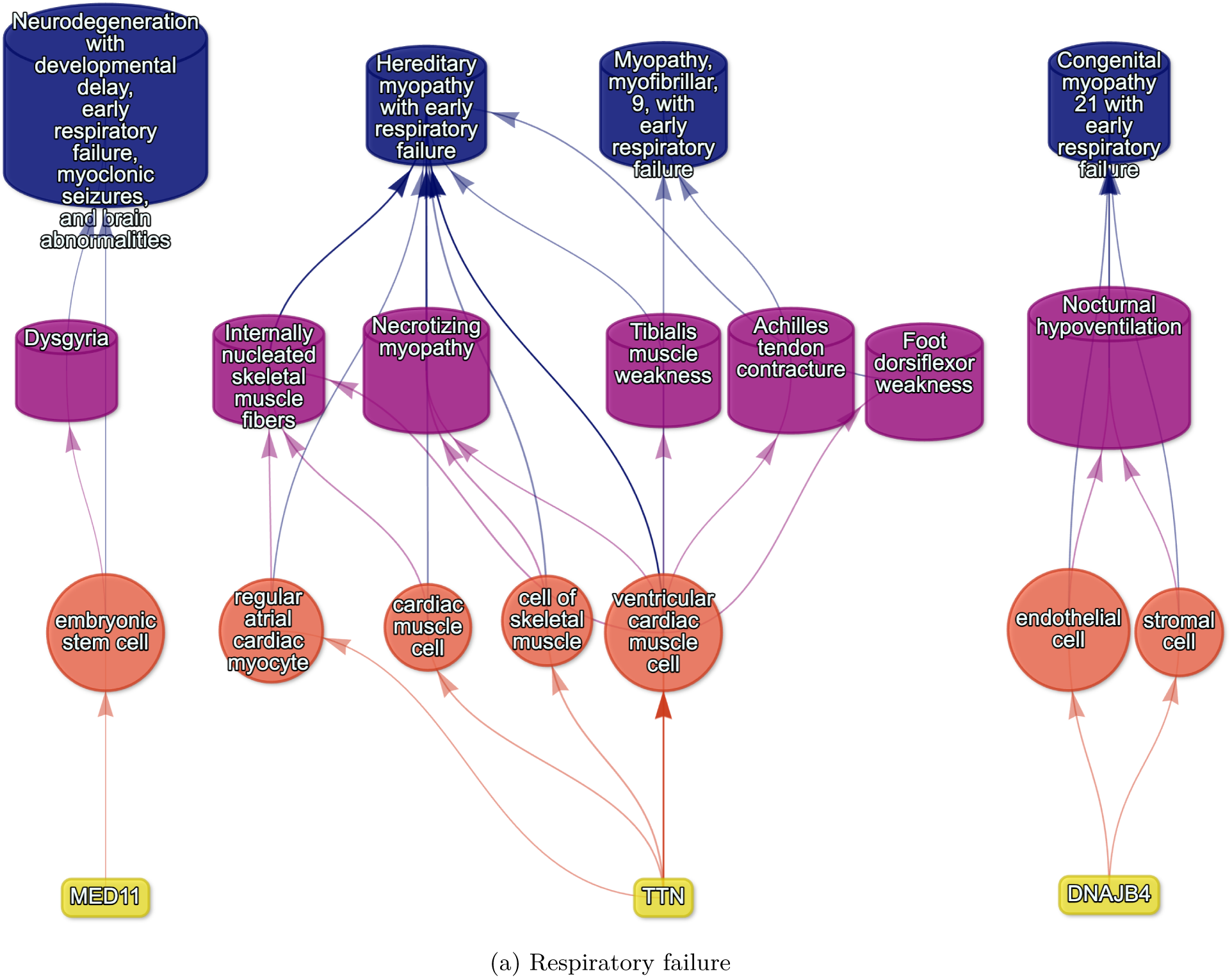
Example cell type-specific gene therapy targets for phenotypes associated with respiratory failure-related diseases. Each disease (blue cylinders) is connected to its phenotype (purple cylinders) based on well-established clinical observations recorded within the HPO^11^.Phenotypes are connected to cell types (red circles) via association testing between weighted gene sets (FDR<0.05). Each cell type is connected to the prioritised gene targets (yellow boxes) based on the driver gene analysis. The thickness of the edges connecting the nodes represent the (mean) fold-change from the bootstrapped enrichment tests. Nodes were spatially arranged using the Sugiyama algorithm^97^.

**Figure 20:**
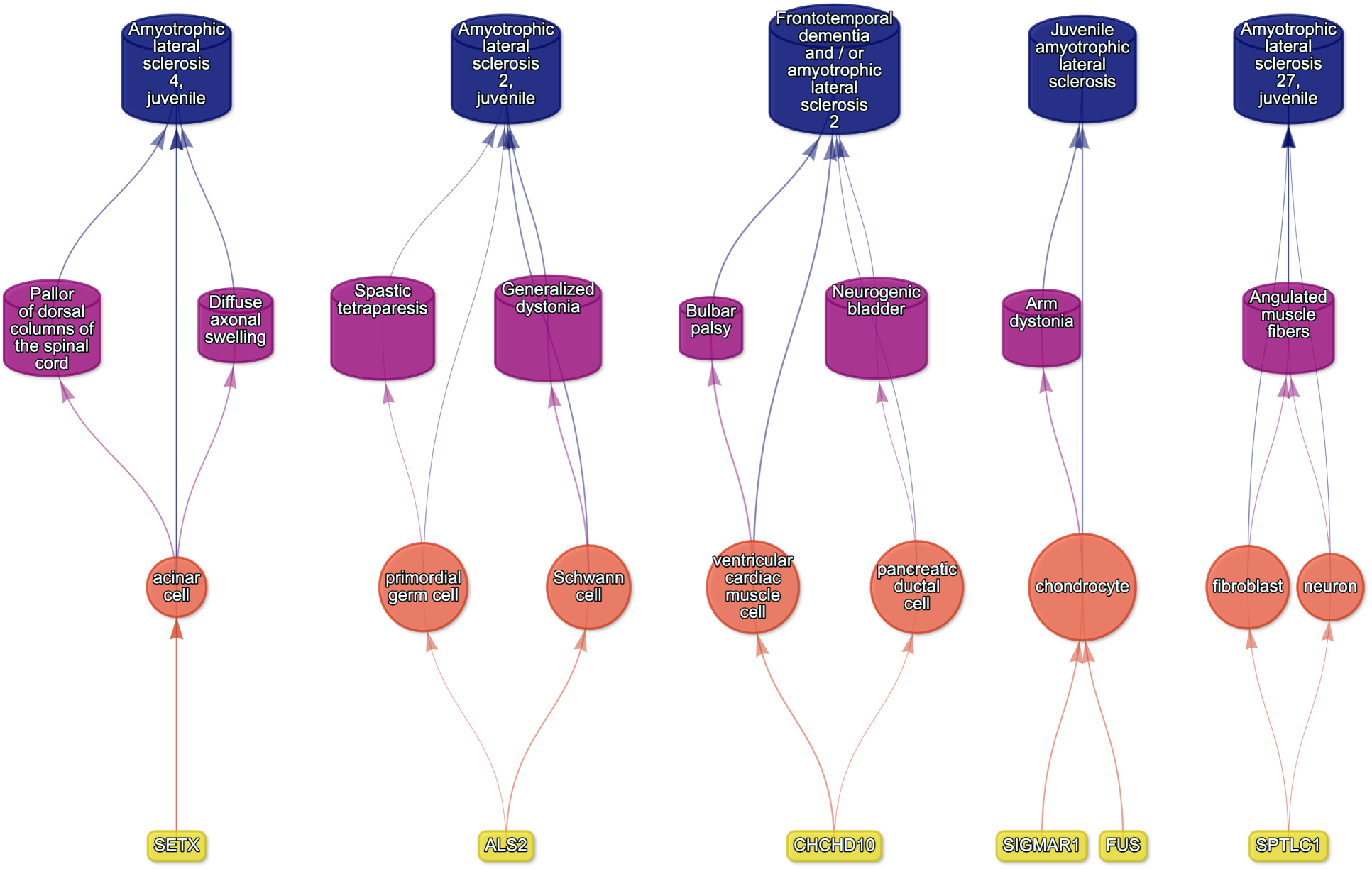
Causal multi-scale network for phenotypes associated with Amyotrophic Lateral Sclerosis (ALS).

**Figure 21:**
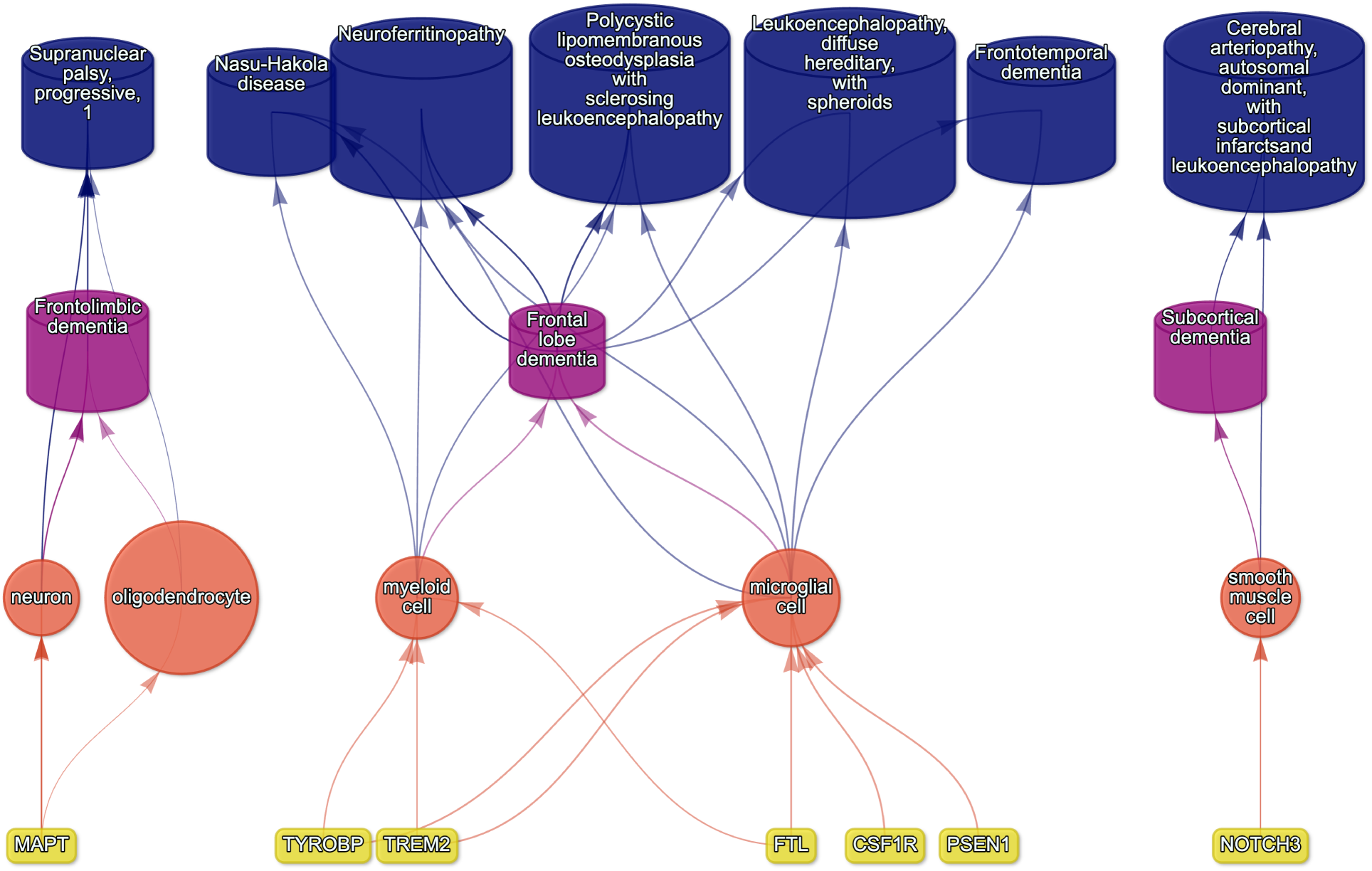
Causal multi-scale network for dementia phenotypes.

**Figure 22:**
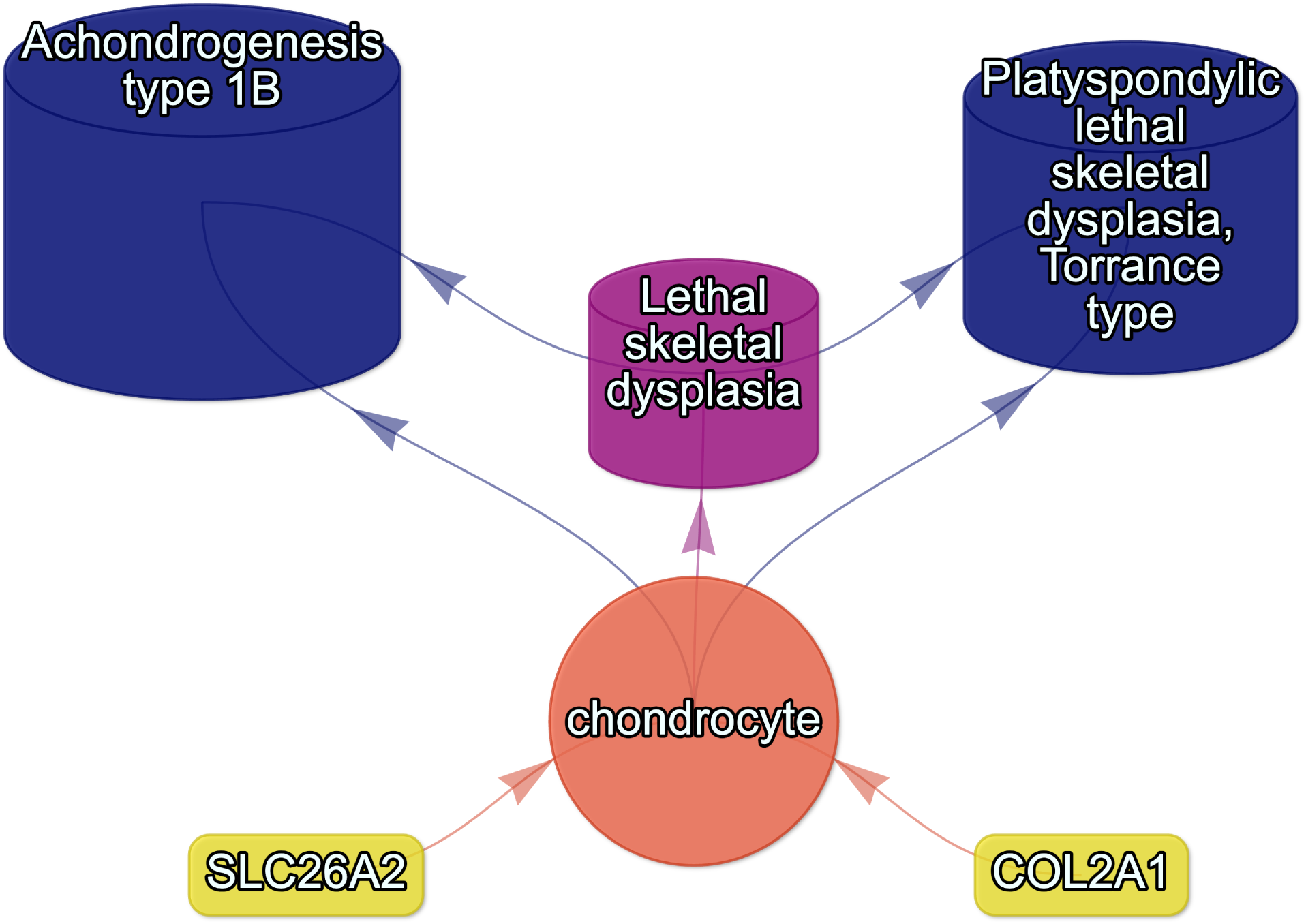
Causal multi-scale network for the phenotype lethal skeletal dysplasia.

**Figure 23:**
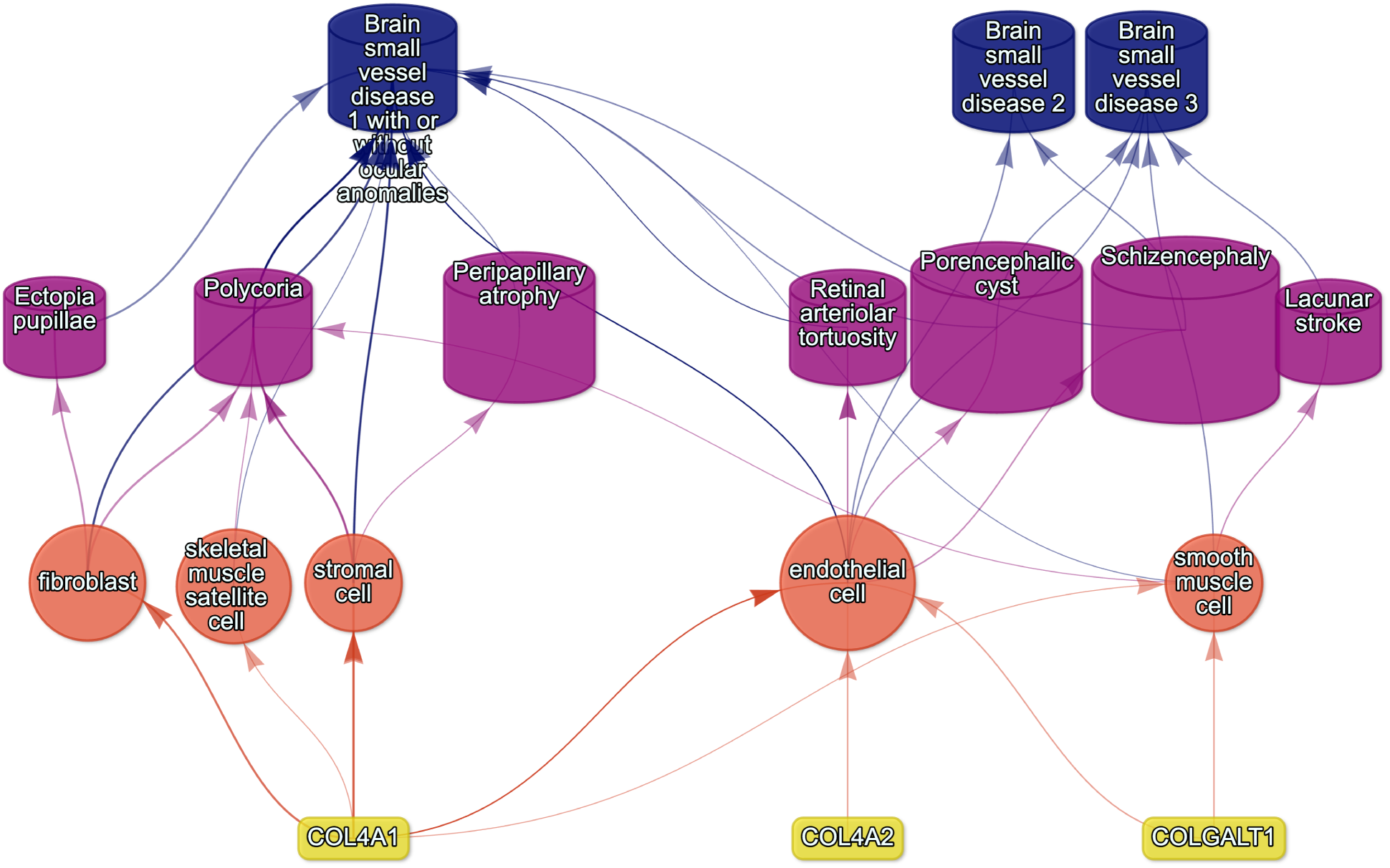
Causal multi-scale network for phenotypes associated with small vessel disease.

**Figure 24:**
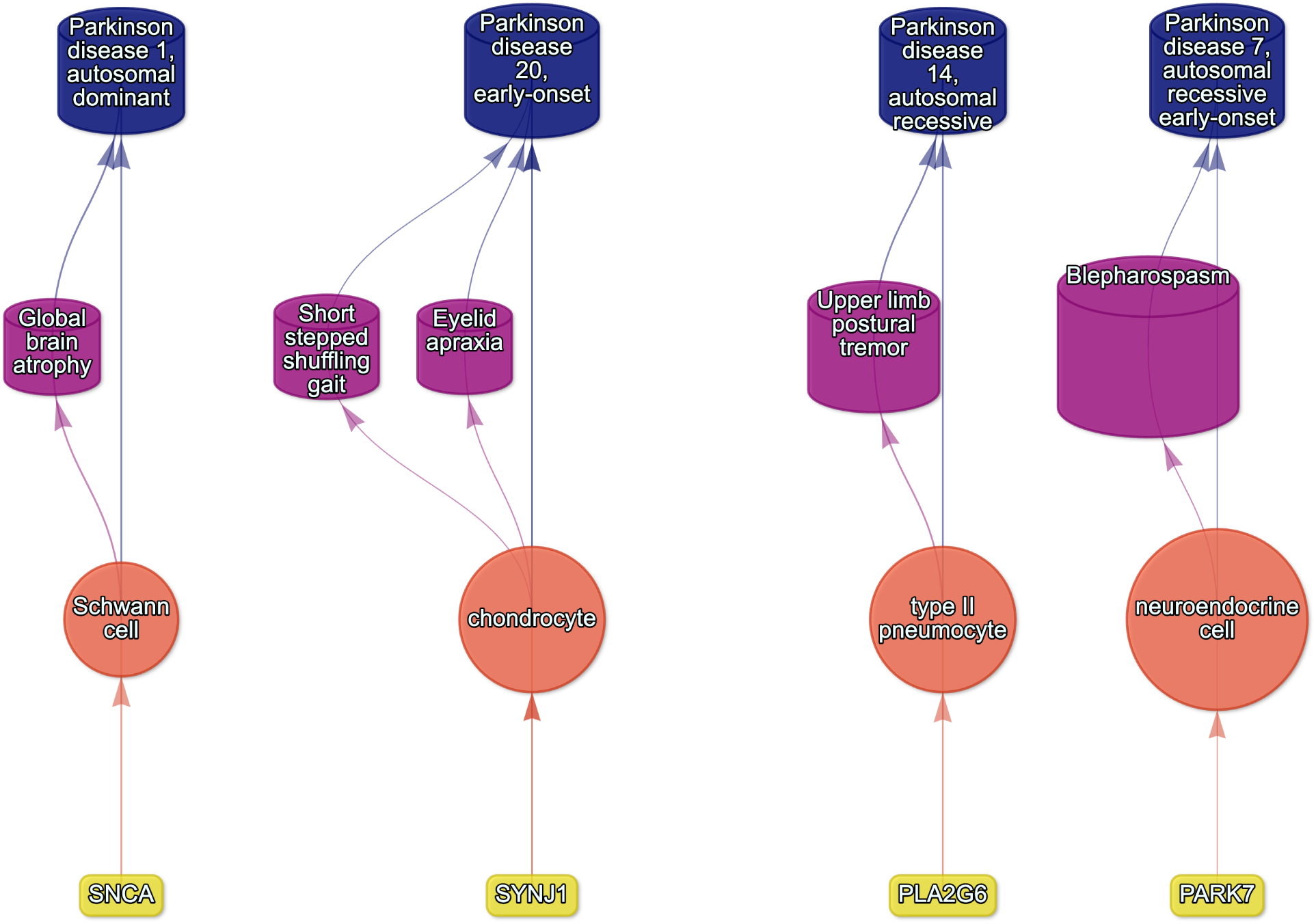
Causal multi-scale network for phenotypes associated with various subtypes of Parkinson’s disease.

**Figure 25:**
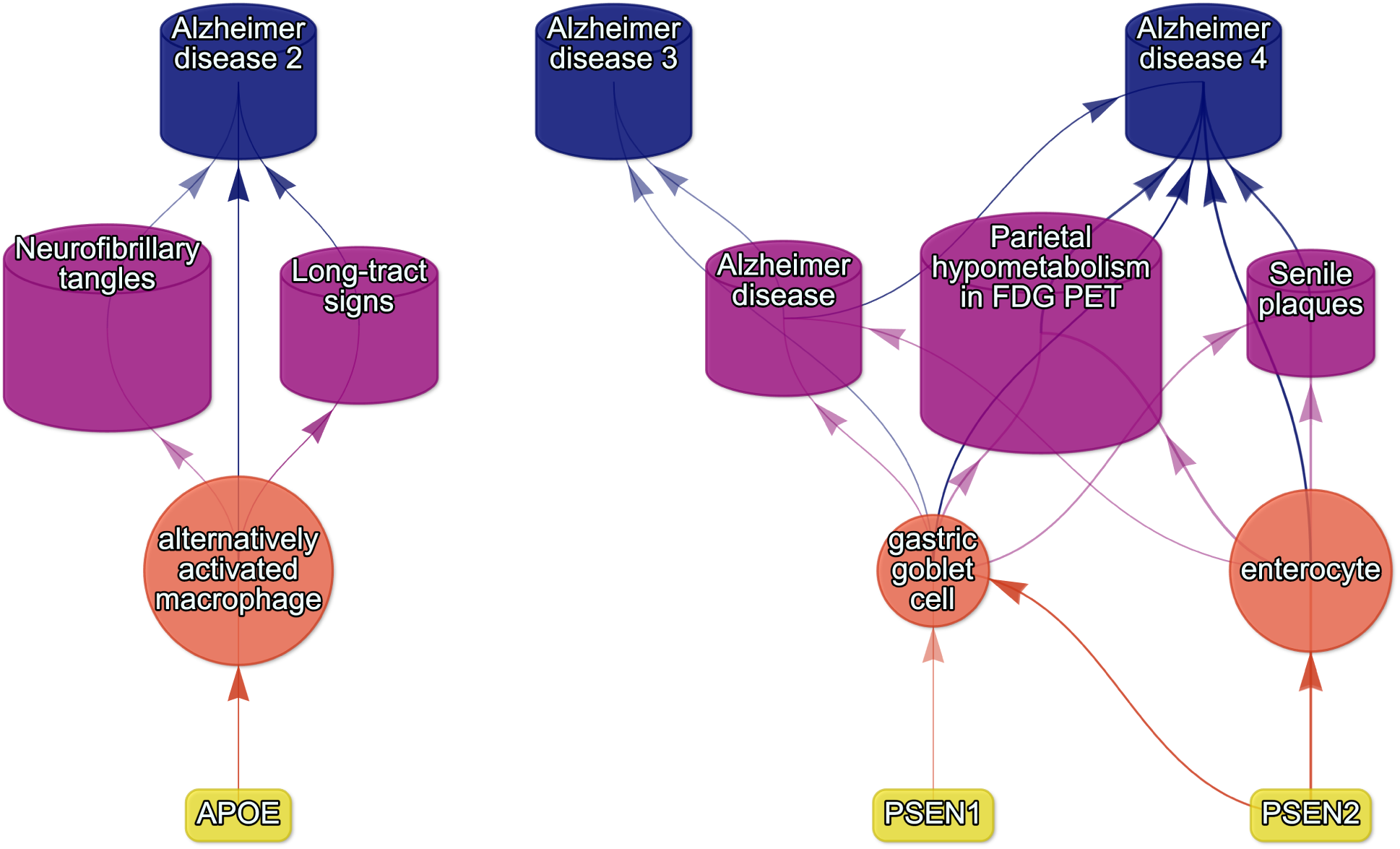
Causal multi-scale network for phenotypes associated with various subtypes of Alzheimer’s disease.

### Supplementary Tables

**Table 1:** Mappings between HPO phenotypes and other medical ontologies. “source” indicates the medical ontology and “distance” indicates the cross-ontology distance. “source terms” and “HPO terms” indicates the number of unique IDs mapped from the source ontology and HPO respectively. “mappings” is the total number of cross-ontology mappings within a given distance. Some IDs may have more than one mapping for a given source due to many-to-many relationships.

| source | distance | source terms | HPO terms | mappings |
| --- | --- | --- | --- | --- |
| ICD10 | 2 | 25 | 23 | 25 |
| ICD10 | 3 | 839 | 876 | 1170 |
| ICD9 | 1 | 21 | 21 | 21 |
| ICD9 | 2 | 434 | 306 | 462 |
| ICD9 | 3 | 1052 | 920 | 1816 |
| SNOMED | 1 | 4413 | 3483 | 4654 |
| SNOMED | 2 | 75 | 21 | 78 |
| SNOMED | 3 | 1796 | 833 | 9605 |
| UMLS | 1 | 12898 | 11601 | 13049 |
| UMLS | 2 | 140 | 113 | 142 |
| UMLS | 3 | 1871 | 1204 | 11021 |

**Table 3:**
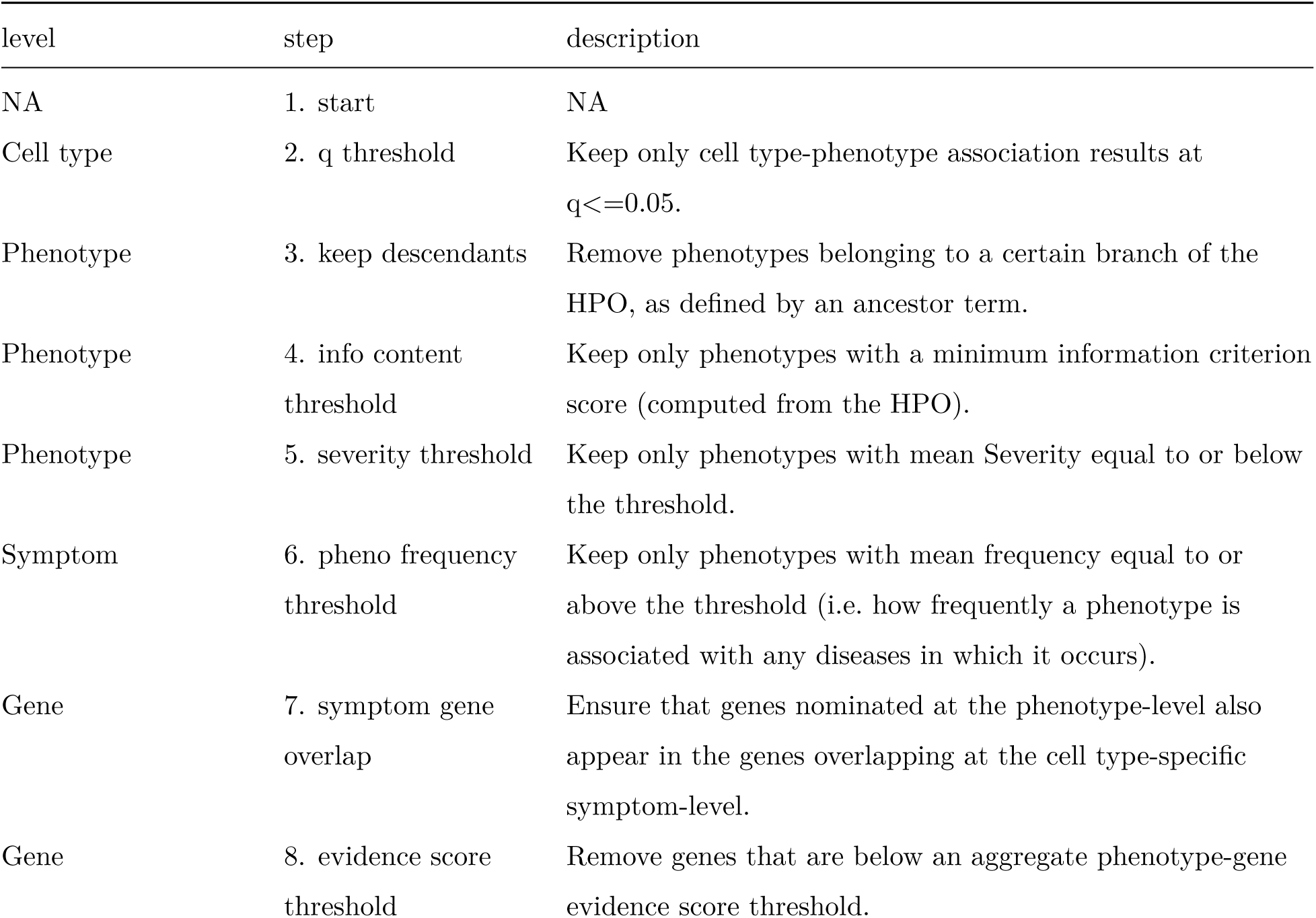
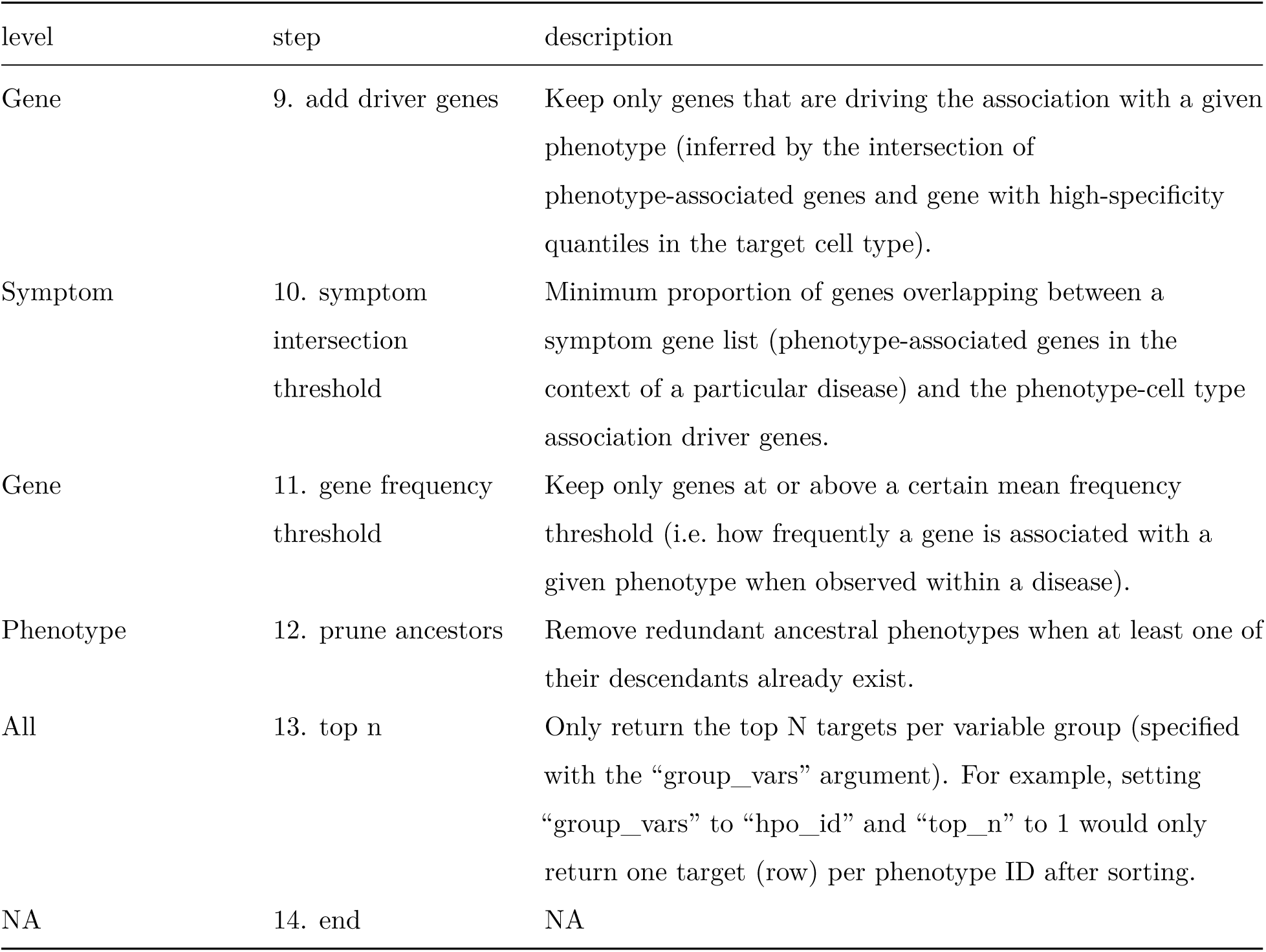
Description of each filtering step performed in the multi-scale therapeutic target prioritisation pipleline. ‘level’ indicates the biological scale at which the step is applied to.

| level | step | description |
| --- | --- | --- |
| NA | 1. start | NA |
| Cell type | 2. q threshold | Keep only cell type-phenotype association results at $q \leq 0.05$. |
| Phenotype | 3. keep descendants | Remove phenotypes belonging to a certain branch of the HPO, as defined by an ancestor term. |
| Phenotype | 4. info content threshold | Keep only phenotypes with a minimum information criterion score (computed from the HPO). |
| Phenotype | 5. severity threshold | Keep only phenotypes with mean Severity equal to or below the threshold. |
| Symptom | 6. pheno frequency threshold | Keep only phenotypes with mean frequency equal to or above the threshold (i.e. how frequently a phenotype is associated with any diseases in which it occurs). |
| Gene | 7. symptom gene overlap | Ensure that genes nominated at the phenotype-level also appear in the genes overlapping at the cell type-specific symptom-level. |
| Gene | 8. evidence score threshold | Remove genes that are below an aggregate phenotype-gene evidence score threshold. |
| Gene | 9. add driver genes | Keep only genes that are driving the association with a given phenotype (inferred by the intersection of phenotype-associated genes and gene with high-specificity quantiles in the target cell type). |
| Symptom | 10. symptom intersection threshold | Minimum proportion of genes overlapping between a symptom gene list (phenotype-associated genes in the context of a particular disease) and the phenotype-cell type association driver genes. |
| Gene | 11. gene frequency threshold | Keep only genes at or above a certain mean frequency threshold (i.e. how frequently a gene is associated with a given phenotype when observed within a disease). |
| Phenotype | 12. prune ancestors | Remove redundant ancestral phenotypes when at least one of their descendants already exist. |
| All | 13. top n | Only return the top N targets per variable group (specified with the “group_vars” argument). For example, setting “group_vars” to “hpo_id” and “top_n” to 1 would only return one target (row) per phenotype ID after sorting. |
| NA | 14. end | NA |

**Table 2:** Summary statistics of enrichment results stratified by single-cell atlas. Summary statistics at multiple levels (tests, cell types, phenotypes, diseases, cell types per phenotype, phenotypes per cell type) stratified by the single-cell atlas that was used as a cell type signature reference (Descartes Human or Human Cell Landscape).

|  | DescartesHuman | HumanCellLandscape | all |
| --- | --- | --- | --- |
| tests significant | 19,929 | 26,585 | 46,514 |
| tests | 848,078 | 1,358,916 | 2,206,994 |
| tests significant (%) | 2.35 | 1.96 | 2.11 |
| cell types significant | 77 | 124 | 201 |
| cell types | 77 | 124 | 201 |
| cell types significant (%) | 100 | 100 | 100 |
| phenotypes significant | 7,340 | 9,049 | 9,575 |
| phenotypes tested | 11,014 | 10,959 | 11,028 |
| phenotypes | 11,047 | 11,047 | 11,047 |
| phenotypes significant (%) | 66.4 | 81.9 | 86.7 |
| diseases significant | 8,628 | 8,627 | 8,628 |
| diseases | 8,631 | 8,631 | 8,631 |
| diseases significant (%) | 100 | 100 | 100 |
| cell types per phenotype (mean) | 1.81 | 2.43 | 4.22 |
| cell types per phenotype (median) | 1 | 2 | 3 |
| cell types per phenotype (min) | 0 | 0 | 0 |
| cell types per phenotype (max) | 31 | 28 | 59 |
| phenotypes per cell type (mean) | 259 | 214 | 231 |
| phenotypes per cell type (median) | 252 | 200 | 209 |
| phenotypes per cell type (min) | 71 | 57 | 57 |
| phenotypes per cell type (max) | 696 | 735 | 735 |

**Table 4:** Cross-ontology mappings between HPO and CL branches. The last two columns represent the number of cell types that were overrepresented in the on-target HPO branch and the total number of cell types in that branch. A disaggregated version of this table with all descendant cell type names is available in Table 6.

| HPO branch | Phenotypes |  | Cell types |  |
| --- | --- | --- | --- | --- |
|  | (total) | CL branch | (overrepresented) | (total) |
| Abnormality of the cardiovascular system | 673 | cardiocyte | 5 | 6 |
| Abnormality of the endocrine system | 291 | endocrine cell | 3 | 4 |
| Abnormality of the eye | 721 | photoreceptor cell/retinal cell | 5 | 5 |
| Abnormality of the immune system | 255 | leukocyte | 14 | 14 |
| Abnormality of the musculoskeletal system | 2155 | cell of skeletal muscle/chondrocyte | 4 | 4 |
| Abnormality of the nervous system | 1647 | neural cell | 17 | 24 |
| Abnormality of the respiratory system | 292 | respiratory epithelial cell/epithelial cell of lung | 3 | 3 |

**Table 5:** Encodings for GenCC evidence scores. Assigned numeric values for the GenCC evidence levels.

| classification_curie | classification_title | encoding |
| --- | --- | --- |
| GENCC:100001 | Definitive | 6 |
| GENCC:100002 | Strong | 5 |
| GENCC:100003 | Moderate | 4 |
| GENCC:100009 | Supportive | 3 |
| GENCC:100004 | Limited | 2 |
| GENCC:100005 | Disputed Evidence | 1 |
| GENCC:100008 | No Known Disease Relationship | 0 |
| GENCC:100006 | Refuted Evidence | 0 |

**Table 6:** On-target cell types for each Human Phenotype Ontology (HPO) ancestral branch. Cell type-phenotype branch pairings were manually curated by comparing high-level HPO terms to terms within the Cell Ontology (CL). Each HPO branch is shown as bolded row dividers. Ancestral CL branch names are shown in the first column, along with the specific CL names and IDs.

| CL branch | CL name | CL ID |
| --- | --- | --- |
| **Abnormality of the cardiovascular system** |  |  |
| cardiocyte | cardiac muscle cell | CL:0000746 |
| cardiocyte | regular atrial cardiac myocyte | CL:0002129 |
| cardiocyte | endocardial cell | CL:0002350 |
| cardiocyte | epicardial adipocyte | CL:1000309 |
| cardiocyte | ventricular cardiac muscle cell | CL:2000046 |
| **Abnormality of the endocrine system** |  |  |
| endocrine cell | endocrine cell | CL:0000163 |
| endocrine cell | neuroendocrine cell | CL:0000165 |
| endocrine cell | chromaffin cell | CL:0000166 |
| **Abnormality of the eye** |  |  |
| photoreceptor cell / retinal cell | photoreceptor cell | CL:0000210 |
| photoreceptor cell / retinal cell | amacrine cell | CL:0000561 |
| photoreceptor cell / retinal cell | Mueller cell | CL:0000636 |
| photoreceptor cell / retinal cell | retinal pigment epithelial cell | CL:0002586 |
| **Abnormality of the immune system** |  |  |
| leukocyte | T cell | CL:0000084 |
| leukocyte | mature neutrophil | CL:0000096 |
| leukocyte | mast cell | CL:0000097 |
| leukocyte | microglial cell | CL:0000129 |
| leukocyte | professional antigen presenting cell | CL:0000145 |
| leukocyte | macrophage | CL:0000235 |
| leukocyte | B cell | CL:0000236 |
| leukocyte | dendritic cell | CL:0000451 |
| leukocyte | monocyte | CL:0000576 |
| leukocyte | plasma cell | CL:0000786 |
| leukocyte | alternatively activated macrophage | CL:0000890 |
| leukocyte | thymocyte | CL:0000893 |
| leukocyte | innate lymphoid cell | CL:0001065 |
| **Abnormality of the musculoskeletal system** |  |  |
| cell of skeletal muscle / chondrocyte | chondrocyte | CL:0000138 |
| cell of skeletal muscle / chondrocyte | cell of skeletal muscle | CL:0000188 |
| cell of skeletal muscle / chondrocyte | skeletal muscle satellite cell | CL:0000594 |
| **Abnormality of the nervous system** |  |  |
| neural cell | bipolar neuron | CL:0000103 |
| neural cell | granule cell | CL:0000120 |
| neural cell | Purkinje cell | CL:0000121 |
| neural cell | glial cell | CL:0000125 |
| neural cell | astrocyte | CL:0000127 |
| neural cell | oligodendrocyte | CL:0000128 |
| neural cell | microglial cell | CL:0000129 |
| neural cell | neuroendocrine cell | CL:0000165 |
| neural cell | chromaffin cell | CL:0000166 |
| neural cell | photoreceptor cell | CL:0000210 |
| neural cell | inhibitory interneuron | CL:0000498 |
| neural cell | neuron | CL:0000540 |
| neural cell | neuronal brush cell | CL:0000555 |
| neural cell | amacrine cell | CL:0000561 |
| neural cell | GABAergic neuron | CL:0000617 |
| neural cell | Mueller cell | CL:0000636 |
| neural cell | glutamatergic neuron | CL:0000679 |
| neural cell | retinal ganglion cell | CL:0000740 |
| neural cell | retina horizontal cell | CL:0000745 |
| neural cell | Schwann cell | CL:0002573 |
| neural cell | retinal pigment epithelial cell | CL:0002586 |
| neural cell | visceromotor neuron | CL:0005025 |
| neural cell | sympathetic neuron | CL:0011103 |
| **Abnormality of the respiratory system** |  |  |
| respiratory epithelial cell / epithelial cell of lung | type II pneumocyte | CL:0002063 |
| respiratory epithelial cell / epithelial cell of lung | epithelial cell of lower respiratory tract | CL:0002632 |

**Table 7:**
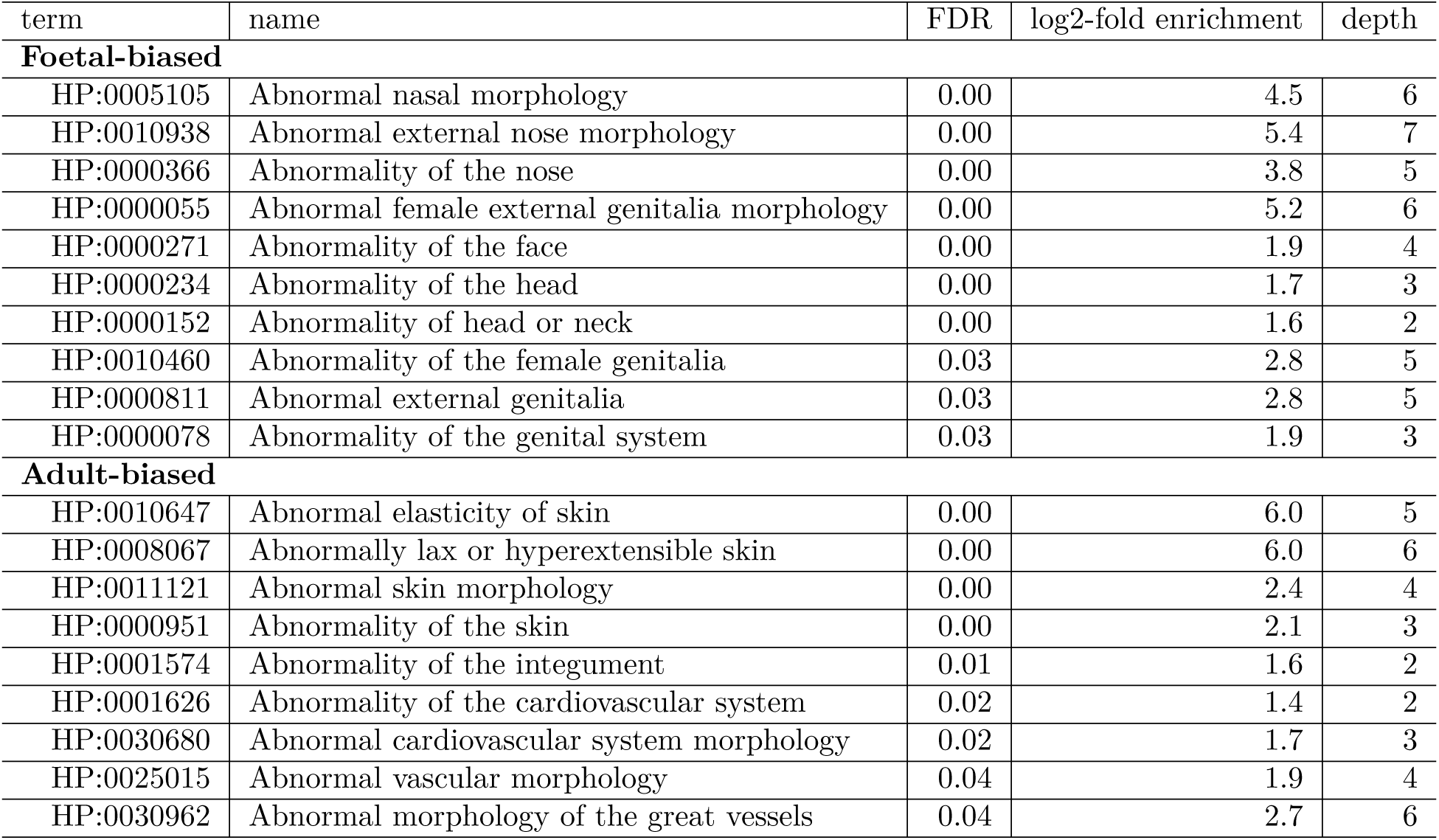
Some HPO phenotype categories or more biased towards foetal- or adult-versions of the same cell type. We took the top 50 phenotypes with the greatest bias towards foetal-cell type associations (“Foetal-biased”) and the greatest bias towards adult-cell type associations (“Adult-biased”) and fed each list of terms into ontological enrichment tests to get a summary of the representative HPO branches for each group. The phenotypes most biased towards associations with only the foetal versions of cell type and those biased towards the adult versions of cell types. “FDR” is the False Discovery Rate-adjusted p-value from the enrichment test, “log2-fold enrichment” is the log2 fold-change from the enrichment test, and “depth” is the depth of the enriched HPO term in the ontology.

**Table 8:** Examples of specific phenotypes that are most biased towards associations with only the foetal versions of cell types (“Foetal-biased”) and those biased towards the adult versions of cell types (“Adult-biased”). “p-value difference” is the difference in the association p-values between the foetal and adult version of the equivalent cell type (foetal-adult bias ∶ *p*_𝑎𝑑𝑢𝑙*t*_ − *p*_𝑓*o*𝑒*t*𝑎𝑙_ = Δ*p* ∈ [−1, 1]).

| HPO name | HPO ID | CL ID | CL name | p-value difference |
| --- | --- | --- | --- | --- |
| **Foetal-biased** |  |  |  |  |
| Short middle phalanx of the 2nd finger | HP:0009577 | CL:0000138 | chondrocyte | 0.99 |
| Abnormal morphology of the nasal alae | HP:0000429 | CL:0000057 | fibroblast | 0.95 |
| Abnormal labia minora morphology | HP:0012880 | CL:0000499 | stromal cell | 0.94 |
| Acromesomelia | HP:0003086 | CL:0000138 | chondrocyte | 0.93 |
| Left atrial isomerism | HP:0011537 | CL:0000163 | endocrine cell | 0.92 |
| Fixed facial expression | HP:0005329 | CL:0000499 | stromal cell | 0.92 |
| Migraine without aura | HP:0002083 | CL:0000163 | endocrine cell | 0.92 |
| Truncal ataxia | HP:0002078 | CL:0000163 | endocrine cell | 0.92 |
| Anteverted nares | HP:0000463 | CL:0000057 | fibroblast | 0.91 |
| Short 1st metacarpal | HP:0010034 | CL:0000138 | chondrocyte | 0.90 |
| **Adult-biased** |  |  |  |  |
| Symblepharon | HP:0430007 | CL:0000138 | chondrocyte | -0.97 |
| Abnormally lax or hyperextensible skin | HP:0008067 | CL:0000057 | fibroblast | -0.94 |
| Reduced bone mineral density | HP:0004349 | CL:0000057 | fibroblast | -0.94 |
| Paroxysmal supraventricular tachycardia | HP:0004763 | CL:0000138 | chondrocyte | -0.93 |
| Lack of skin elasticity | HP:0100679 | CL:0000057 | fibroblast | -0.92 |
| Excessive wrinkled skin | HP:0007392 | CL:0000057 | fibroblast | -0.91 |
| Bruising susceptibility | HP:0000978 | CL:0000057 | fibroblast | -0.91 |
| Corneal opacity | HP:0007957 | CL:0000057 | fibroblast | -0.90 |
| Broad skull | HP:0002682 | CL:0000138 | chondrocyte | -0.90 |
| Emphysema | HP:0002097 | CL:0000057 | fibroblast | -0.89 |

## Notes

### Competing Interest Statement

The authors have declared no competing interest.

### Author Declarations

Human Phenotype Ontology https://hpo.jax.org Descartes scRNA-seq atlas https://descartes.brotmanbaty.org/bbi/human-gene-expression-during-development

### Summary of Updates

We have added a pipeline to systematically prioritise gene therapy targets based on the strength of evidence linking phenotypes to specific cell types as well as the severity of the phenotypes themselves (using novel data annotations described herein). We have also significantly re-egineered the Rare Disease Celltyping Portal for improved performance and broader utility.

